# Stochastic epidemic models and their link with methods from survival analysis

**DOI:** 10.1101/2024.02.18.24302991

**Authors:** Hein Putter, Jelle Goeman, Jacco Wallinga

## Abstract

Compartmental models based on ordinary differential equations (ODE’s) quantifying the interactions between susceptible, infectious, and recovered individuals within a population have played an important role in infectious disease modeling. The aim of the present paper is to explain the link between stochastic epidemic models based on the susceptible-infectious-recovered (SIR) model, and methods from survival analysis. We illustrate how standard software for survival analysis in the statistical language R can be used to estimate pivotal parameters in the stochastic SIR model in the very much idealized situation where the epidemic is completely observed. Extensions incorporating interventions, age structure and heterogeneity are explored and illustrated.

## 1 Introduction

Mathematical modeling has played a pivotal role in understanding and managing the dynamics of infectious diseases. In particular, models based on ordinary differential equations (ODE’s) quantifying the interactions between susceptible, infectious, and recovered individuals within a population, the SIR model (Kermack and McKendrick 1927) and its extensions have had enormous impact. These models allow public health professionals to predict the course of an outbreak and assess the impact of various interventions. Moreover, mathematical modeling provides a valuable tool for scenario planning, allowing for the simulation of different outbreak scenarios and the evaluation of their respective outcomes. We refer to Anderson and May (1991) and Diekmann, Heesterbeek, and Britton (2013) for more background on the usefulness and the mathematical properties of ODE models for infectious disease modeling.

Many researchers have pointed to the link with survival analysis (Becker 1989; Becker and Britton 1999; Kenah 2011, 2013, 2015; KhudaBuksh et al. 2019), however, an accessible overview with standard software is lacking. The present paper has two purposes. The first is to explain the link between infectious disease models, in particular the susceptible-infectious-recovered (SIR) model, and methods from survival analysis in a concise and self-contained way. The second is to illustrate how standard software for survival analysis in the statistical language R can be used to estimate pivotal parameters in the infectious disease model.

We concentrate on the very much idealized situation where the epidemic is completely observed, i.e., at each time point we have complete and correct information on the number of susceptible, infected and recovered individuals. The ideas in this manuscript are largely known; the purpose is to establish further connections with *standard* models used in survival analysis, like Cox’s proportional hazards model, the additive hazard model, and Poisson generalized linear models (GLM’s), and to illustrate using R code how standard software for survival analysis can be used for estimating pivotal quantities in the SIR model, specifically the transmission parameter. We also show how methods from survival analysis may be used to cover extensions incorporating interventions, age structure and heterogeneity.

## 2 SIR model

### 2.1 Deterministic SIR model

The SIR model is a system of differential equations, describing the evolution over time of an infectious disease. Defining 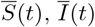 and 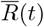 to be the proportion of susceptibles, infectious and recovered individuals in a closed population, the system of ordinary differential equations (ODE) is defined as (Anderson and May 1991)

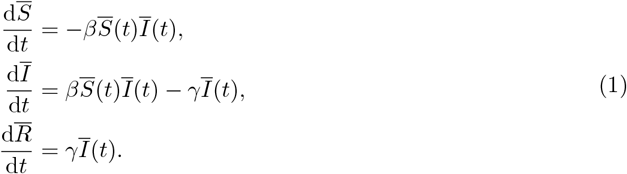

The idea behind Equation 1 is that interactions between susceptible and infectious individuals leading to a new infection occur with a rate quantified by an transmission parameter *β*. In case of an infection the proportion of susceptibles decreases and the proportion of infectious individuals increases by the same amount. Infectious individuals recover with a rate quantified by the recovery parameter *γ*. This class of models is often referred to as *compartimental models*, since it describes the interactions of units from different compartments. Apart from describing the spread of an infection through a population, compartimental models have been used in many other fields of application. Among the many examples we mention models describing the interaction of HIV and CD4+ T-cells within humans (Ho et al. 1995), predator-prey models in biology (Beddington, Free, and Lawton 1975) and economics (Murdoch, Briggs, and Nisbet 2013), and pharmacodynamics (Donnet and Samson 2013). Many extensions of this simple SIR model have been developed, with the aim of coming closer to a description of reality, but the simple SIR model has proved to be remarkably robust, and we will stick with the simple SIR model throughout.

We can numerically solve the ODE system using the {deSolve} package.

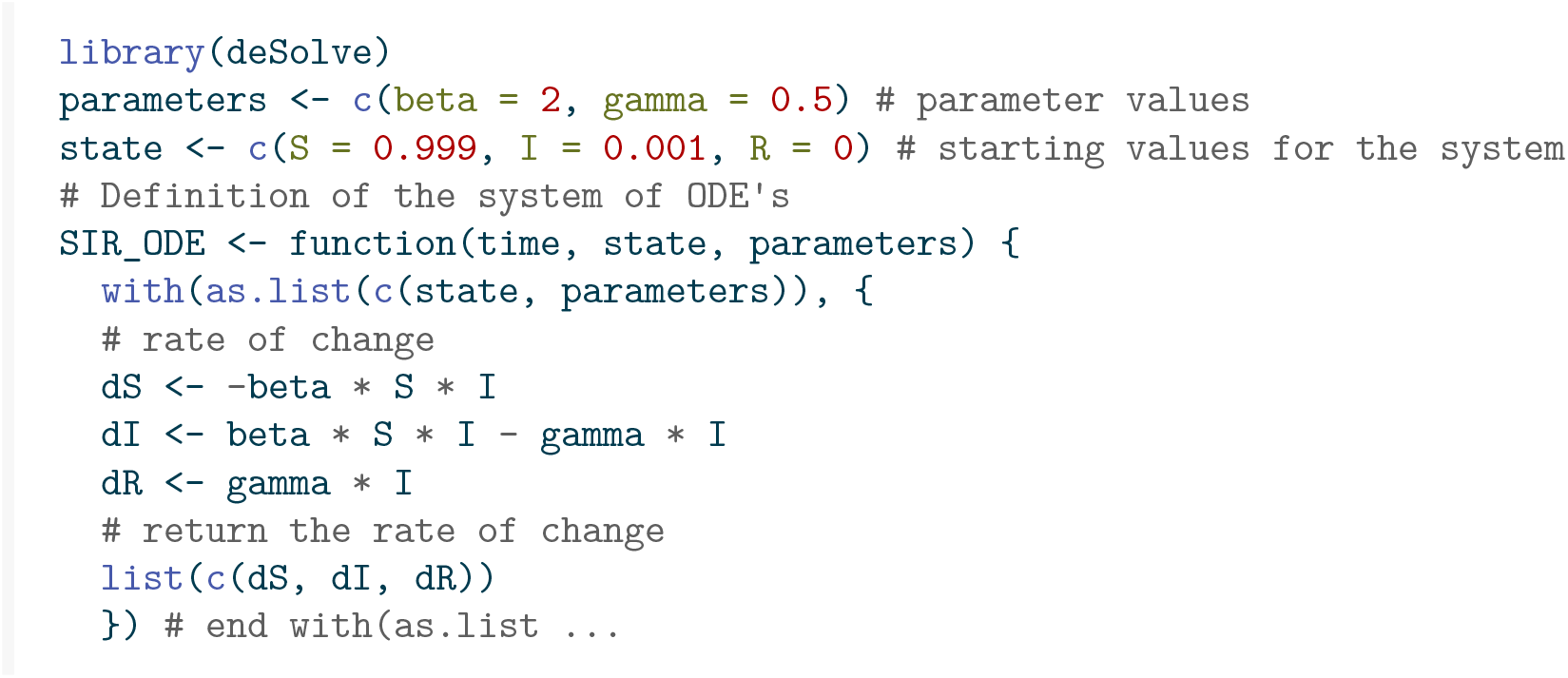
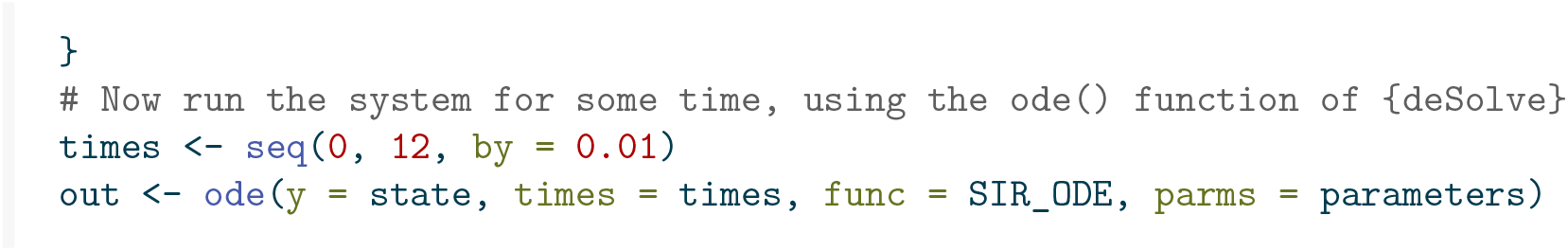

Figure 1 shows the development of the epidemic over time, in the form of a stacked plot. The lowest curve shows the proportion of infected individuals. The distance between the lowest curve and the one directly above that represents the proportion of recovered individuals. The sum of these two is the proportion of individuals that have got infected over time (recovered or not). The remainder is the proportion of susceptible individuals.

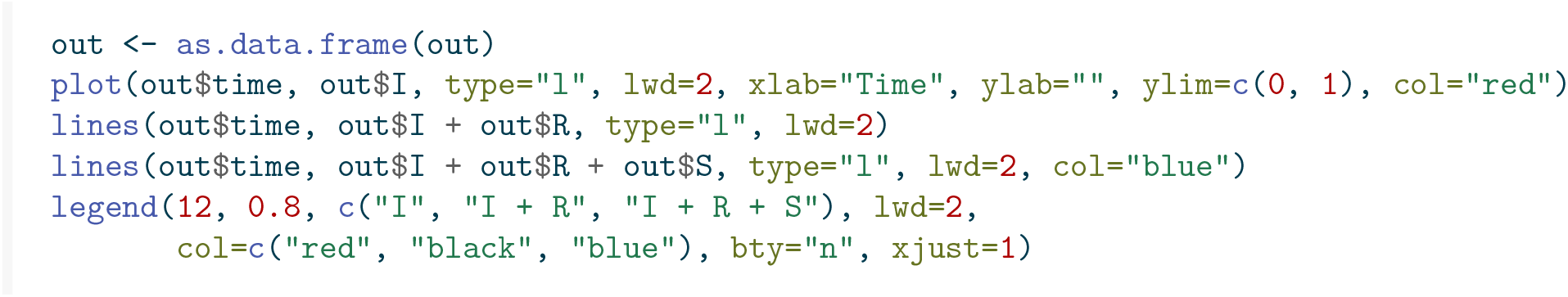

**Figure 1:**
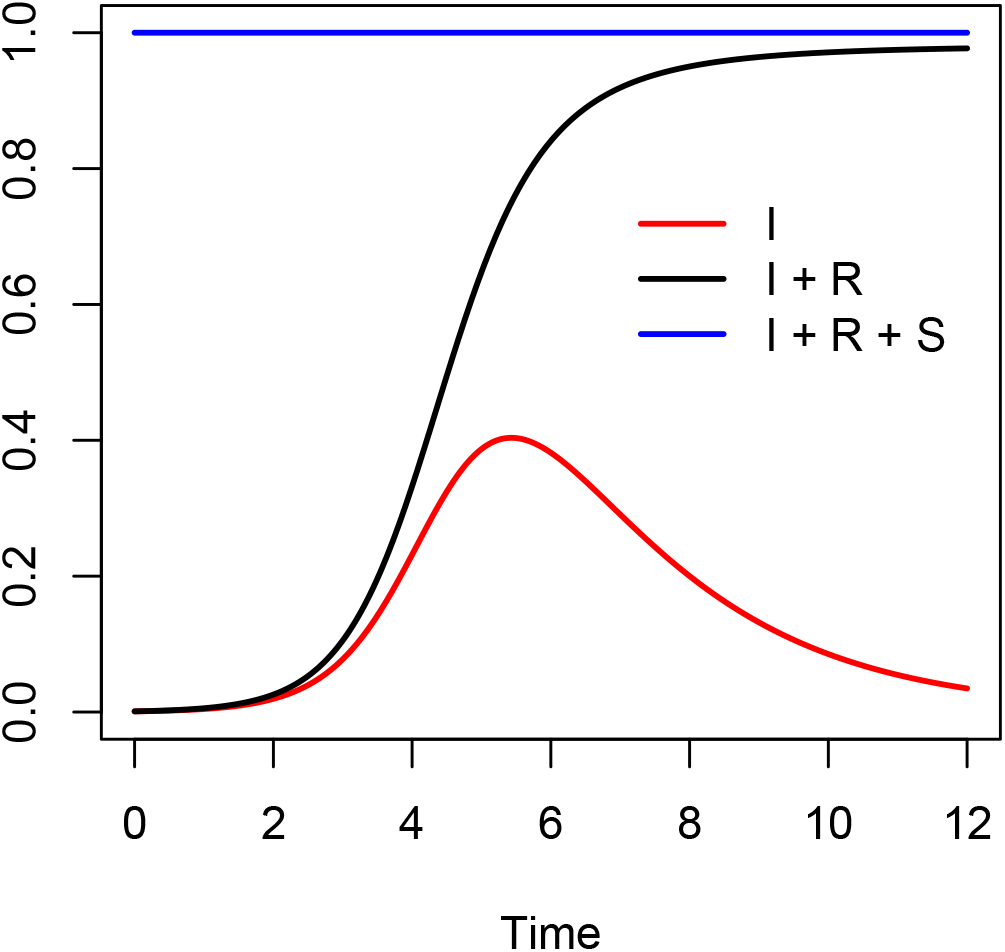
Stacked plot showing the proportion of susceptible, infected and recovered individuals over time in the SIR model; *β* = 2, *γ* = 0.5.

### Stochastic SIR model

The deterministic SIR model can be thought of as a “law of large numbers” limit of a stochastic system consisting of a large number of individuals, each of which has a rate of transitioning between states. The epidemic starts at time *t* = 0 with *Ī*(0), *S*(0) and *R*(0) individuals that are infected, susceptible and recovered, respectively, for a total of *n* = *S*(0) + *Ī*(0) + *R*(0) individuals. Because the system is closed, we in fact have *n = S*(*t*) *+ I*(*t*) *+ R*(*t*) for all t. We again use bars for the proportions 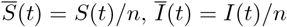 and 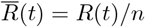 of susceptibles, infecteds and recovered individuals, respectively.

Becker and Britton (1999) discuss maximum likelihood estimation under complete observation. Let *N* (*t*) = *S*(0) − *S*(*t*) be the number of individuals infected in (0, *t*]. This is a counting process, as well as *R*(*t*), which is the number of individuals that recovered in (0, *t*], if *R*(0) = 0. If ℋ_*t*_ is the *σ*-algebra generated by the history {*S*(*u*), *Ī*(*u*); 0 ≤ *u < t*}, then the epidemic can be expressed in terms of the rates of the counting processes *N* (*t*) and *R*(*t*), by

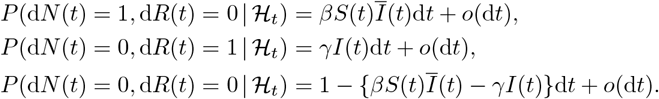

At some finite (random) time *τ* all infectious individuals will have recovered and the epidemic is over. Increasing *β* leads to higher infection rates, while lower *γ* leads to a longer time being infected and thus more opportunity to infect others. The ratio *β/γ* is known as the *basic reproduction number R*_0_, the average number of infections caused by one typical infectious individual in a completely susceptible environment, which in our example equals 4.

Code can be written to generate an epidemic outbreak, as a function of *β, γ* and the initial numbers of susceptible, infected and recovered (at *t* = 0).

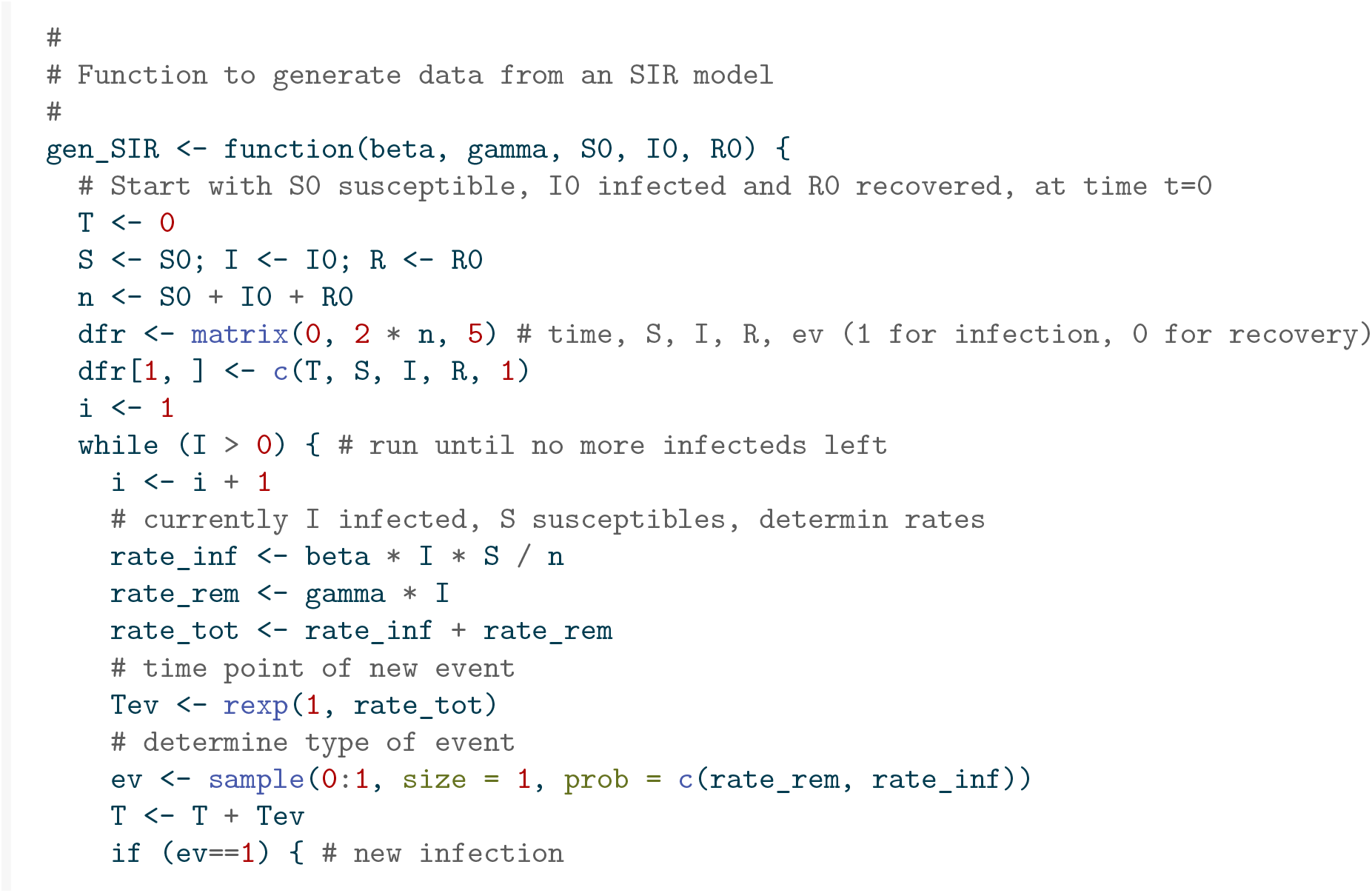
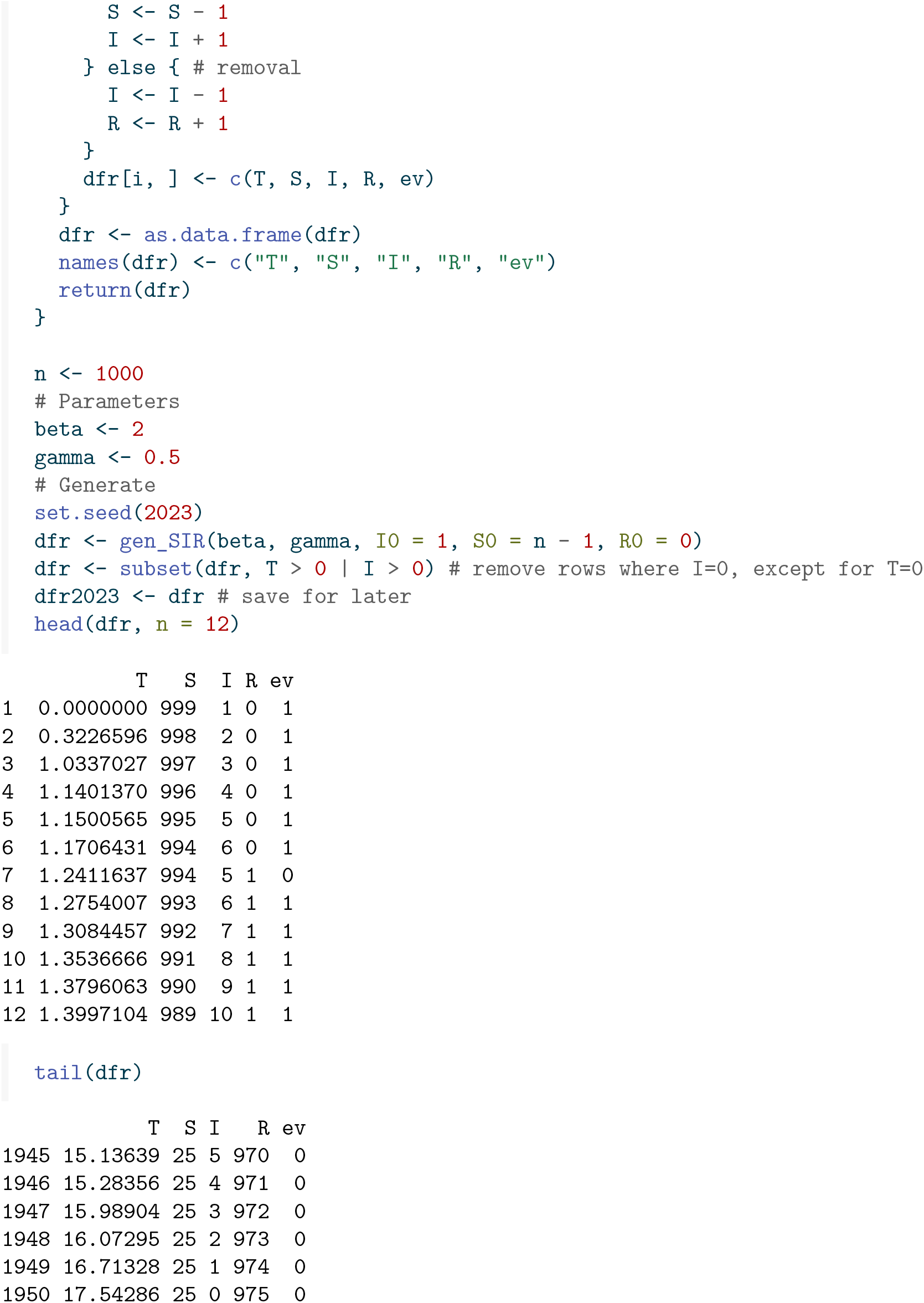

The data shows one realization of the schochastic process, with the same parameters as before, a population size of 1000, and a single infectious individual at *t* = 0, with (in column `T`) the time points at which events (either infections or recoveries) occur (column `ev` denotes whether it is an infection (`ev` =1) or a recovery (`ev` =0)). The columns `S`,`I` and`R` denote the number of susceptibles, infected and recovered in the population (total size = 1000). Note that 25 subjects have remained uninfected. Figure 2 shows the randomly generated epidemic.

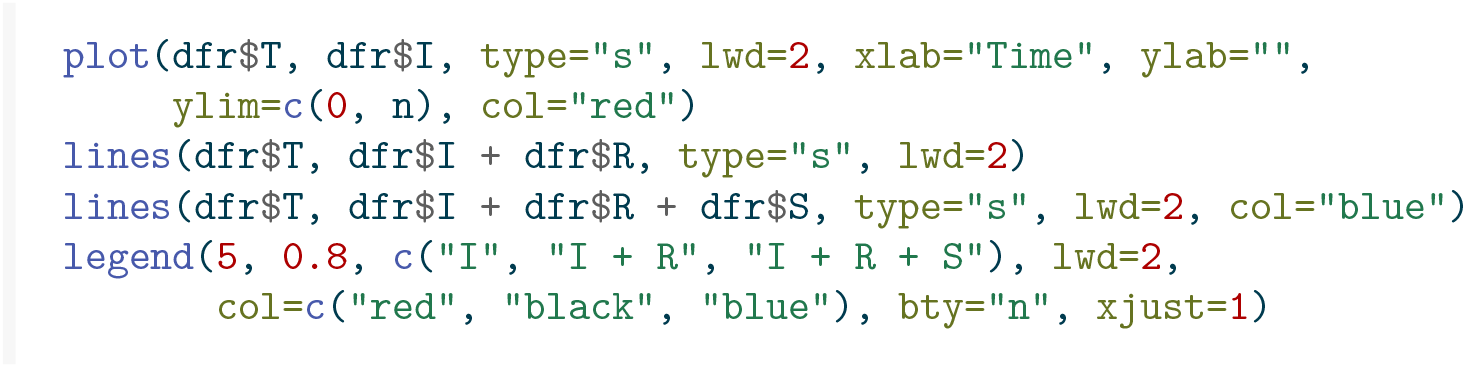

**Figure 2:**
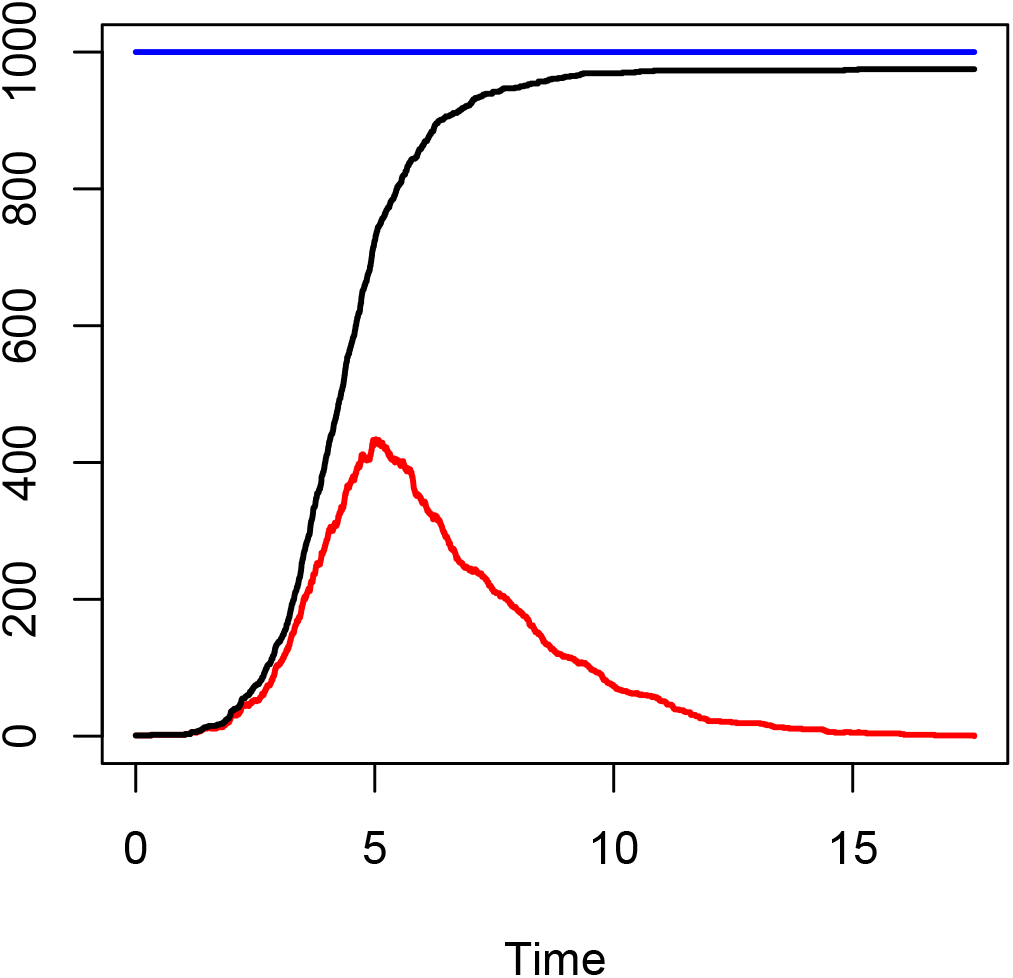
Stacked plot showing the number of susceptible, infected and recovered individuals over time from a stochastic SIR model.

This looks very much like the deterministic system, but with, on average, a slight delay in the time of peak prevalence. Note, however, that the system is now stochastic. With another seed we would obtain a different epidemic. Occasionally the epidemic might just not start off, because by chance the early infected individuals recover before they managed to infect new individuals. Figure 3 shows a histogram of (a) the peaks and (b) the time at which they occurred in 1000 simulated epidemics, with the same parameters as before, a population size of 1000 and one initial infected individual.

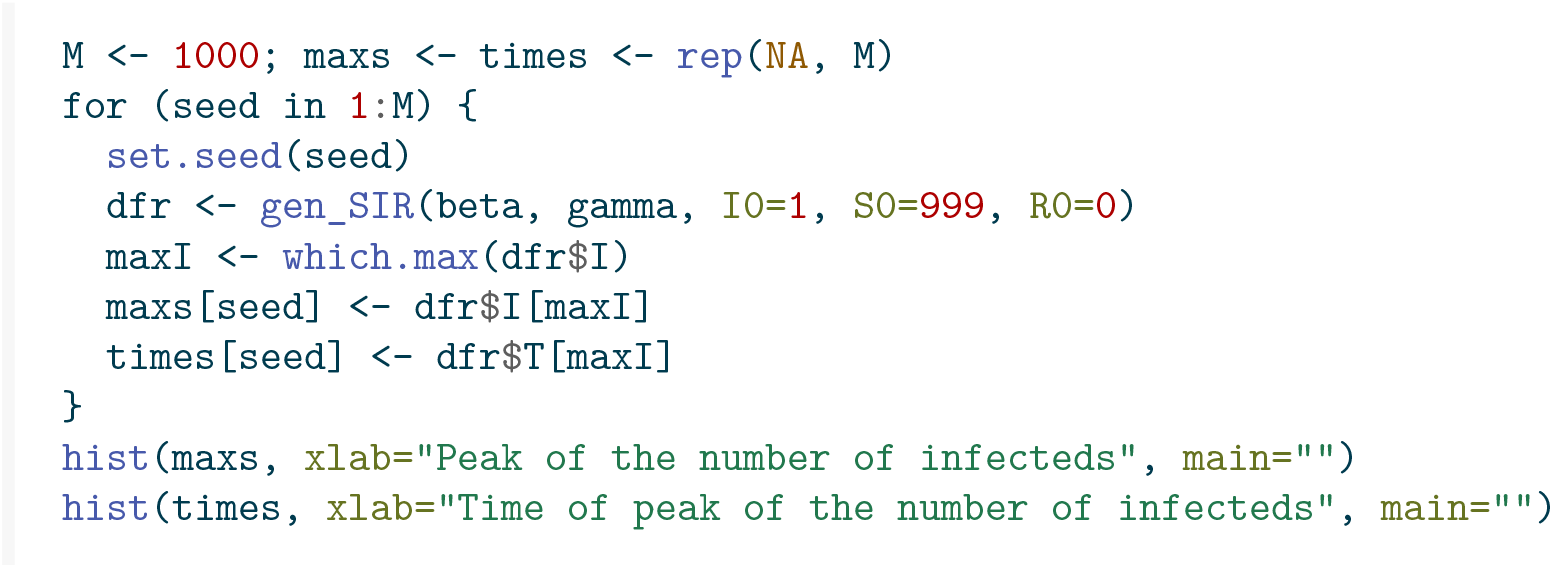

**Figure 3:**
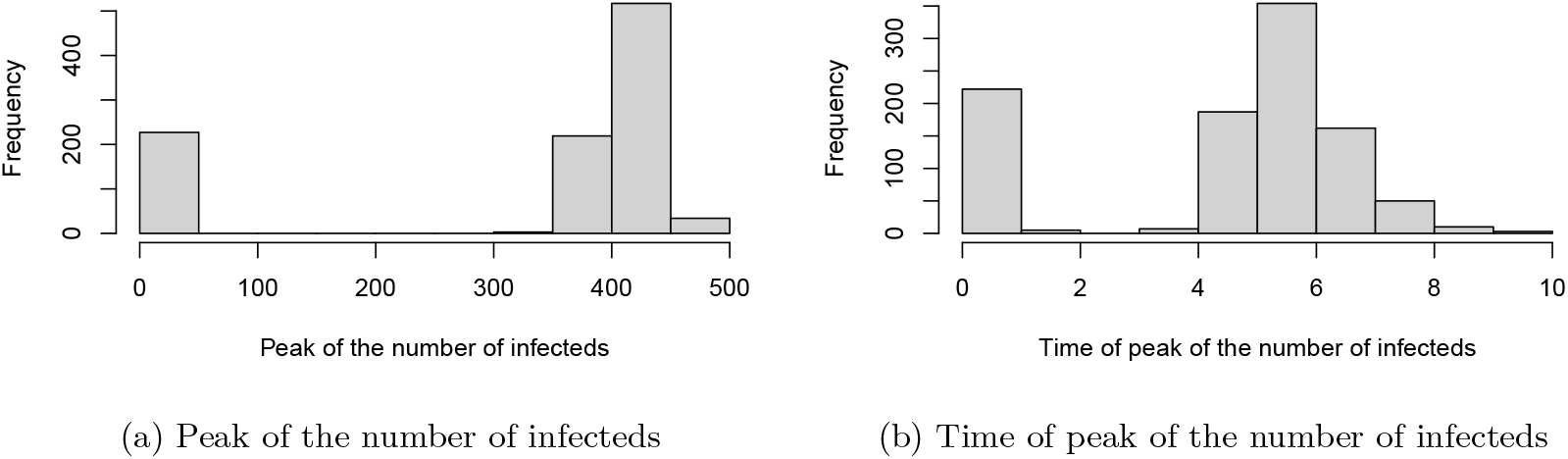
Results of 1000 simulated SIR epidemics

We see that in about 23% of the simulated epidemics, only a very limited number of individuals (here at most five, including the index case) got infected. This coincides with mathematical theory saying that with one initial infectious individual the probability of the epidemic not spreading to a large part of the population equals 1*/R*_0_ (Diekmann, Heesterbeek, and Britton 2013).

Becker and Britton (1999) derived the likelihood under complete observation, as

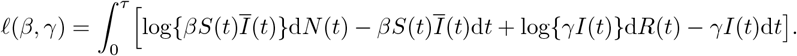

The first thing to notice is that the log-likelihood can be written as a sum of terms only depending on *β* and only on *γ*, respectively, implying that *β* and *γ* can be maximized separately. The maximum likelihood estimator (MLE) of *β* can then be analytically derived by taking the derivative of the part involving *β*, with respect to the parameter *β*, leading to (ignoring a *β*^−1^ term)

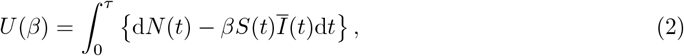

and setting this to zero. This yields

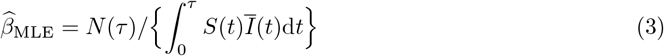

as the maximum likelihood estimator of *β*. Similarly (assuming *R*(0) = 0), we arrive at

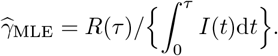

These estimates can be readily calculated from the current data.

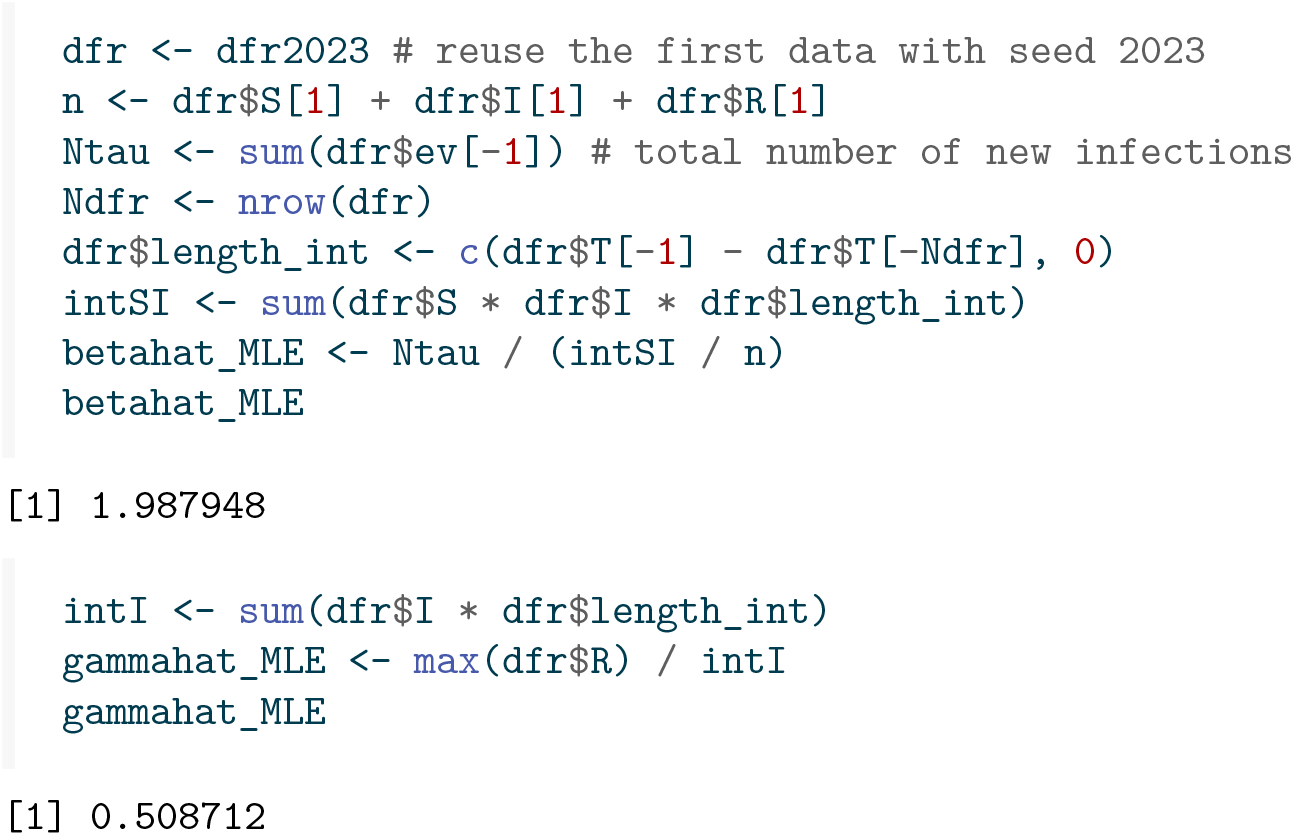

Estimates of the variances of 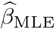 and 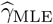 are also provided in Becker and Britton (1999). From now on we concentrate on the transmission parameter *β*. We see from Becker and Britton (1999) that 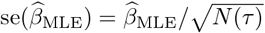. This gives the following value for the standard error of 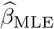:

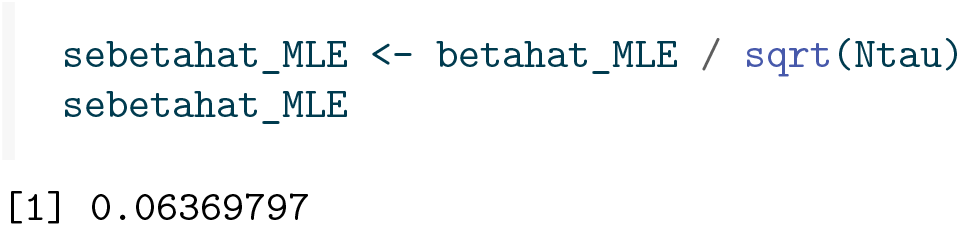

## 3 The link with survival analysis

Counting processes and their associated rates play a pivotal role in survival analysis so it is not surprising that methods from survival analysis can be used to estimate transmission parameters in SIR models. From now on, we restrict to estimation of *β* and ignore *γ*. The rates of the counting processes defined above are all aggregated over all susceptible and infected individuals. Restricting our attention to *N* (*t*), which is counting the total number of new infections occurring in (0, *t*], the link with survival analysis becomes clearer if we consider each of the individual counting processes *N*_*i*_(*t*) leading to 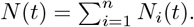. Thus, *N*_*i*_(*t*) counts the number of new infections *of individual Ī* occurring in (0, *t*], and this will be 1 or 0, depending on whether individual *i* has been infected or not before time *t*. Define *S*_*i*_(*t*), *Ī*_*i*_(*t*), and *R*_*i*_(*t*) to be random indicator variables taking the value 1 if at time *t* individual *i* is susceptible, infected or recovered, respectively, and 0 otherwise. The total number of susceptible, infected and recovered individuals is then 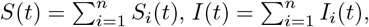, and 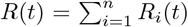, respectively. Individual *i* is only at risk of becoming infected while being susceptible. The usual notation in survival to indicate whether or not individual *i* is at risk at time *t* is *Y*_*i*_(*t*). Since the event of interest here is becoming infected, being at risk is the same as being susceptible, for which we have already defined the notation *S*_*i*_(*t*), so we have *Y*_*i*_(*t*) = *S*_*i*_(*t*). In the remainder of this paper we will be using *Y*_*i*_(*t*) when recalling general theory from survival analysis, and *S*_*i*_(*t*) when applying this to the SIR model. The rate of *N*_*i*_(t) is given by *λ*_*i*_(*t*) = *βĪ*(*t*), while being susceptible, which can then be written as *λ*_*i*_(*t*) = *βS*_*i*_(*t*)*Ī*(*t*). In this way, adding over all individuals, we see that the rate of *N* (*t*) equals 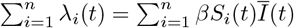, leading to a total rate of *βS*(*t*)*Ī*(*t*), the same as in Equation 2. Also, each *infected* individual has a constant rate *γ* of recovering.

This perspective is called *individual-based* or *agent-based* modeling, see KhudaBuksh et al. (2019). With the purpose of estimating *β*, we can exploit the link to survival analysis, again under complete observation, by translating the original data, documenting the number of S, I, R individuals over time, into a long format data set where each susceptible subject occurs individually, and where at the same time the number of infected individuals is kept track of, using start-stop notation, also called Andersen-Gill (Andersen and Gill 1982) or counting process notation. The following code establishes this, starting from the original `dfr` data.

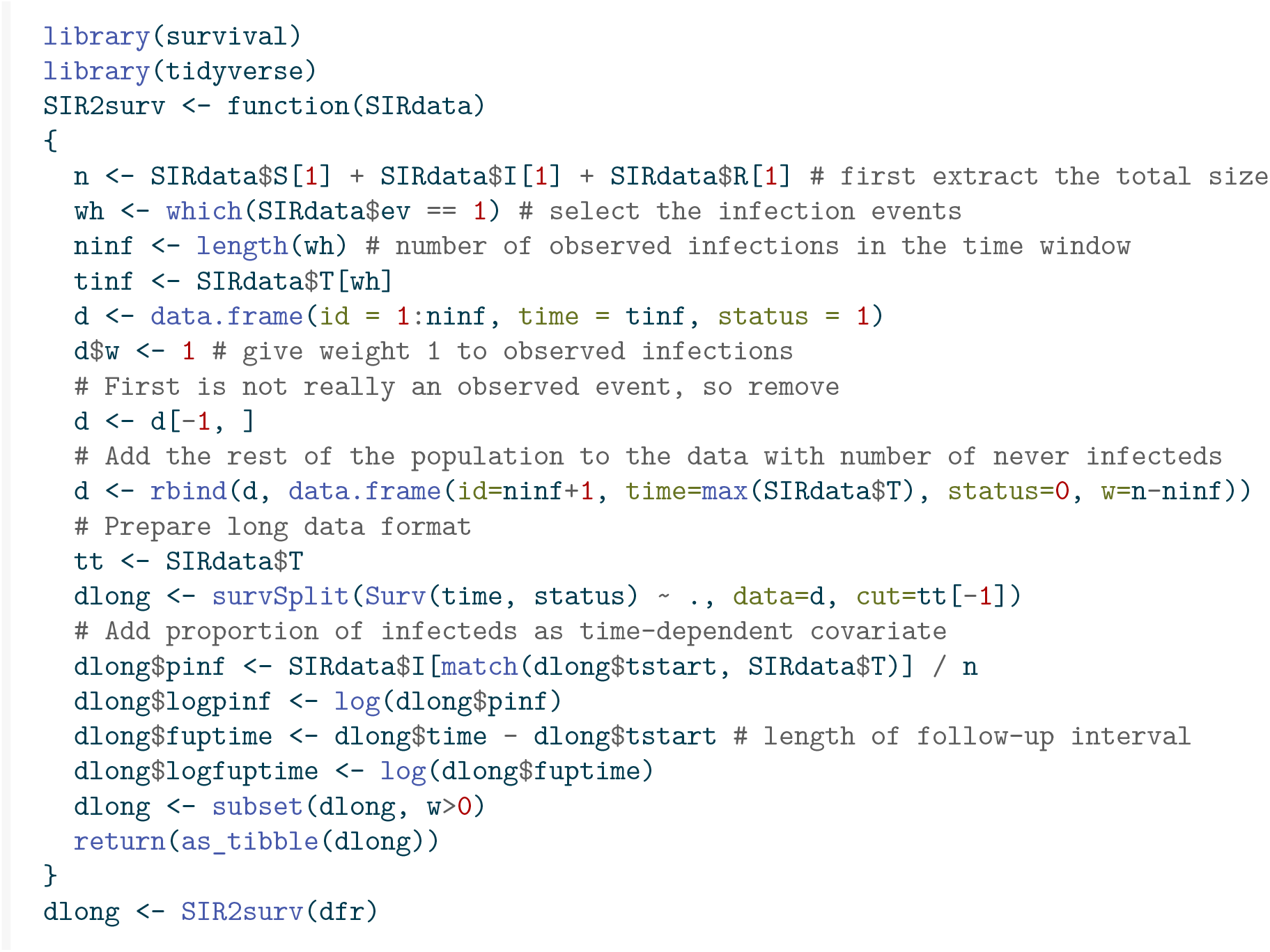

The head and tail of this expanded data set looks like this. The column `fuptime` has the length of the time interval between `tstart` and time, and its logarithm, `logfuptime`, will prove to be useful for Poisson regression later.

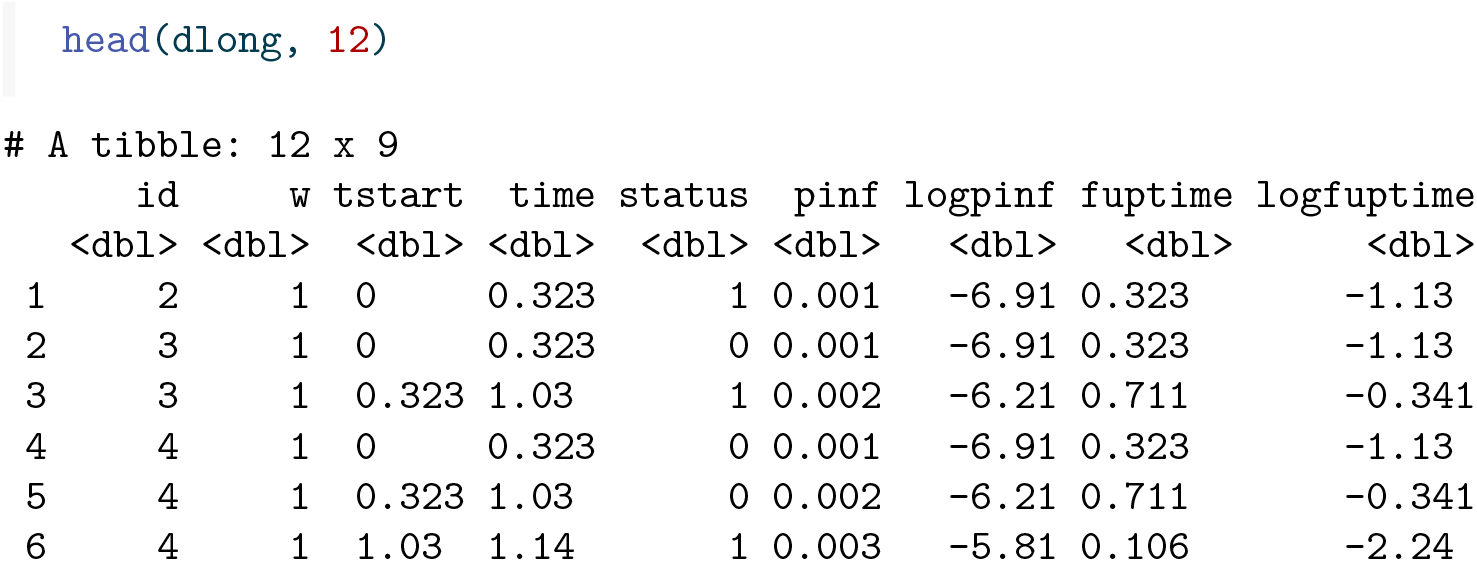
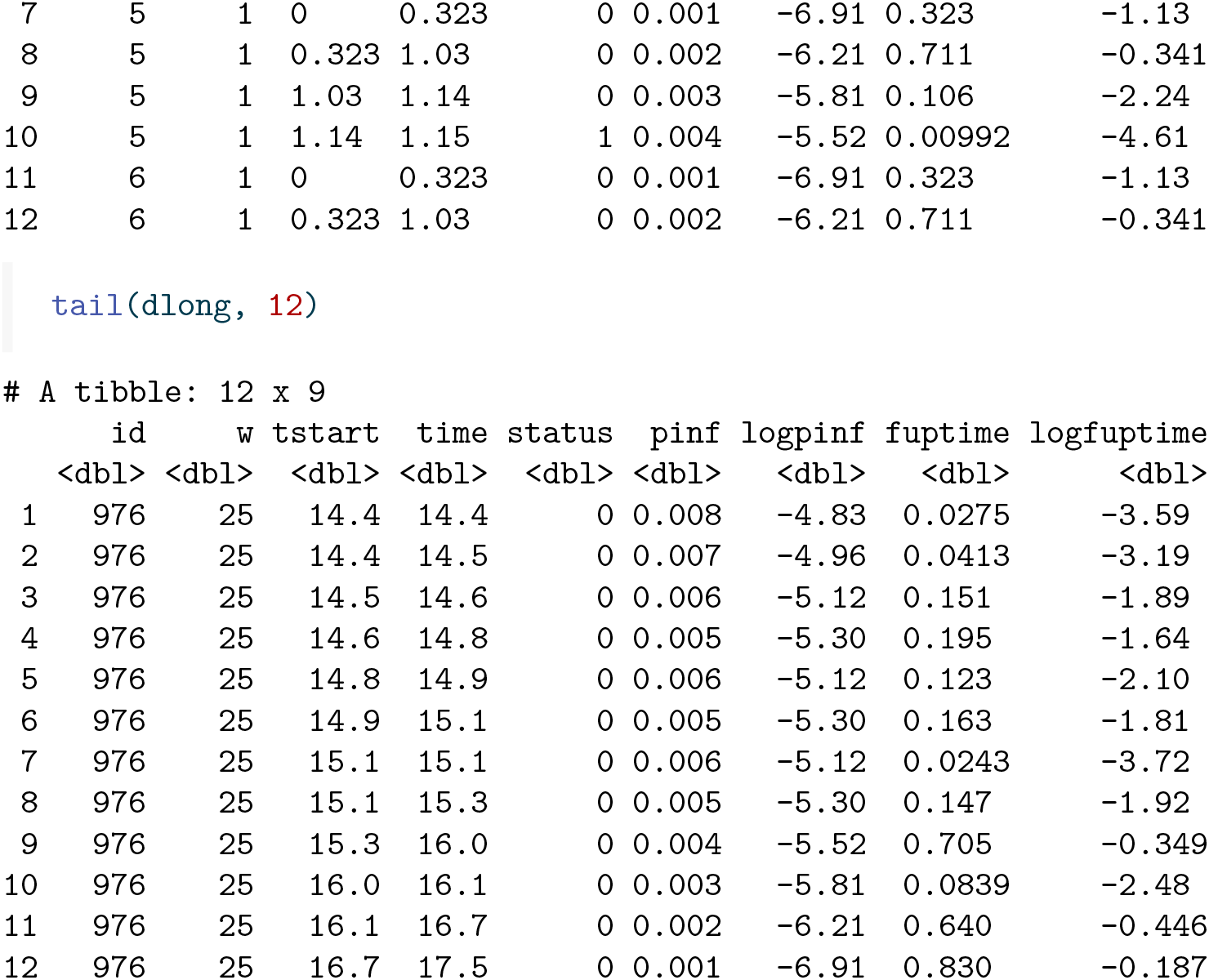

A more concise version of this long format data can be obtained by combining all the rows with the same time interval (`tstart, time`), and adding all the weights, separately for `status=0` and`status=1` .

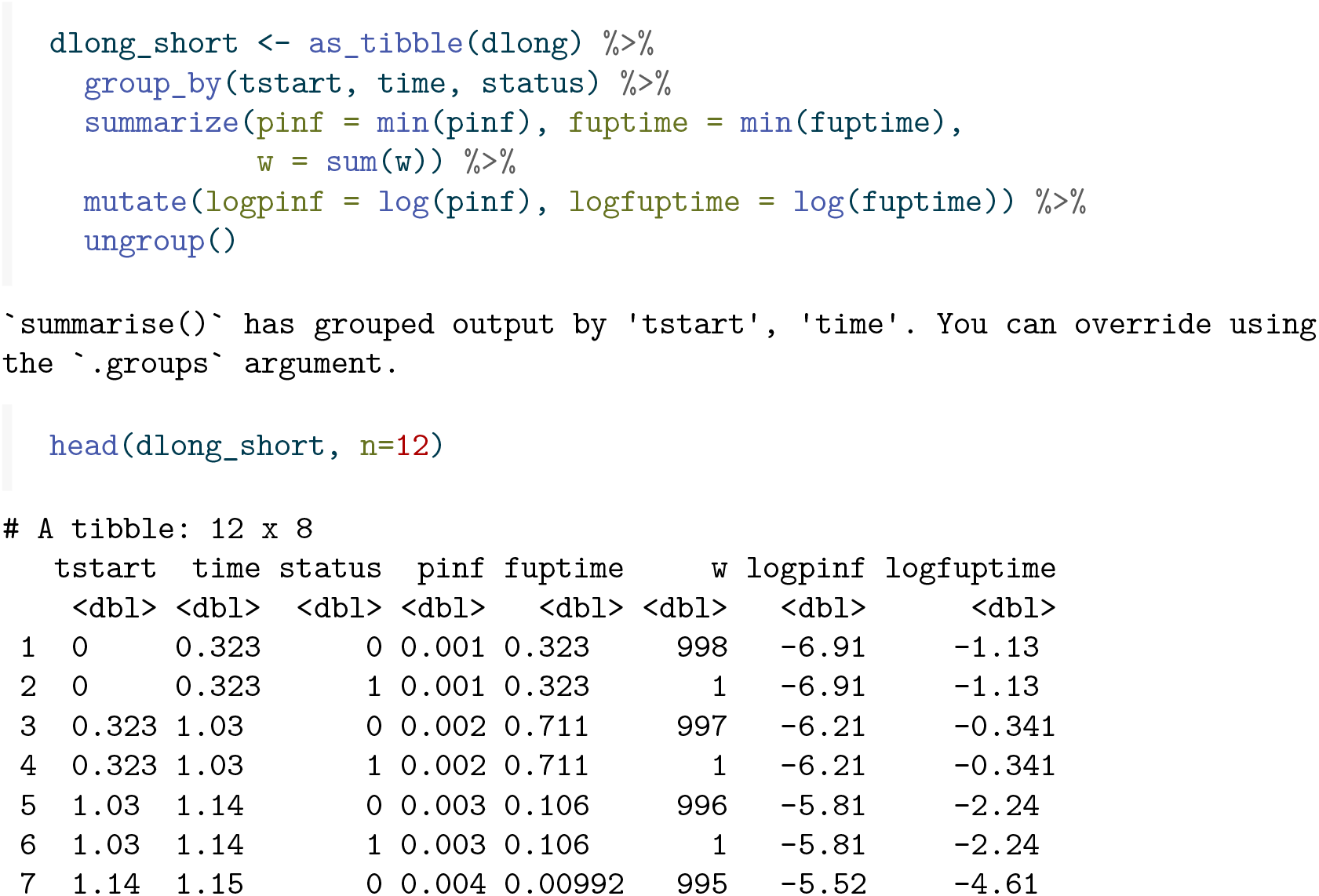
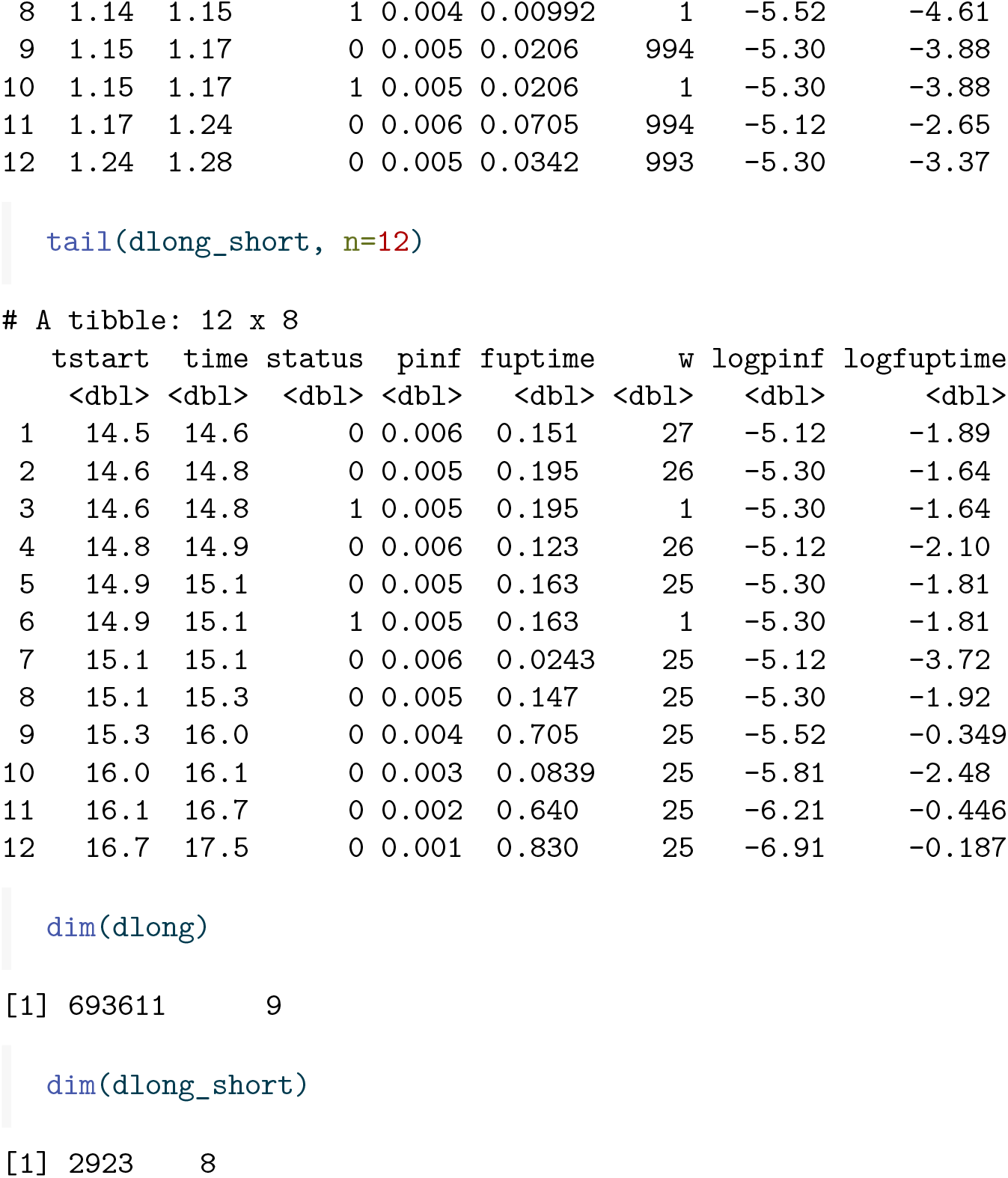

In the remainder of this section we will review a number of standard models in survival analysis, show how they apply to the SIR model, and illustrate how standard software for survival analysis can be used, in combination with the long format data, to fit these models.

### 3.1 Additive hazards models

The additive hazards model (Aalen 1980, 1989) specifies the rate of new infections as a sum of (typically time-dependent) linear combinations of the covariates, which themselves may also be time-dependent:

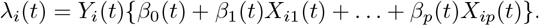

The first term within brackets, *β*_0_(*t*), is an intercept, comparable to the baseline hazard in the Cox model, *X*_*i*1_(*t*), …, *X*_*ip*_(*t*) is a set of possibly time-dependent covariates, and *β*_1_(*t*),, *β*_*p*_(*t*) are the regression coefficients. Typically these are taken to be time-dependent, but we will also consider the case later where the *β*(*t*)’s are constant over time. Aalen’s ordinary least squares (OLS) estimates focus on the cumulative regression functions 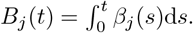.

Defining vectors **N**(*t*) = (*N*_1_(*t*), …, *N*_*n*_(*t*))^⊤^, **B**(*t*) = (*B*_0_(*t*), *B*_1_(*t*), …, *B*_*p*_(*t*))^⊤^, and the matrix **X**(*t*), with *i*th row (*Y*_*i*_(*t*), *Y*_*i*_(*t*)*X*_*i*1_(*t*), …, *Y*_*i*_(*t*)*X*_*ip*_(*t*)), then Aalen, Borgan, and Gjessing (2008) derive

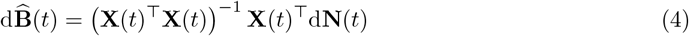

as estimate of the increment of **B**(*t*), provided **X**(*t*) has full rank. In the absence of the intercept term *β*_0_(*t*) in the model, the constant elements in the first column of **X**(*t*) are removed.

Applying this model to the SIR setting, since the rate of each susceptible individual equals *βĪ*(*t*), we can view this as a term *β/n* being added to the hazard for each infectious individual. So we would have no intercept, *p* = 1, and *X*_*i*1_(*t*) = *X*_*i*_*(t*) = *Ī*(*t*) for each individual, and **X**(*t*) would be a *N ×* 1 vector with *i*th element *Y*_*i*_(*t*)*X*_*i*_(*t*) = *S*_*i*_(*t*)*Ī*(*t*). Using Equation 4 leads to

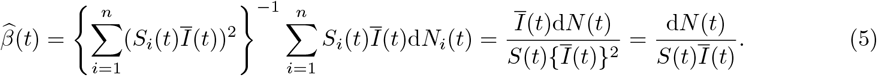

Here we have used that 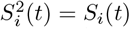 and *S*_*i*_(*t*)d*N*_*i*_(*t*) = d*N*_*i*_(*t*) (because an event can only happen when someone is at risk). This 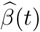can be estimated in the {timereg} package in R, by fitting an additive hazards model with *Ī*(*t*) (in column `pinf`) as covariate, *excluding* an intercept (hence `pinf - 1` in the formula below). This idea has been used before in Wolkewitz et al. (2002). The plot in Figure 4 shows an estimate of 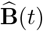 over time.

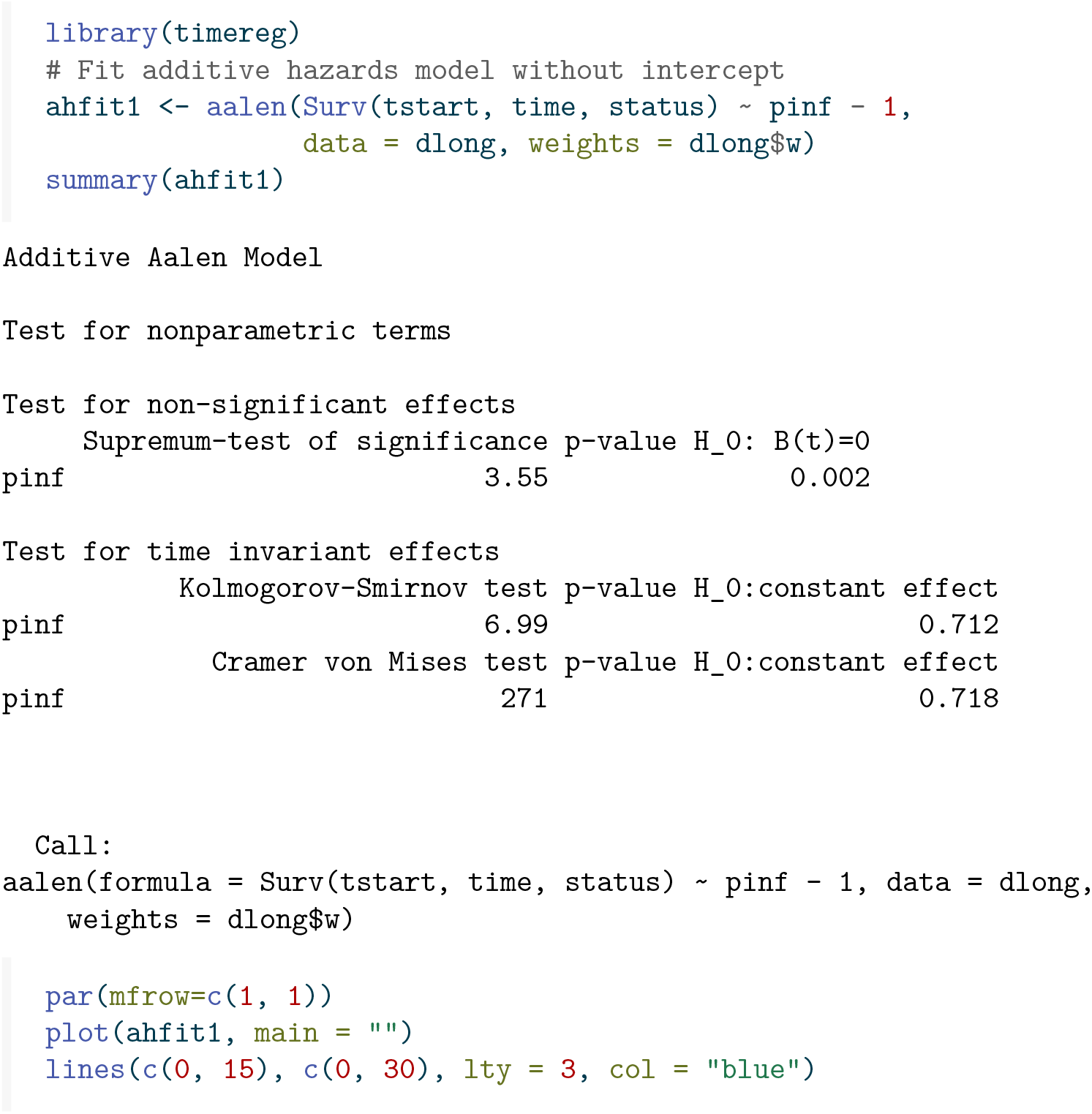

**Figure 4:**
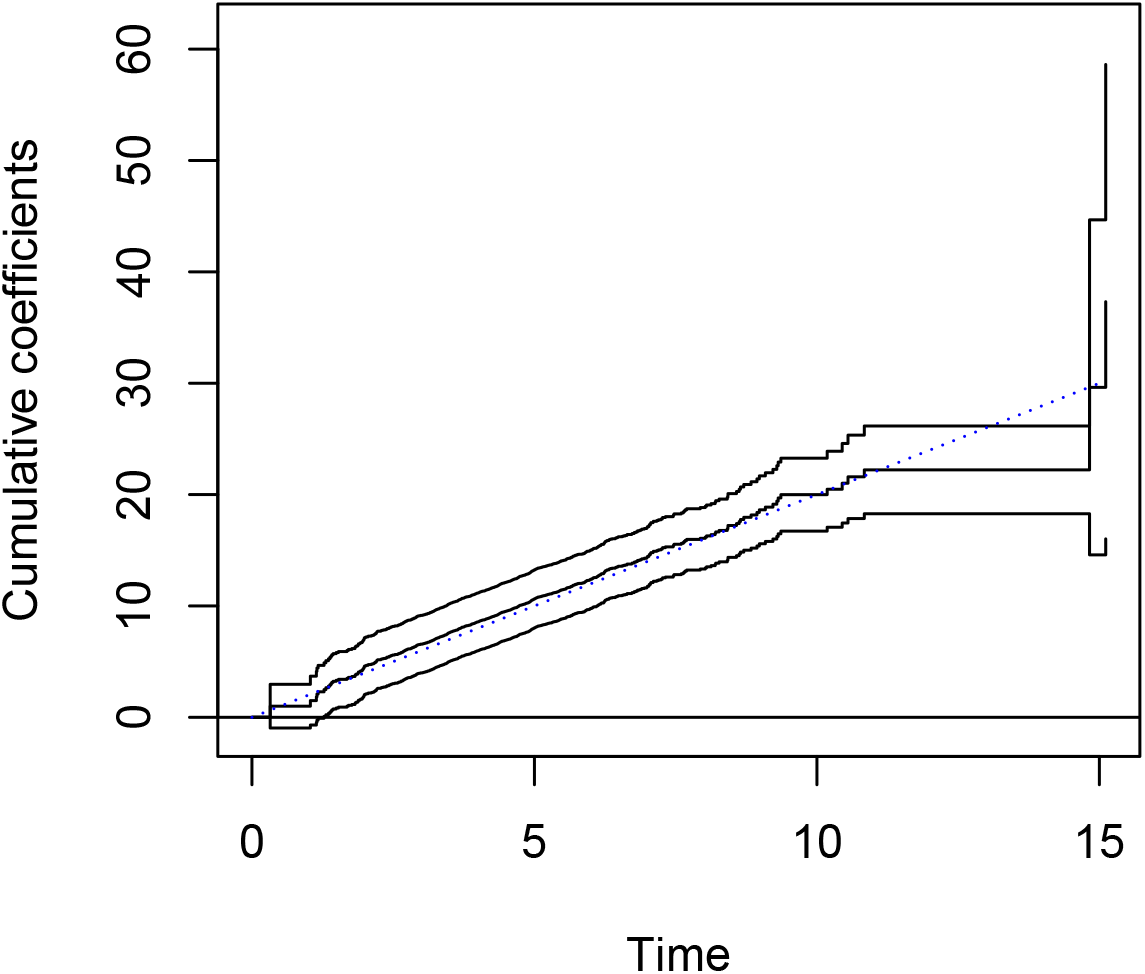
Estimate of *B*(*t*) over time; the dotted line represents the true *B*(*t*).

It is reassuring that the estimate of *B*(*t*) indeed seems to follow a straight line, indicating constant *β*(*t*). The hypothesis of *β*(*t*) being constant is actually added in the “tests for time invariant effects”, of which both the Kolmogorov - Smirnov and the Cramer - von Mises tests show no significant departures from time invariant effects. For background on these tests we refer to Martinussen and Scheike (2006). The slope is approximately equal to *β* = 2, as we would hope, see the dotted line in Figure 4.

Inspired by least squares theory, a least squares estimator would be given by a variant of the displayed equation between (2.6) and (2.7) of Lin and Ying (1994), the difference being the absence of the baseline hazard term, given by

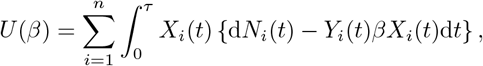

where we recall that the time-dependent covariate is given by *X*_*i*_(*t*) = *Ī*(*t*), and *Y*_*i*_(*t*) is the at risk indicator, which in our case was earlier denoted by *S*_*i*_(*t*). We will first elaborate on the general theory, then replace *X*_*i*_(*t*) by *Ī*(*t*) and *Y*_*i*_(*t*) by *S*_*i*_(*t*) to apply it to the SIR case. Setting *U* (*β*) to zero and solving for *β* yields

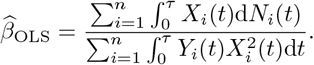

Following Lin and Ying (1994), define

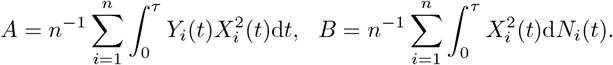

Then the variance of 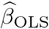 can be consistently estimated by *B/*(*nA*^2^), which equals

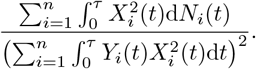

We go back to the formulas for 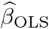 and its variance, and now replace *X*_*i*_(*t*) by *Ī*(*t*) and *Y*_*i*_(*t*) by *S*_*i*_(*t*), leading to

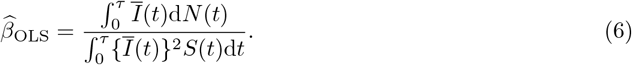

Note first that 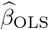 is of a similar form as the estimate 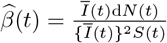 sof Equation 5, but with both the numerator and denominator being integrated over time. Note also that 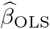 is similar to 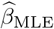 in Equation 3, except that 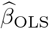 has an extra weighting term *Ī*(*t*) in both numerator and denominator.

The estimate of its variance is given by

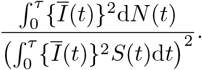

Here is the result when implementing these formulas in our data, given the following estimate:

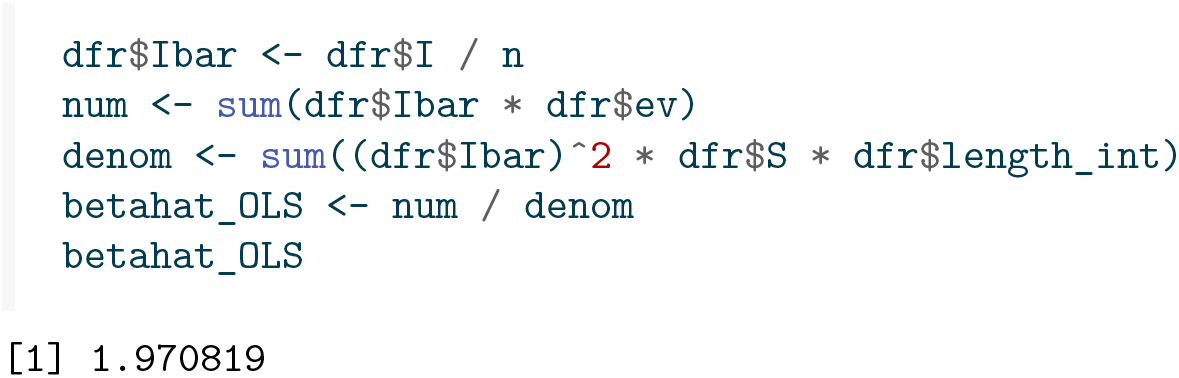

and its estimated variance:

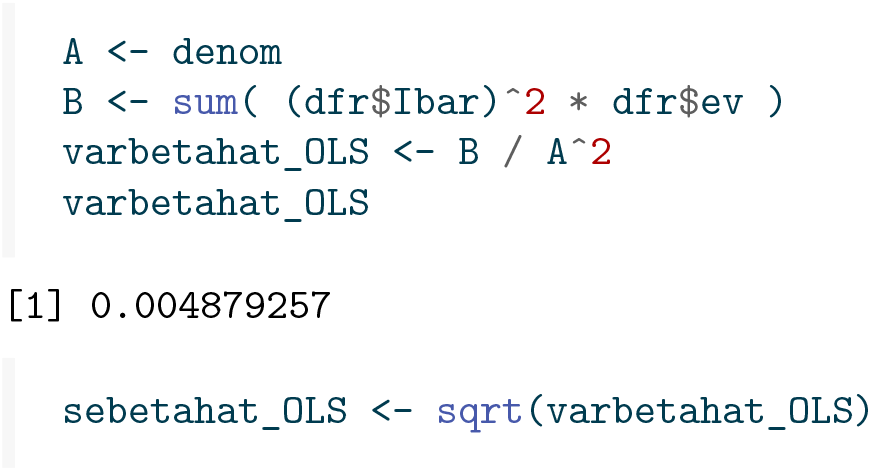

The standard error equals 0.070, which is about 10% larger than the one obtained by Becker and Britton (1999), which was 0.064. Summarizing, the estimate and its 95% confidence interval are given by

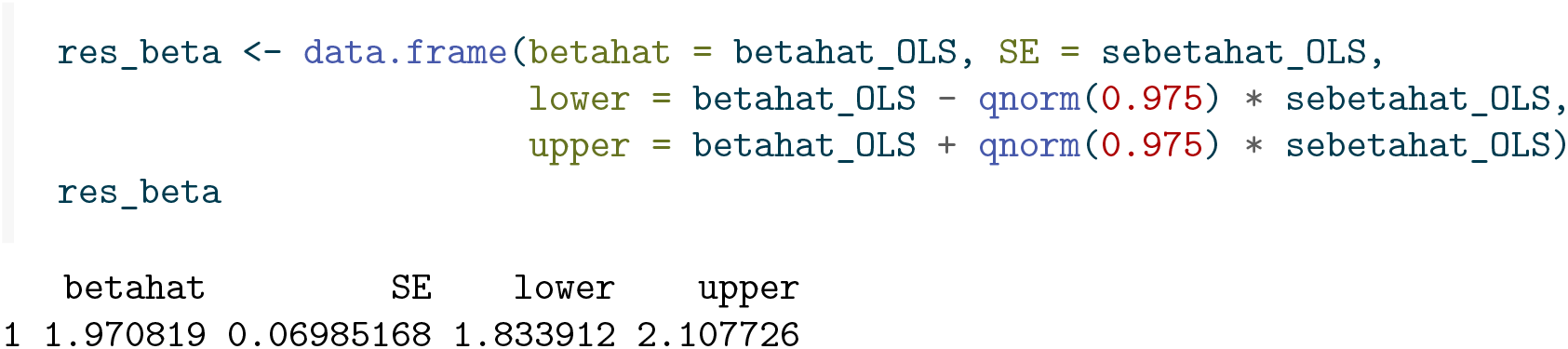

The {timereg} package can in principle also estimate constant *β*(*t*), but somehow not without an intercept, so as far as I have been able to see the above procedure can not be implemented in {timereg}.

### 3.2 Multiplicative models

#### 3.2.1 Cox models

Recall that the rate of *N*_*i*_(*t*) equals *βĪ*(*t*) while being at risk. The Cox model assumes that the rate of the event equals *Y*_*i*_(*t*)*h*_0_(*t*) exp(*β*_1_*X*_*i*1_(*t*) + … + *β*_*p*_*X*_*ip*_(*t*)) = *Y*_*i*_(*t*)*h*_0_(*t*) exp(*β*^⊤^*X*_*i*_(*t*)). Here *Y*_*i*_(*t*) again is the at risk indicator, *h*_0_(*t*) is an unspecified baseline hazard, *X*_*i*1_(*t*), …, *X*_*ip*_(*t*) are possibly time-dependent covariates, and *β*_1_, …, *β*_*p*_ are regression coefficients to be estimated. In the absence of ties, the vector of regression coefficients is estimated by maximizing the *partial likelihood*, given by

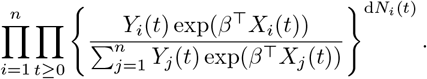

This partial likelihood is maximized with respect to *β* to obtain estimates 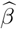 of *β*. For given *β*, in the absence of ties the estimate of the baseline hazard increment is given by Breslow’s estimate

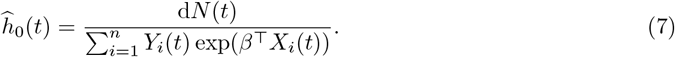

If we take *p* = 1, *X*_*i*1_(*t*) = log(*Ī*(*t*)), and fix *β*_1_ = 1, we get a hazard rate of *h*_0_(*t*) exp(log(*Ī*(*t*))) = *h*_0_(*t*)*Ī*(*t*). Using Equation 7 we get

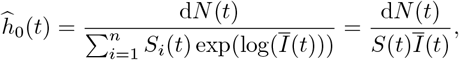

which can be seen to equal the hazard increments 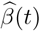 from Equation 5 obtained from the additive hazards model!

If we fit a Cox model with log(*Ī*(*t*)) (`logpinf`) as an *offset* (meaning we are not estimating the associated regression coefficient, but setting it to one), then the baseline hazard rate of the Cox model becomes *h*_0_ (*t*) exp(log(*Ī*(*t*))) = *h* (*t*)*Ī*(*t*), the estimated baseline rate 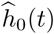 should equal 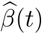, and the cumulative baseline hazard 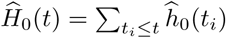 should resemble *βt*, in other words a straight line with intercept 0 and slope *β*. Fitting the Cox regression seems to recover the constant rate, although care is needed to extract the baseline hazard due to the offset term. When adding the estimated cumulative hazard of Aalen’s additive hazard model to the resulting estimate, we indeed see that the Cox and Aalen based baseline hazards are exactly the same.

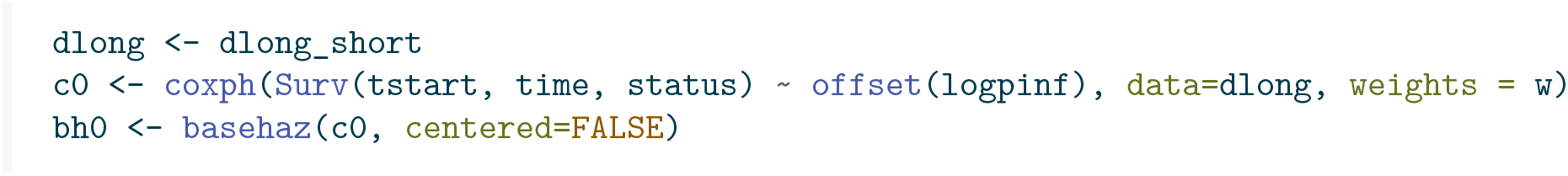
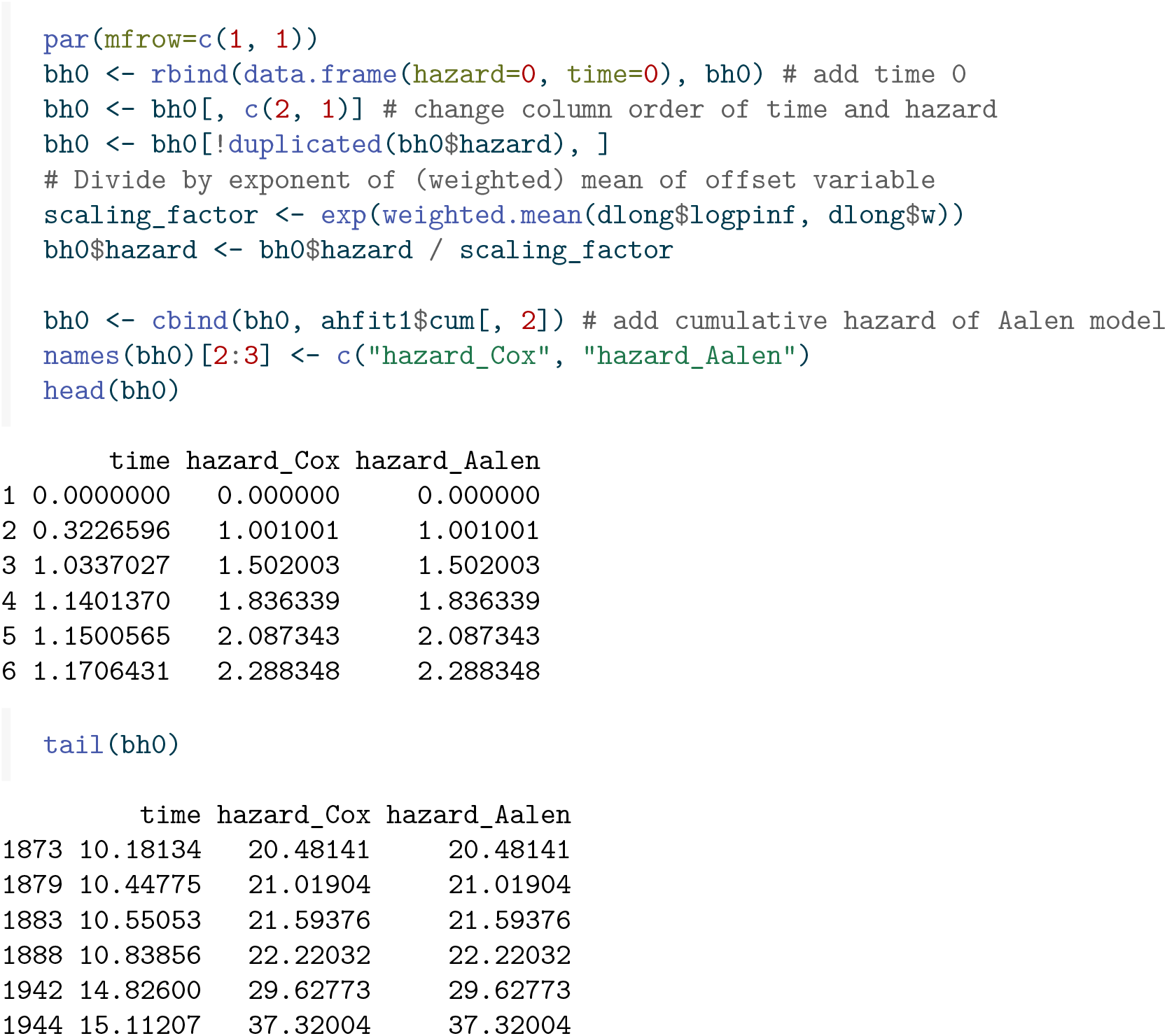

#### 3.2.2 Poisson regression

The Cox model uses an unspecified baseline hazard and does not use the assumption that the infection rate is constant. We could attempt to estimate the infection rate, assuming that it is constant, based on the well established epidemiologic occurrence over exposure (O/E), see Clayton and Hills (1993). The variance of the log rate is the inverse of the number of events, by which we could construct a 95% confidence interval of the estimated rate.

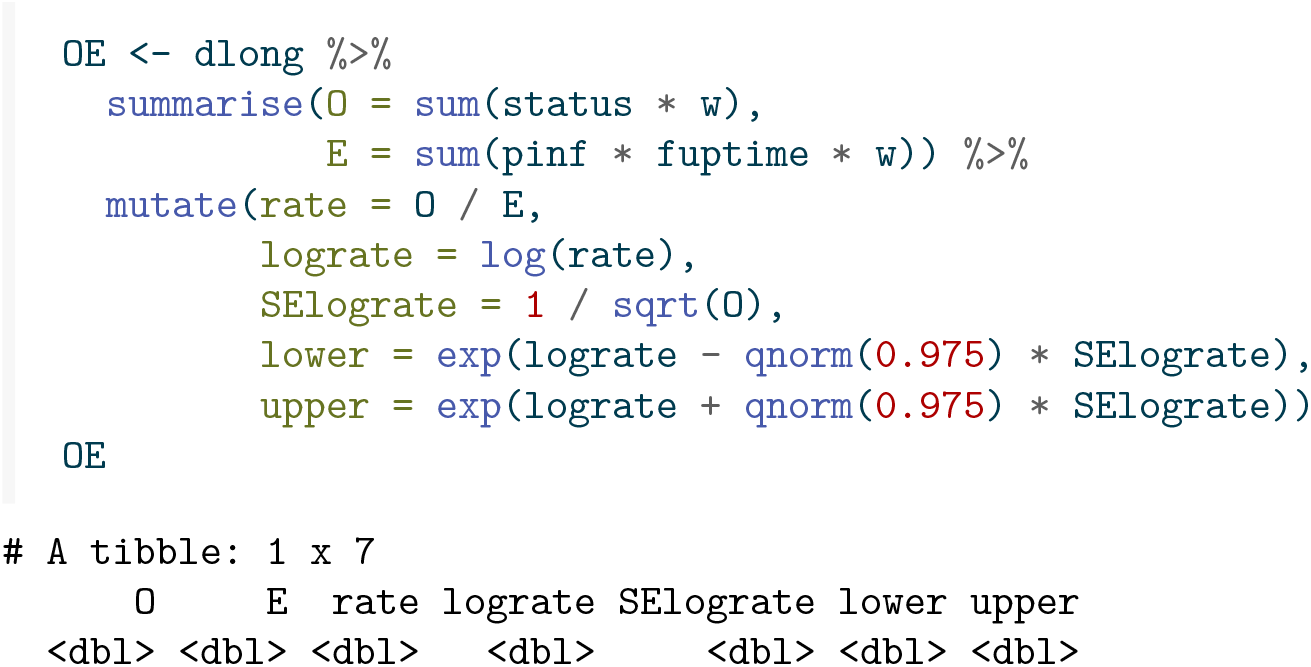
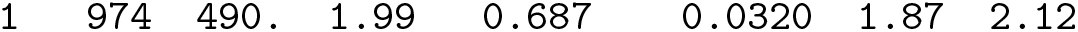

This quite nicely returns the true rate 2. It is also identical to the value of 1.9879483 for *β* obtained by the direct formula of Becker and Britton (1999).

Alternatively, Poisson regression can be used. The expected number of infections in a short interval of length Δ*t* around *t* equals *βĪ*(*t*)Δ*t*, so with a log link this equals exp(log *β* + log *Ī*(*t*) + log Δ*t*). This means that fitting a GLM Poisson model with log link, and with both log *Ī*(*t*) and log Δ*t* as offset terms, we obtain an estimate of log *β*. Taking the exponent of the estimate of log *β* then provides an estimate of *β*.

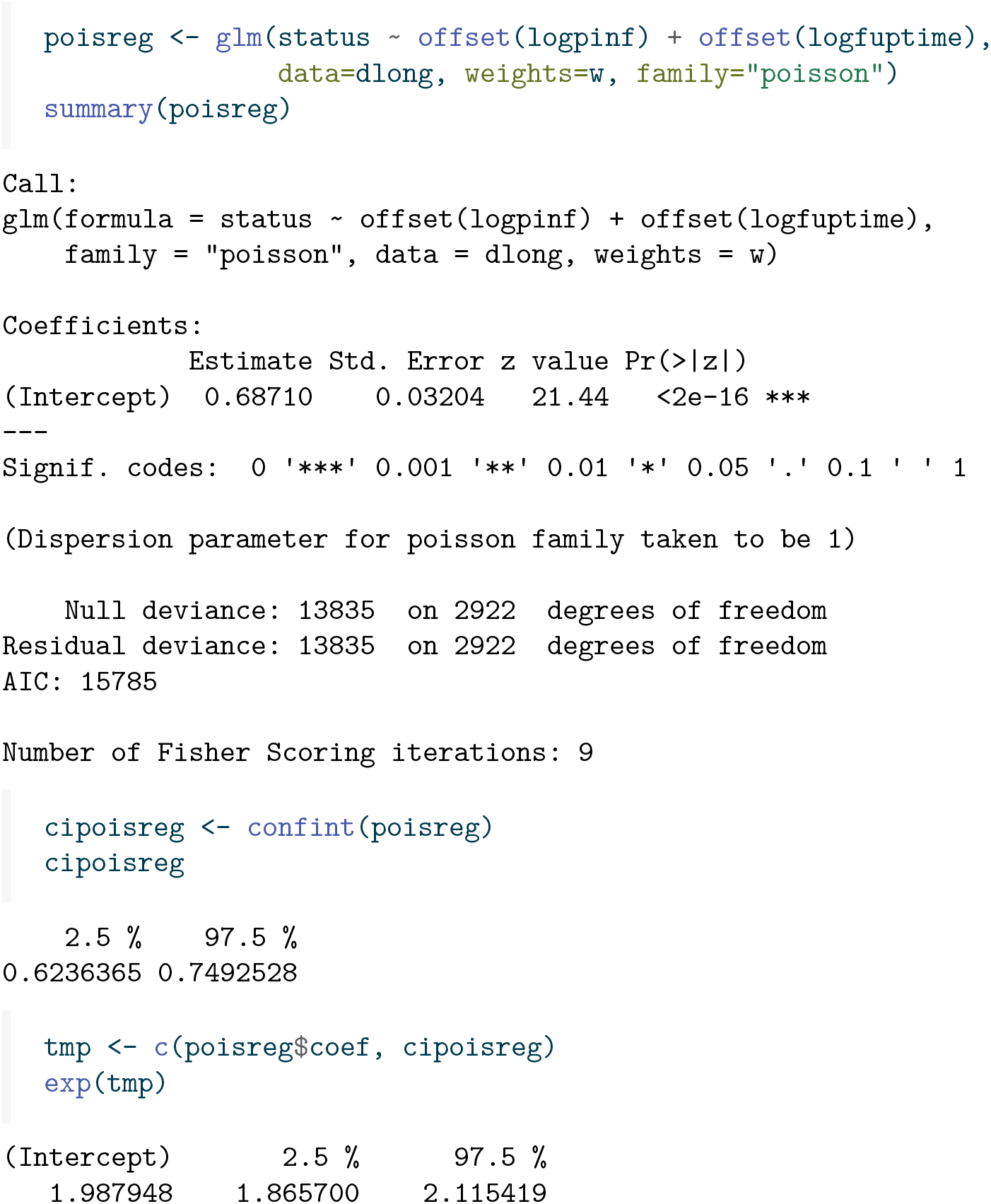

This again gives exactly the same result for 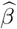 as Becker and Britton (1999). The 95% confidence intervals given by the occurrence/exposure formulas and Poisson regression differ slightly, but this is due to different ways of constructing these confidence intervals (Wald versus likelihood-based). The standard errors from the Poisson regression and the occurrence / exposure formulas *are* exactly the same.

The Poisson regression model also gives the opportunity to estimate *β*(*t*) as a smooth function of time using splines. We use the natural splines (cubic splines that are linear beyond the outermost knots), as implemented in the `Ns()` function in the {Epi} package, but other splines can also be used.

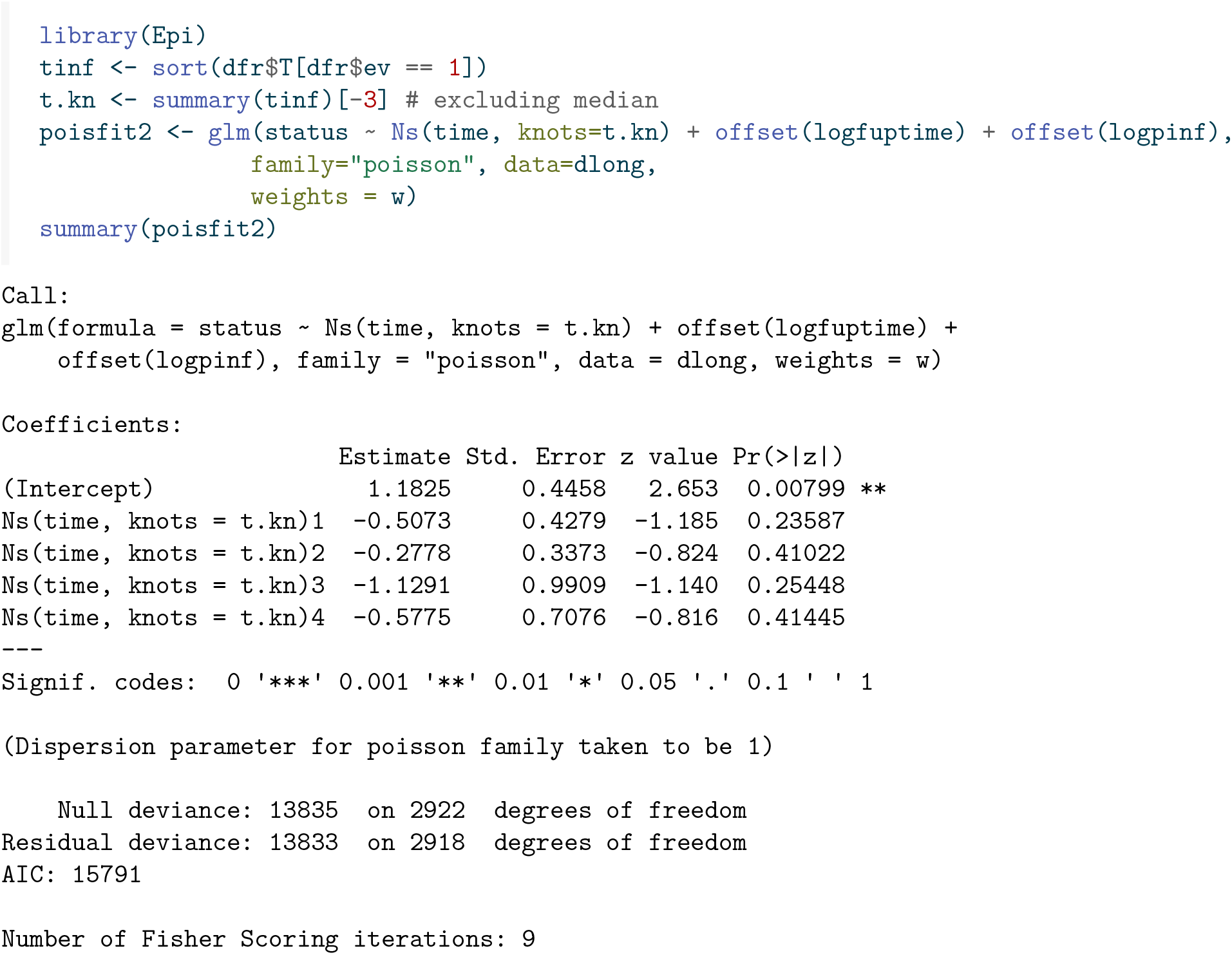

Note that none of the effects of the spline coefficients is significant. This is consistent with a constant infection rate. Otherwise the output is not very informative. It is more informative to make a plot of the fitted time-dependent infection rate. The function `matshade()` from the Epi package makes a nice plot, shown in Figure 5.

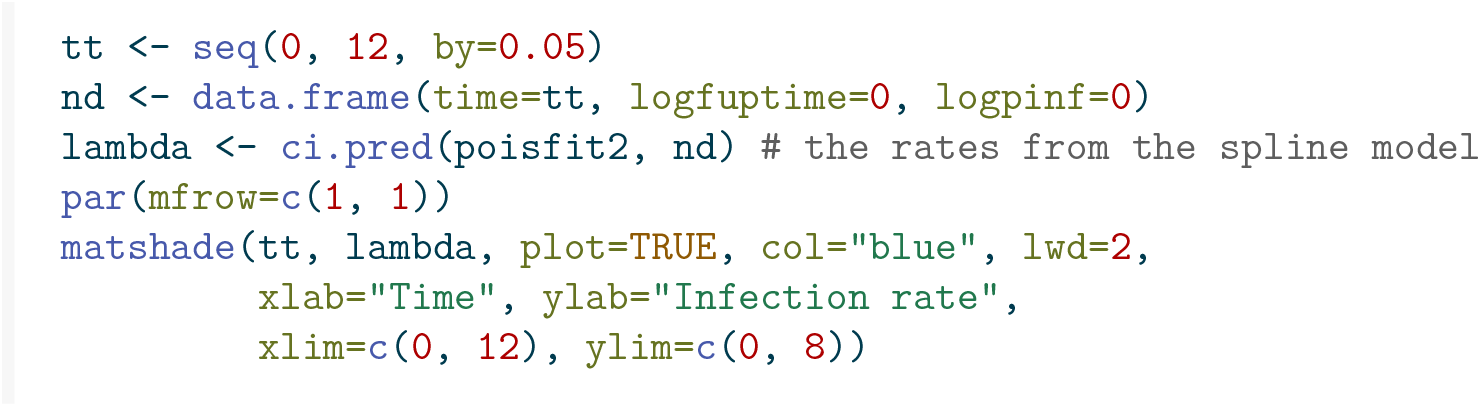

**Figure 5:**
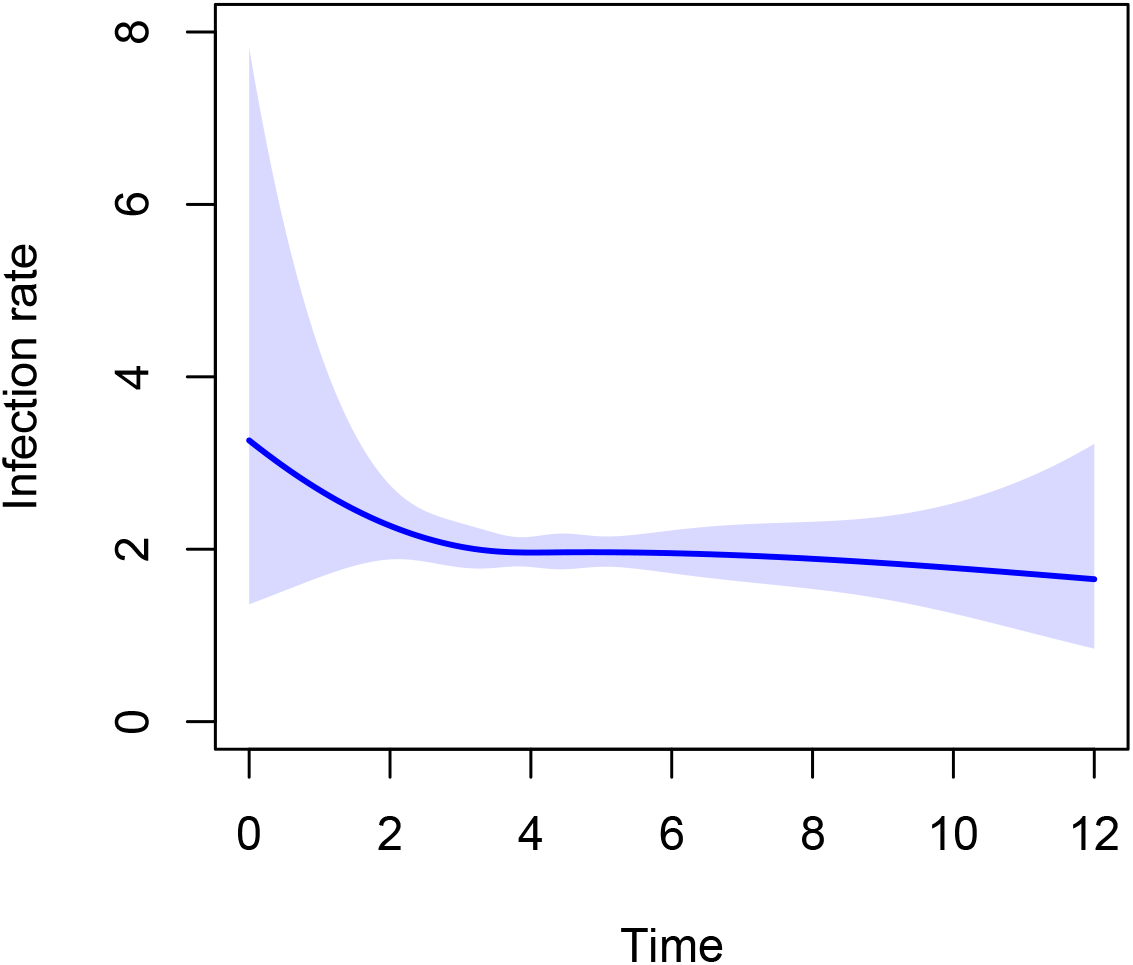
Smooth estimate of *β*(*t*).

It can be seen that the curve is more or less constant, with wider confidence intervals in the beginning.

## 4 Modeling perspectives

### 4.1 Additive versus multiplicative (Aalen versus Cox)

In the previous section we saw that for estimating *β*, when not assuming *β* to be constant, the estimate of *β*(*t*) obtained from the additive hazards model and the Cox model gave exactly the same result. The perspective of the additive hazards model is that we can view the rate *βĪ*(*t*) of a susceptible individual as additive in the number of infected individuals at time *t*; each infected individual *adds β/n* to the hazard. In contrast, the multiplicative hazards model views log(*Ī*(*t*)) as a multiplicative term, with regression coefficient fixed to one (an offset term).

Without any other covariates both perspectives are equally valuable and they give the same result when *β*(*t*) is estimated non-parametrically as a time-dependent function, and slightly different but comparable results when estimating a time-constant *β*. Different contexts will call for different modeling strategies, and we argue that in many cases a hybrid form might be advantageous.

### 4.2 Multiplicative models

Implementations of intervention measures are most naturally expressed in a multiplicative way. It makes sense to assume that such measures will decrease the hazard of every individual by a certain percentage. In ideal settings the effect of the intervention can be estimated together with the transmission parameter, using Poisson regression.

We are now going to explore the use of Cox and Poisson regression models in a situation where intervention measures are put into place, triggered here by the proportion of infected individuals reaching a certain threshold. The function below incorporates a single intervention, which is suspended by the time the proportion of infected individuals again reaches a lower threshold. The function is the same as before, with three extra parameters, the first, `effect`, being the effect of the intervention as a rate ratio, the second and third, `pinf_start` and `pinf_stop`, determining at what stage (in terms of percentage of infected individuals) the intervention is started and stopped.

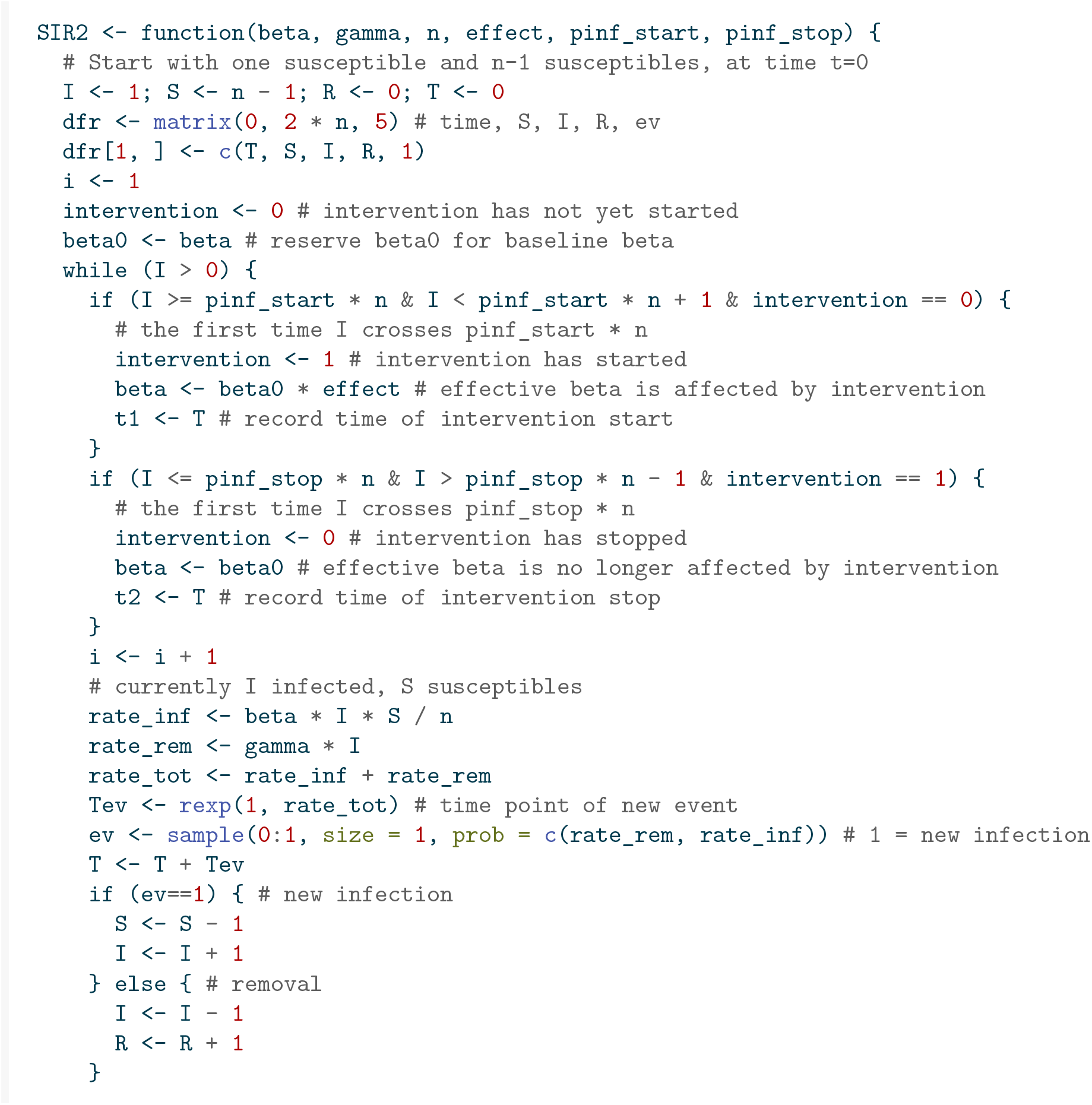
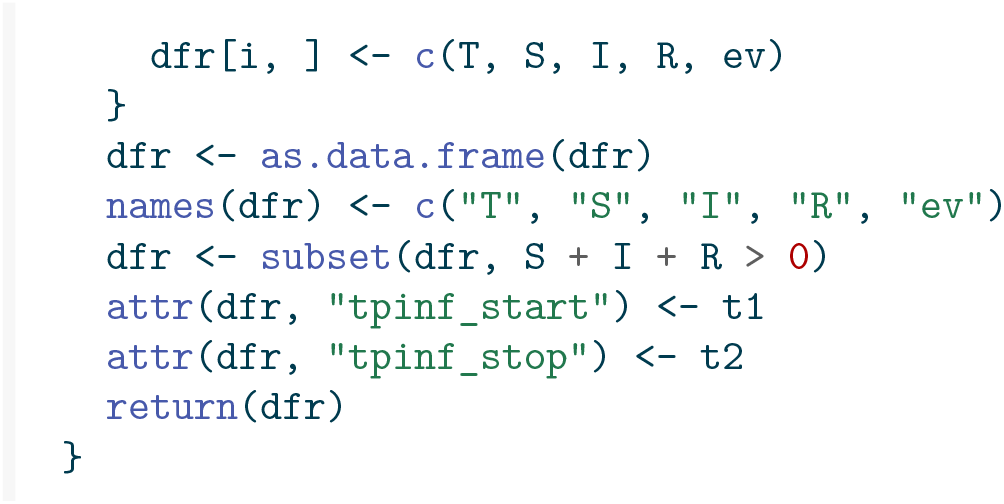

We now generate data according to this SIR model with intervention. The intervention is started as soon as 25% of the population is infected, and suspended as soon as that percentage has dropped to 10%. The effect of the intervention is 80%, in that the original transmission parameter *β* is 2, and after intervention it is 2 · 0.2 = 0.4.

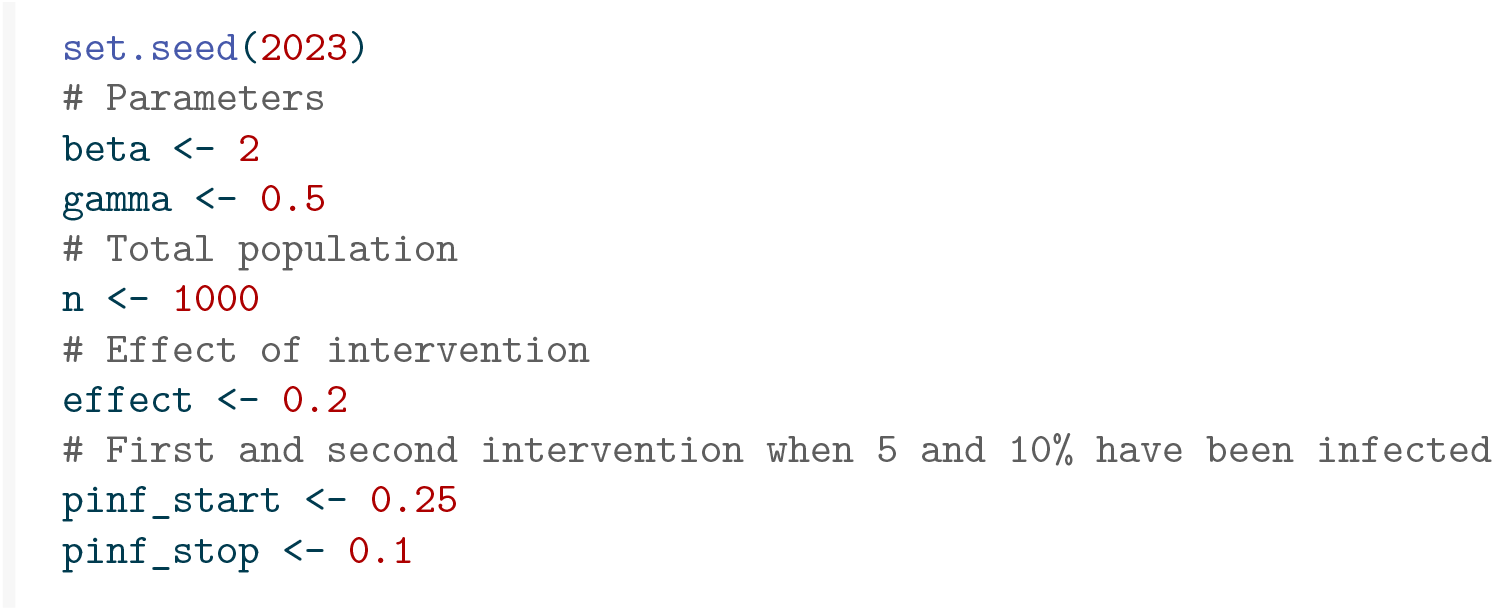

Figure 6 shows what the outbreak looks like. The time points where the intervention is implemented (when 25% of the population is infected) and stopped (when 10% of the population is infected) are indicated at the bottom.

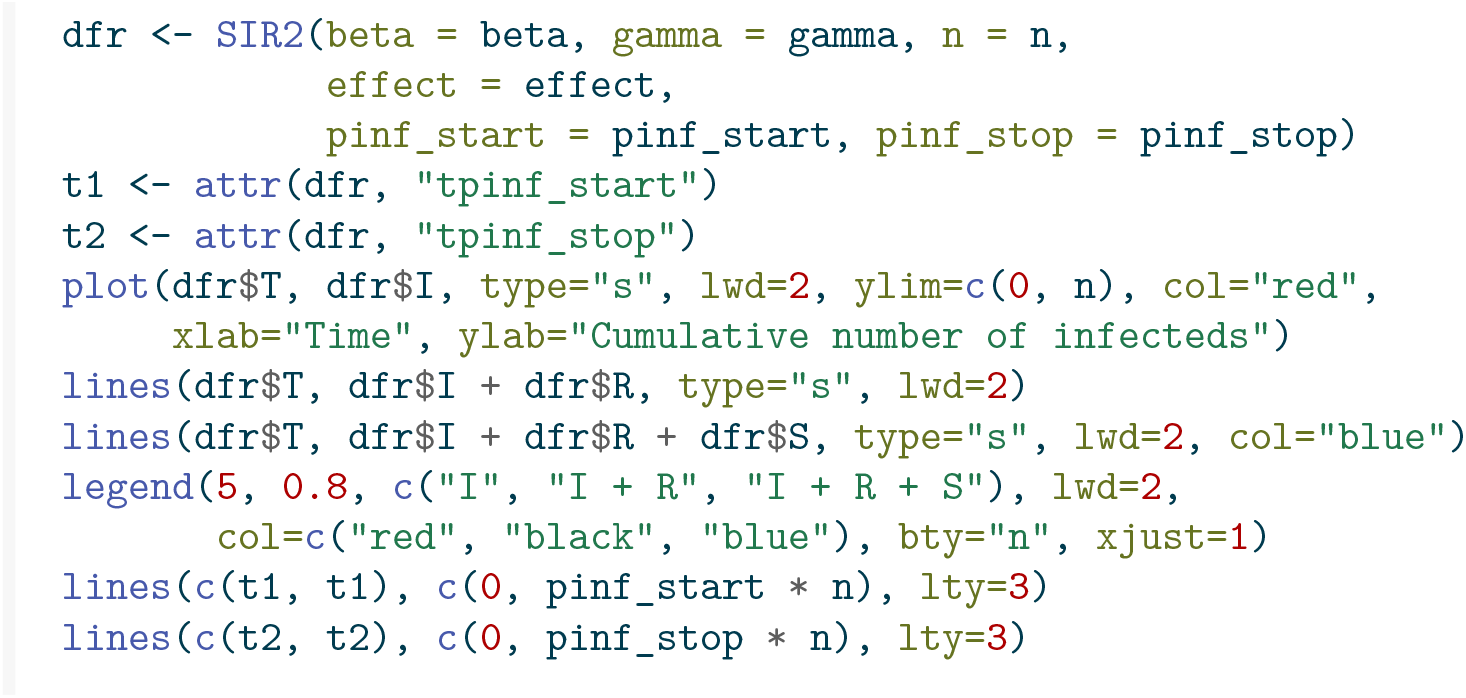

**Figure 6:**
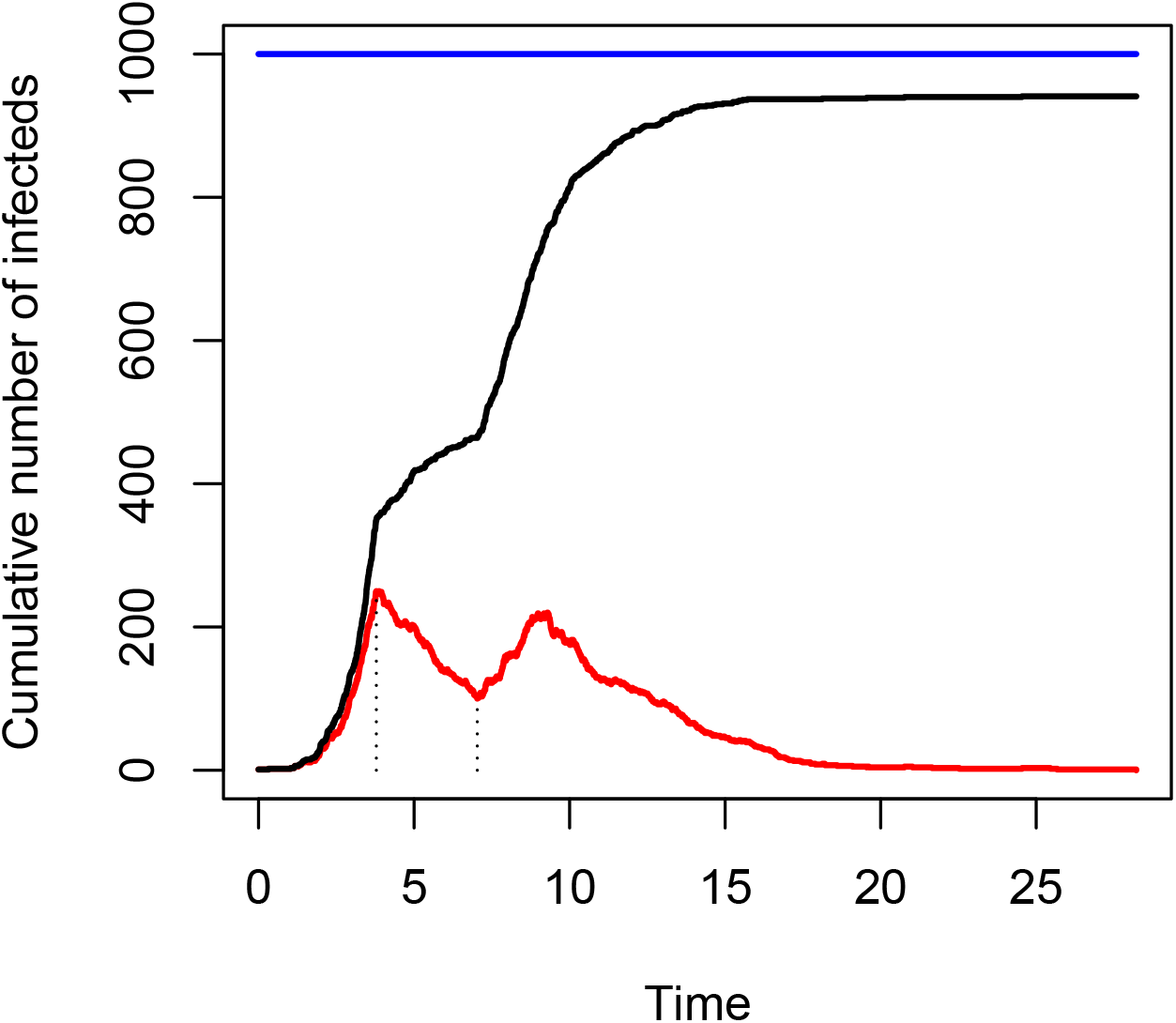
Stacked plot showing the number of susceptible, infected and recovered individuals over time, with intervention start and end indicated.

We see that after the intervention has been suspended the number of infections rises again, almost to 25%, but it decreases again just after that. We can again use the `SIR2surv` function to convert to data for use with Cox and Poisson modelling.

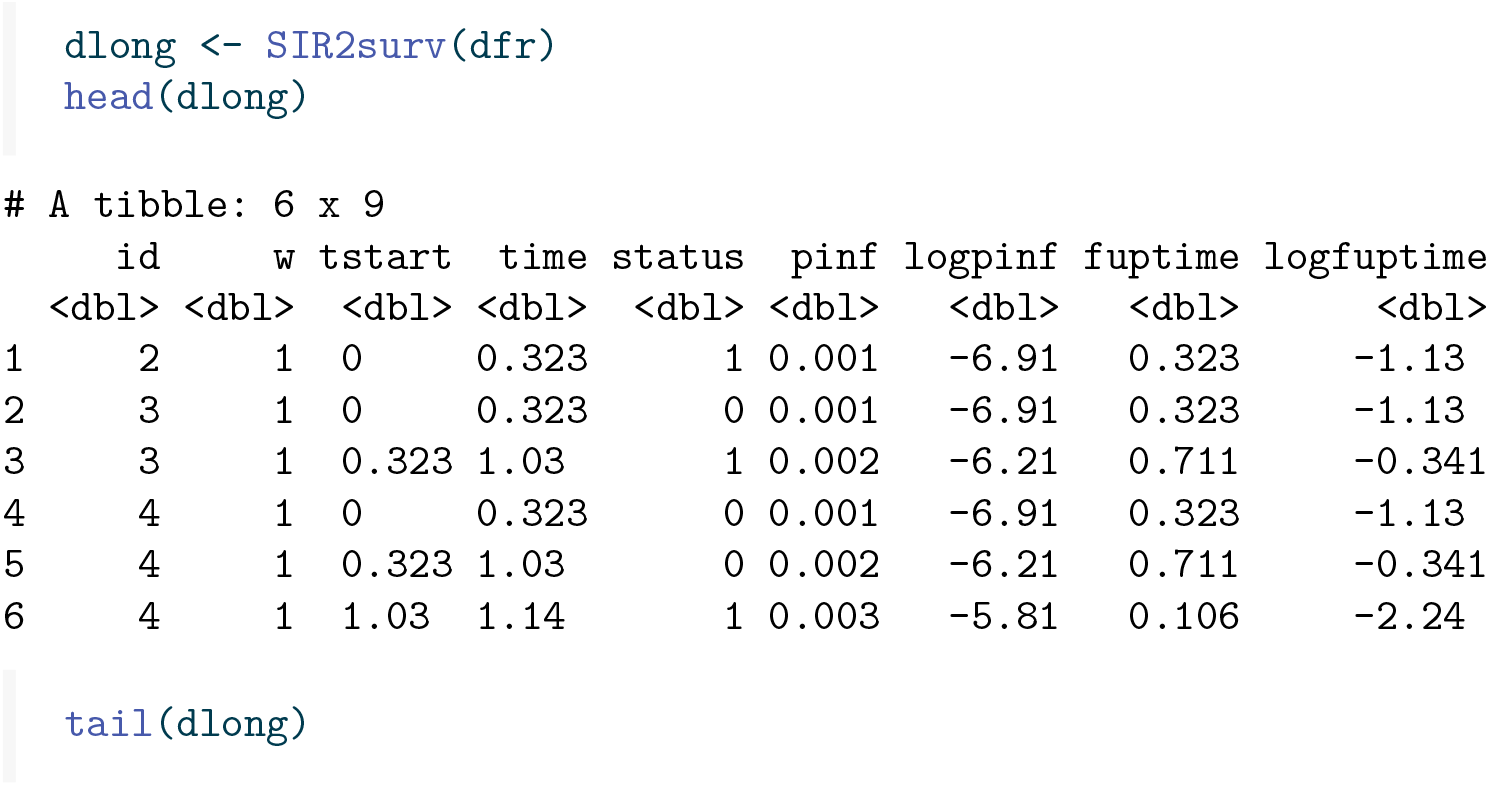
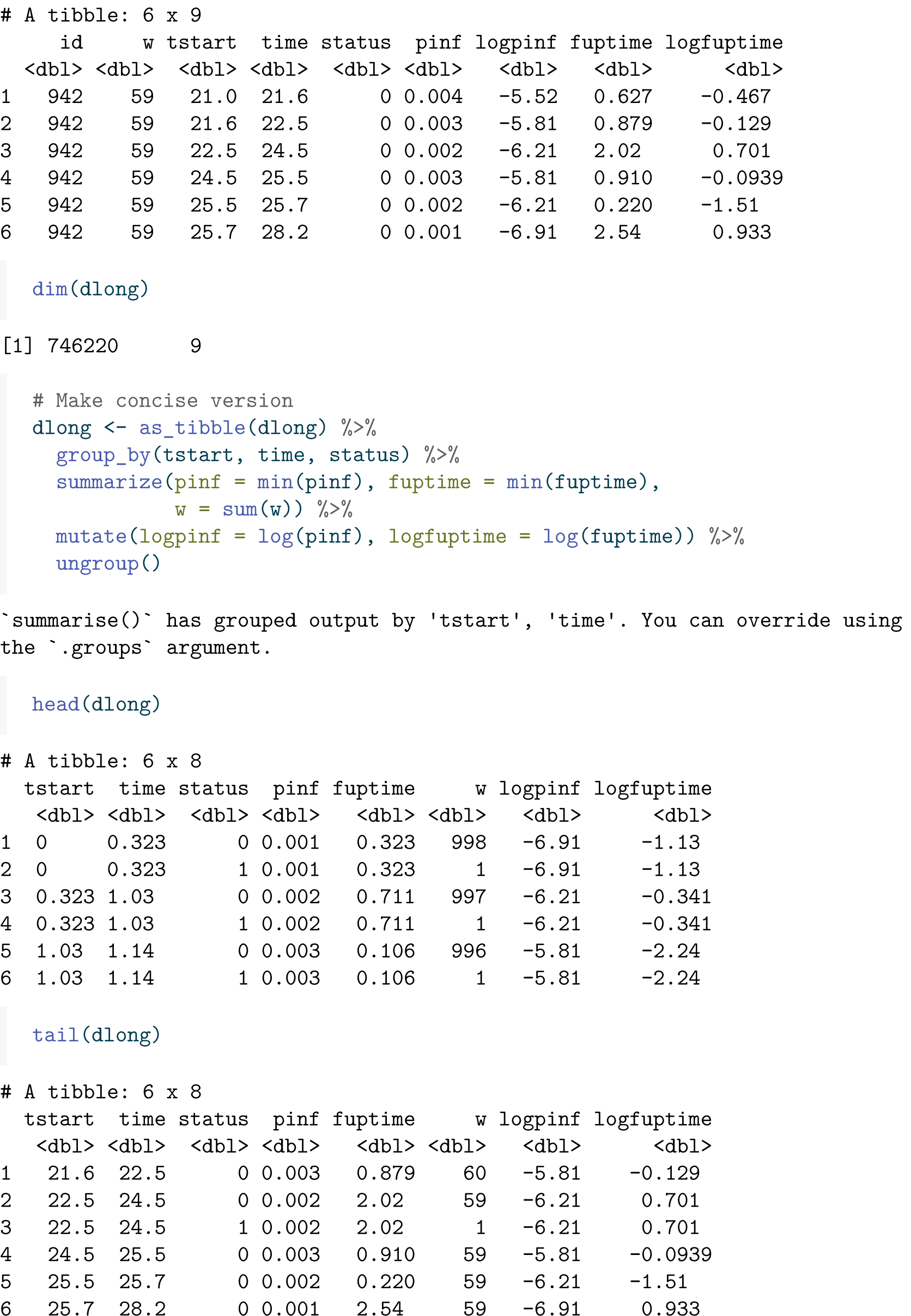
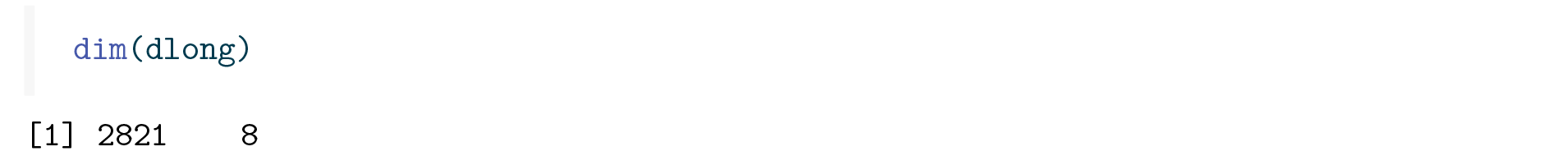

Note that we have included 59 subjects that were at risk for infection and never got infected during the epidemic. In the data it occurs as one subject with weight *w* = 59.

Let’s see whether we can pick up the changes in infection rates without knowing the times at which the interventions were in effect. Our first attempt is with a non-parametric estimate of the cumulative rate, using `coxph()`, shown in Figure 7.

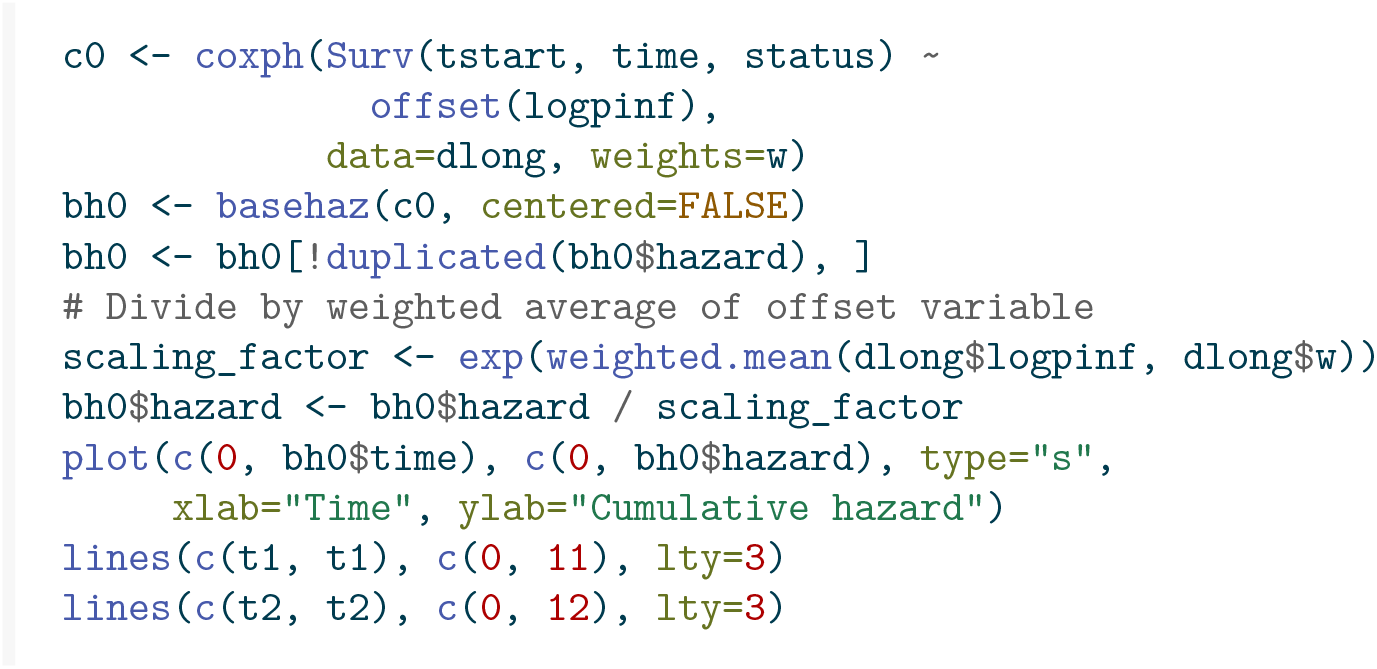

**Figure 7:**
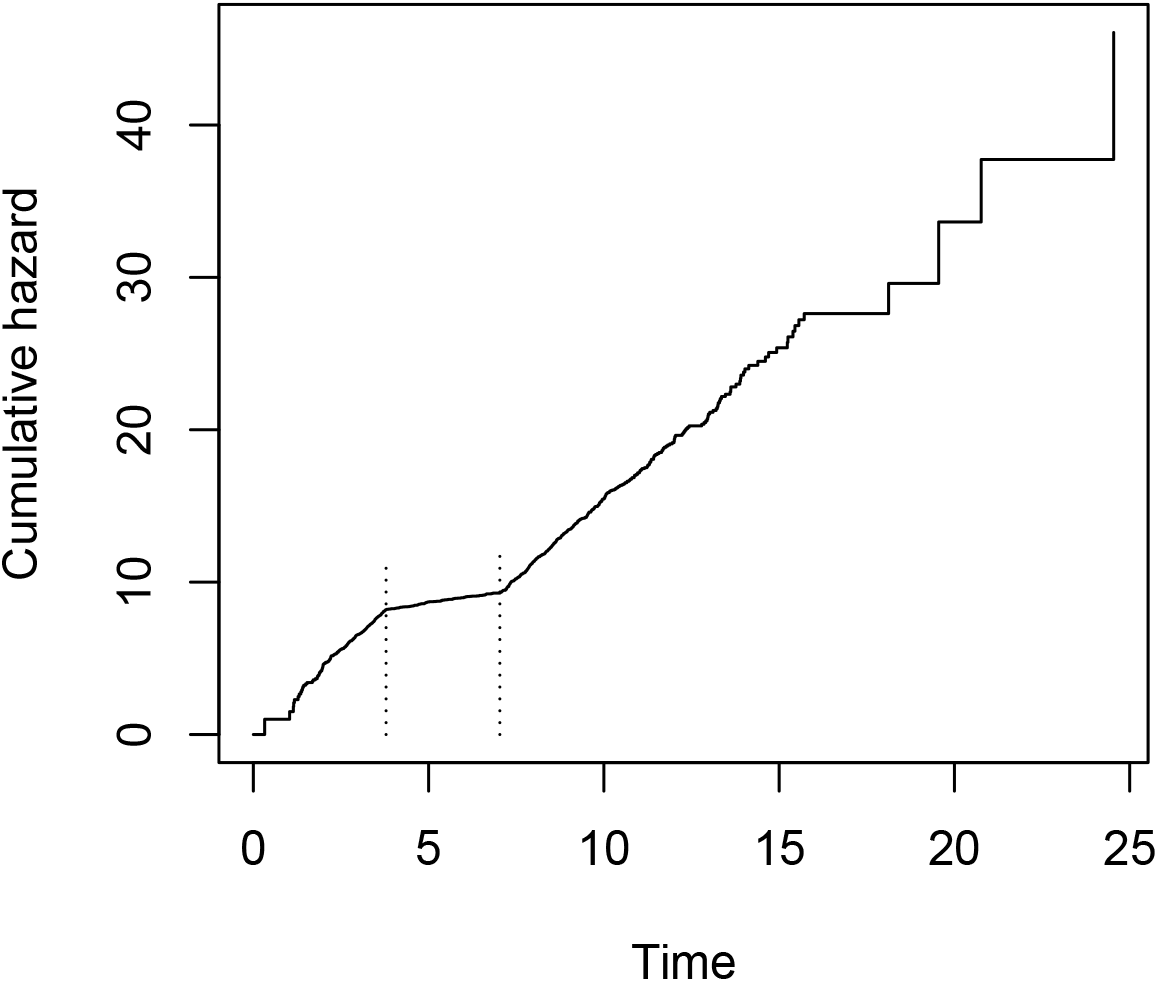
Baseline hazard of the Cox model under intervention.

Knowing the time of the intervention, a decreasing rate of infection can be seen, including an increase to the rate before intervention after the intervention has been suspended. We can also use Poisson regression again, with natural splines (same choice of knots as before, based on quartiles), to obtain a smooth estimate of *β*(*t*) (note, however, that the true *β*(*t*) is piecewise constant, not smooth).

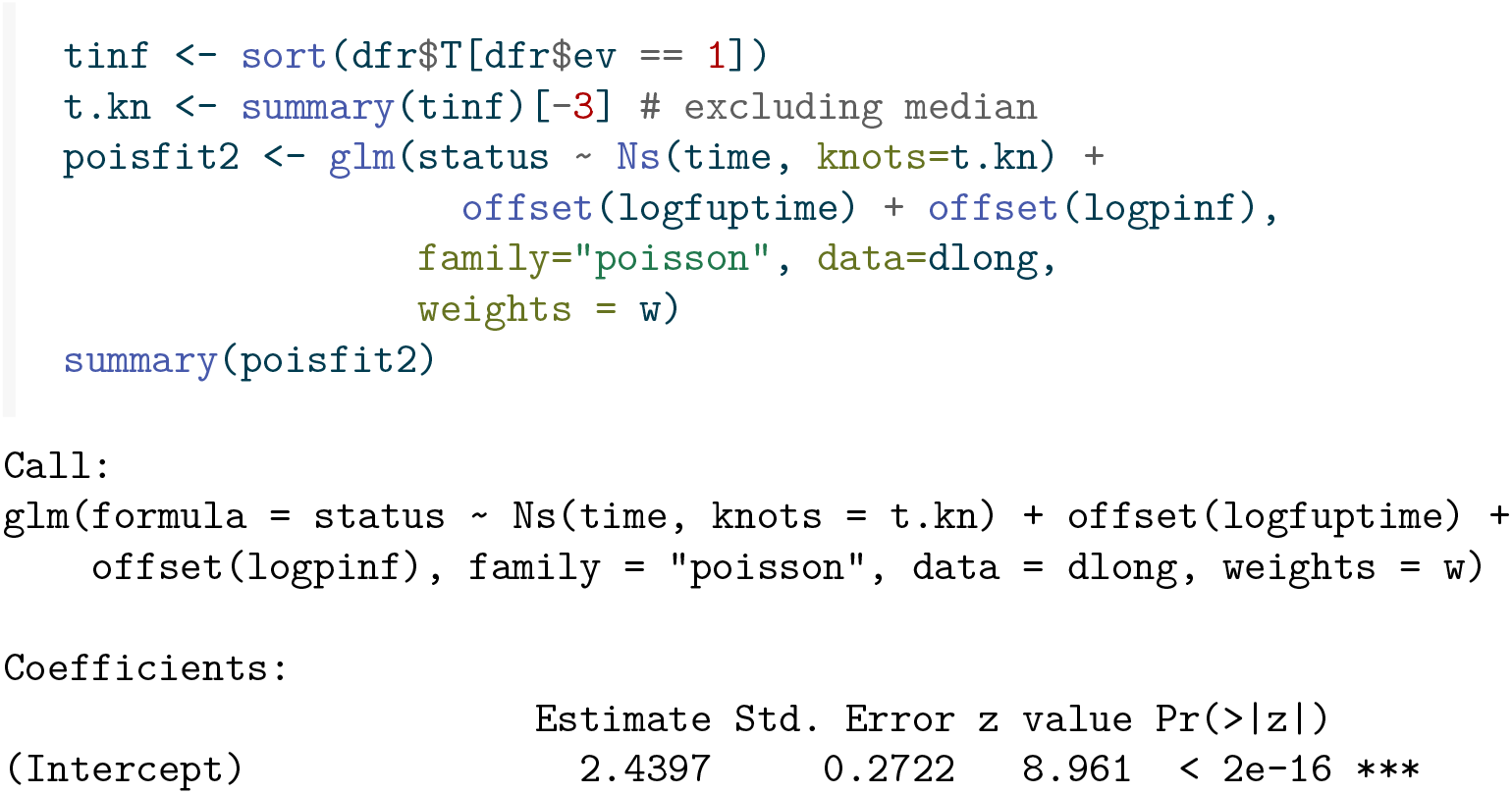
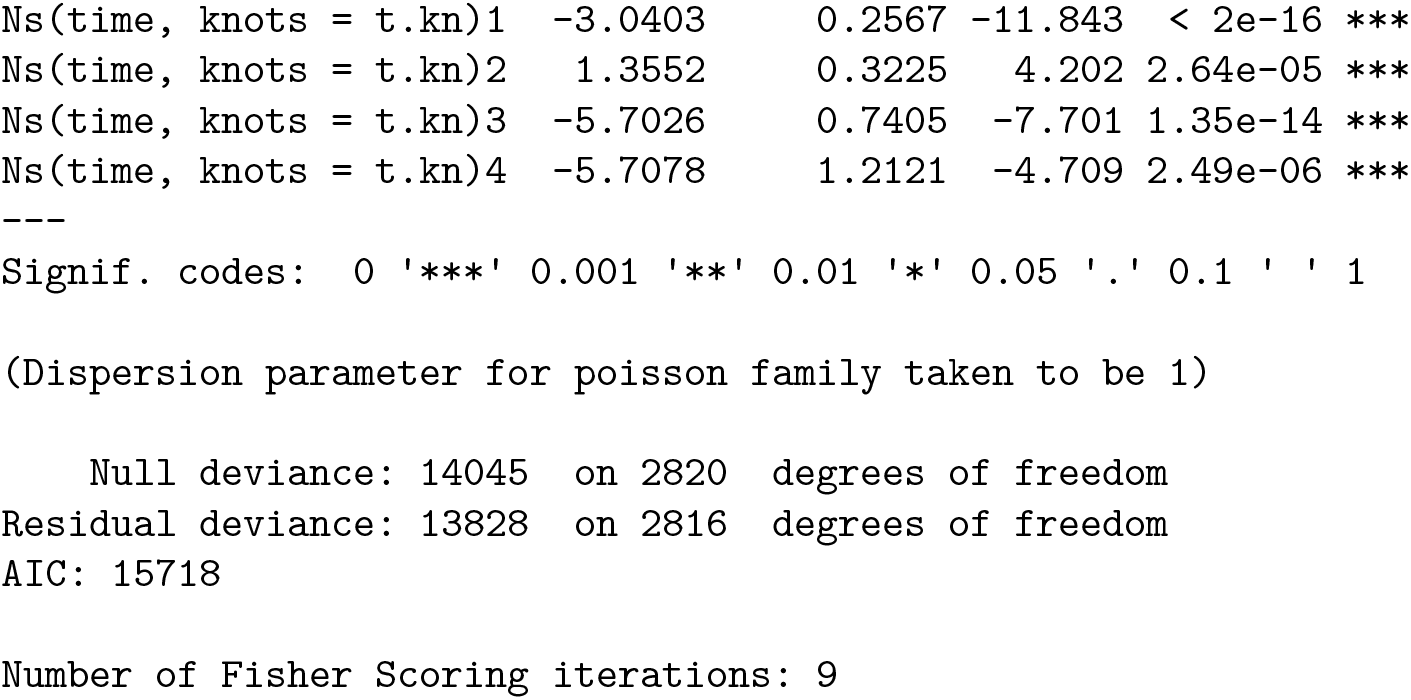

This time some of the non-constant effects of the splines are significant, pointing towards a non-constant infection rate. Let us make a plot of the infection rate *β*(*t*). The true *β*(*t*) is shown as dotted lines, in Figure 8.

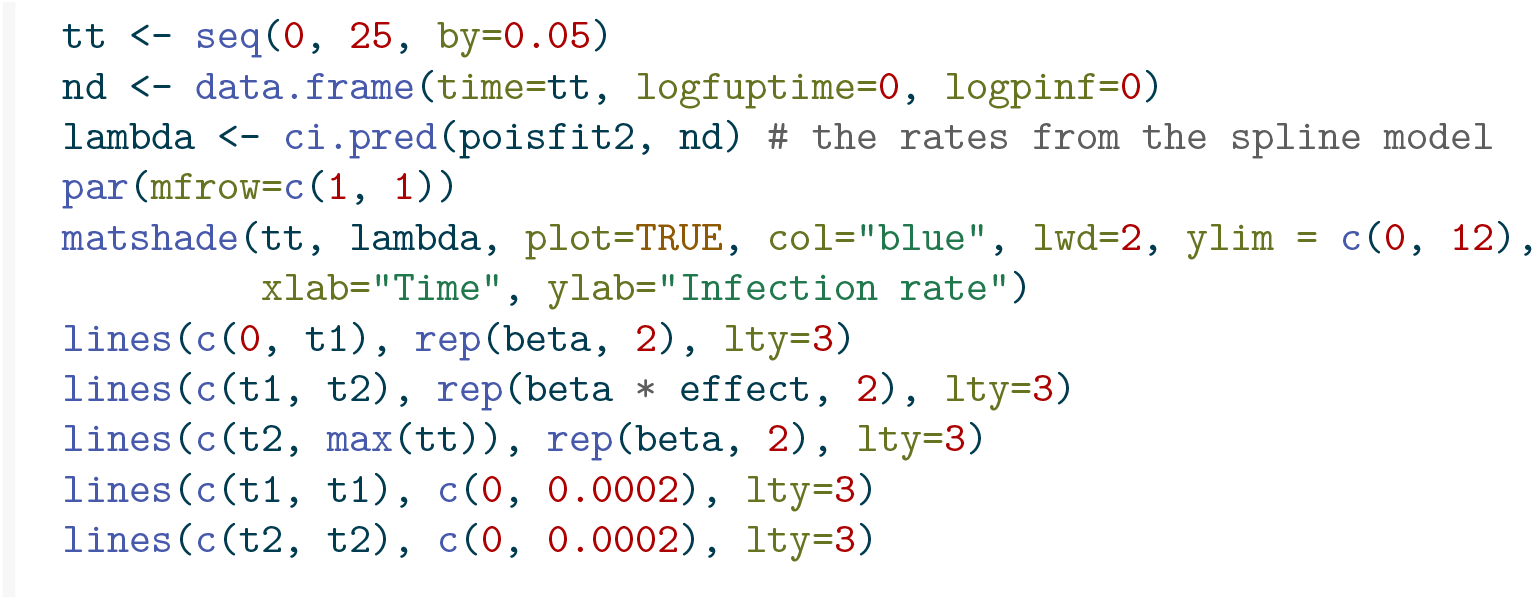

**Figure 8:**
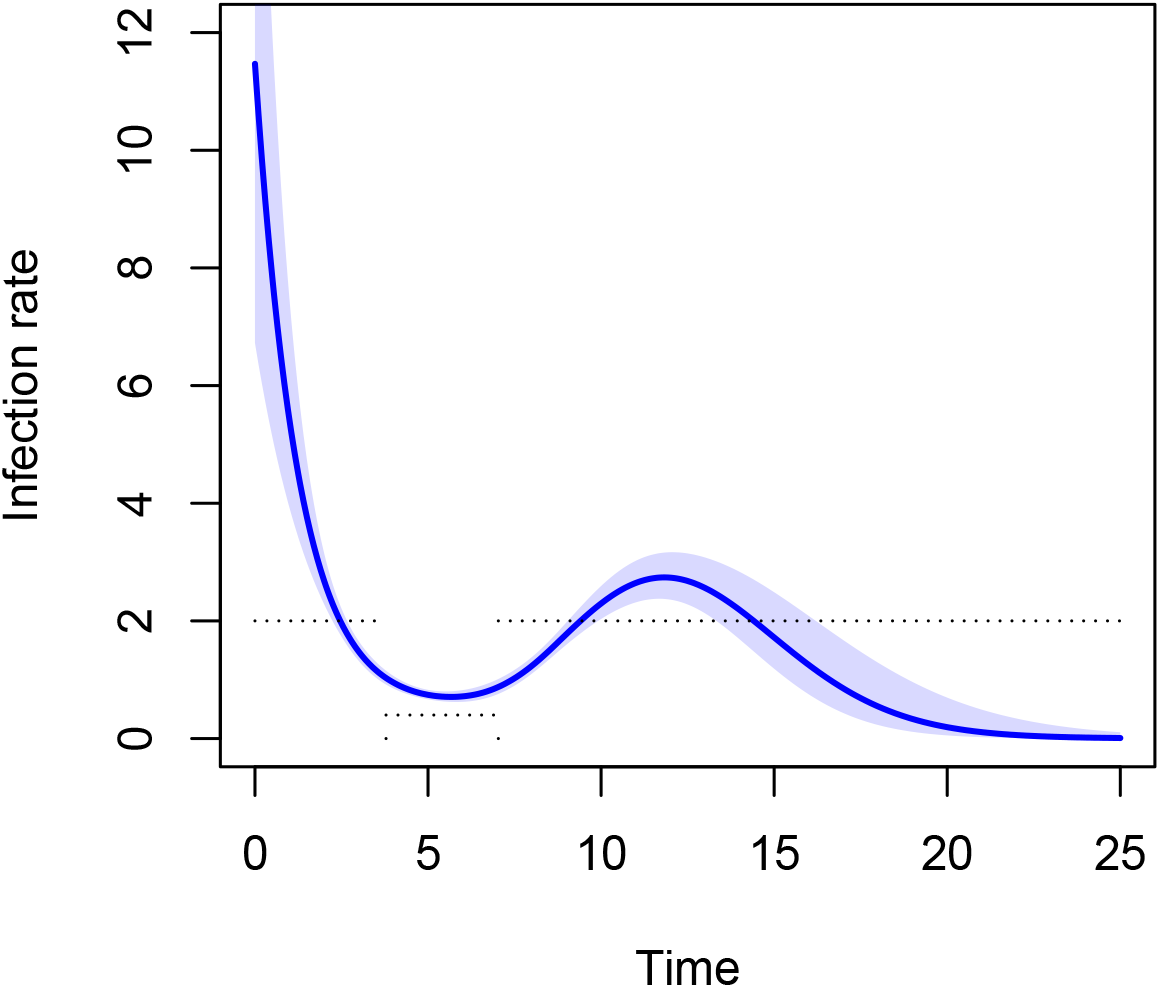
Time course of infection rate.

Because the natural splines try to impose a smooth line through a time-dependent rate that is inherently piecewise constant, the fitted curve is smoothly decreasing from above *β* = 2 to about *βη* = 2 * 0.2 = 0.4, and back to *β* = 2.

To be able to study the effect of the interventions, knowing when they were put in place and later suspended, we have to add information about the interventions to the data.

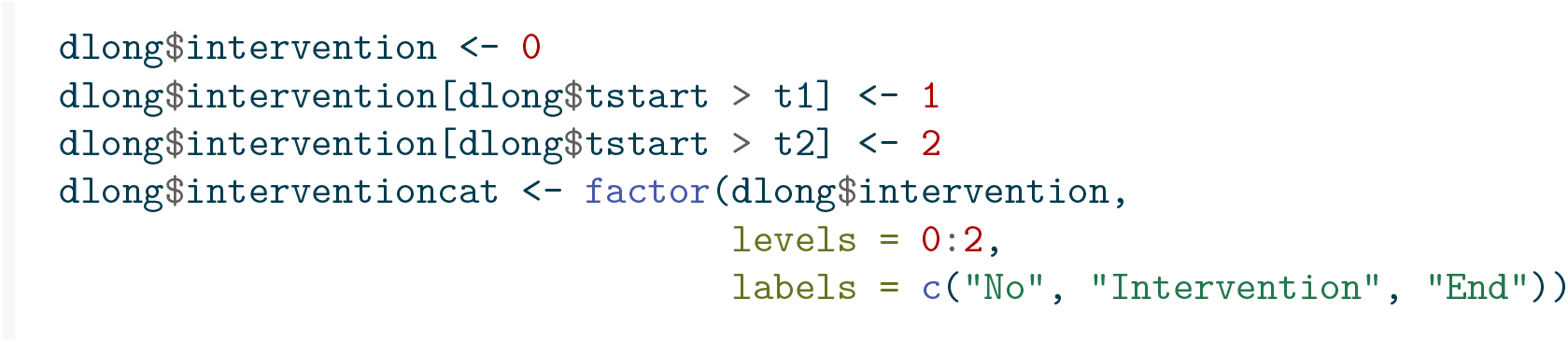

We start again with the epidemiological occurrence to exposure, this time by intervention period.

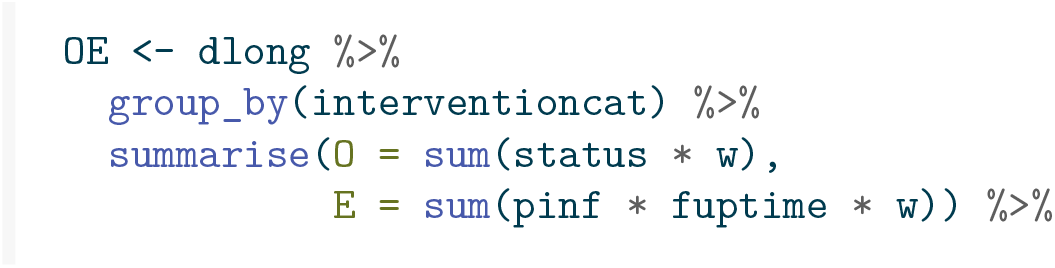
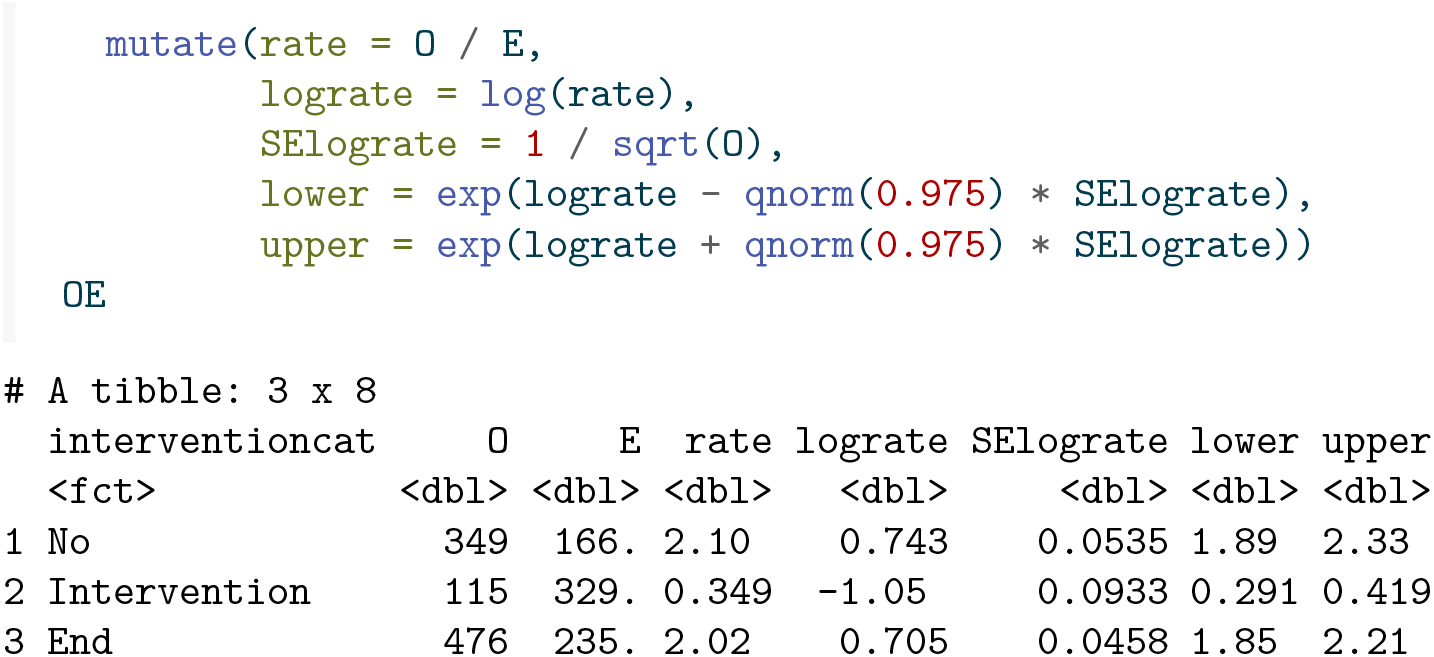

We nicely see that the estimated rates correspond reasonably to the infection rates of 2, 0.4, and 2 in the three periods.

A weighted Cox regression with intervention as categorical covariate (and logarithm of number of infected individual as offset), gives `NA`’s for estimates and standard errors (not shown). This is because the intervention is applied for all subjects in the same period, and hence the intervention effect is confounded with the baseline hazard. The intervention effect would be identifiable if different subjects would experience the interventions at different points in time.

The intervention effect is also identifiable with parametric restrictions on the baseline hazard, like a piecewise constant assumption. We now use weighted Poisson regression, again with log of interval time and log of proportion of infected individuals as intercept, and intervention as categorical covariate.

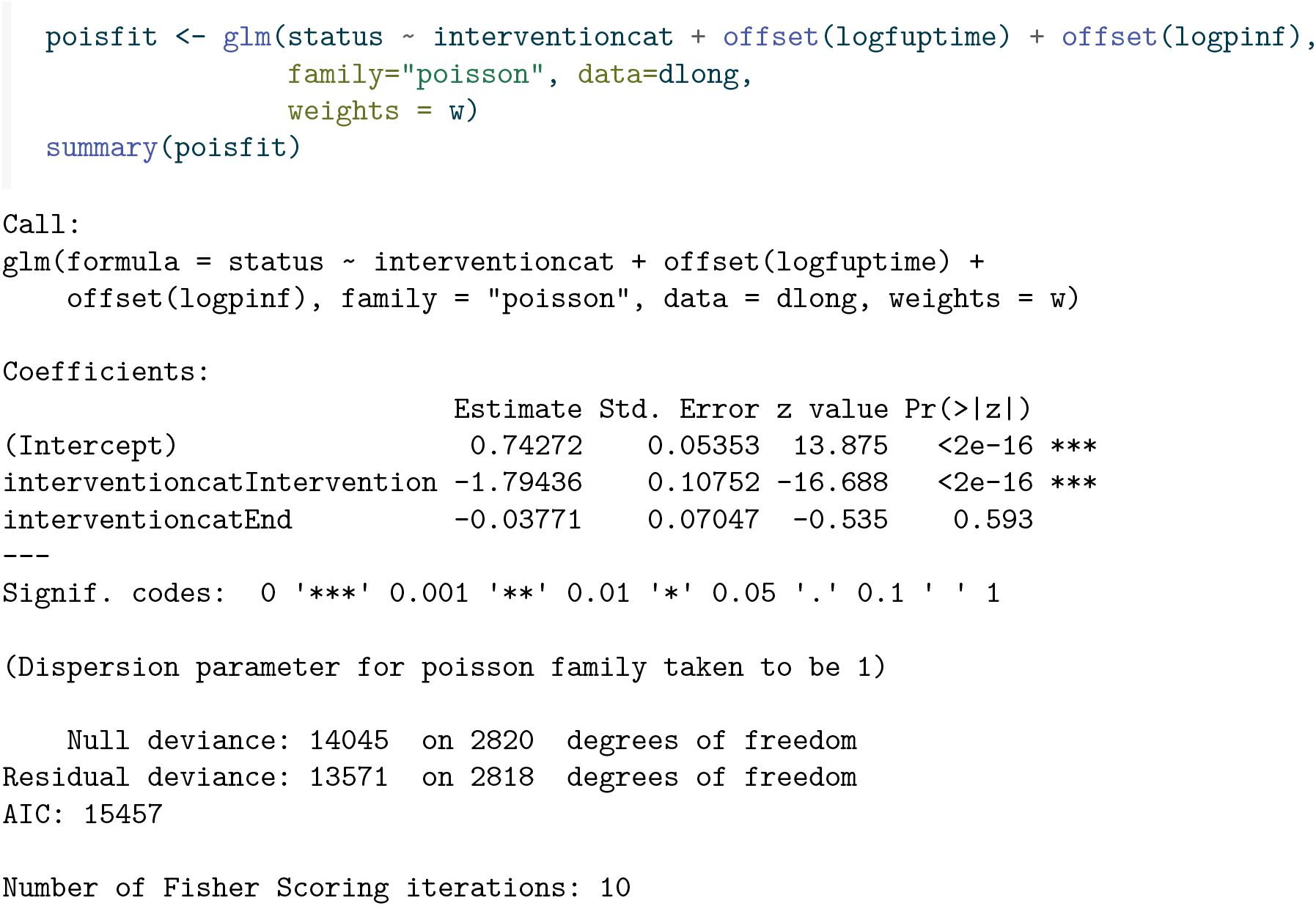
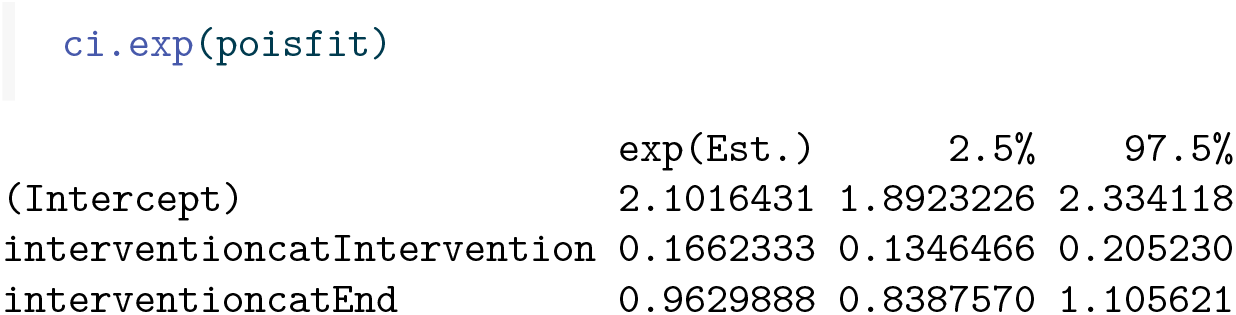

This retrieves the occurrence/exposure estimates, albeit with a different parametrization. The *β* in the pre-intervention period is the exponent of the intercept; the estimated regression parameter −1.794 is the contrast between the log *β* of the intervention period and the log *β* of the pre-intervention period (the intercept). The *β* in the intervention period, given by 0.349, can therefore be retrieved as the exponent of 0.743 + −1.794, which indeed equals 0.349. The *β* in the post-intervention period can be determined similarly.

The baseline infection rate, as well as the intervention effect, `effect = 0.2` is nicely recovered.

### 4.3 Additive hazards models

An interesting application where additive hazards occur very naturally is one where the population can be sub-divided into groups, such as age groups, occupation, households, schools, for instance, and where infections can occur between susceptibles and infected individuals within the same group or across groups.

The figure below shows the number of transmissions across different age groups in the Netherlands in March 2022. It nicely shows the number of transmissions are highest across similar age groups and after that across age groups differing by one generation (most probably transmissions within the same household). But they are numbers, some of which could be higher or lower simply because the groups sizes are different. The objective would be to estimate the infection rates across age groups.

We thus consider multiple groups (for instance age groups) that can infect each other. For group *j* = 1, …, *J* define *S*_*j*_(*t*), *Ī*_*j*_(*t*), *R*_*j*_(*t*) to be the total number of susceptible, infected and recovered individuals in group *j* at time *t*, and denote the total number of subjects in group *j* by *n*_*j*_ = *S*_*j*_(*t*) + *Ī*_*j*_(*t*) + *R*_*j*_(*t*). We again assume that the groups are closed.

Within group *j*, new infections happen with rate 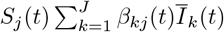, with *Ī*_*k*_(*t*) = *Ī*_*k*_(*t*)*/n*_*j*_. The idea is that each of the susceptibles in group *j* at time *t*, of which there are *S*_*j*_(*t*) can be infected by an infected individual from within group *j* itself or from within one of the other group. The (potentially time-varying) infection rate parameter *β*_*kj*_(*t*) describes the intensity at which contacts are made between an infected individual from group *k* and a susceptible individual from group *j* leading to a new infection. Finally we define the counting processes *N*_*j*_(*t*), *j* = 1, …, *J*, counting the total number of susceptibles in group *j* becoming infected within (0, *t*].

Interest is in estimating the (possibly time-varying) transmission parameters *β*_*kj*_(*t*). To illustrate methods for estimating *β*_*kj*_, we are choosing a setup with two groups (think of it as young and old), where the transmission parameter for susceptibles and infectious from the same group are higher (*β*_11_ = 3, *β*_22_ = 2) than across groups (*β*_12_ = 0.25, *β*_21_ = 0.5). The following code generates data from this model, with total population sizes of *n*_1_ = 1000 in group 1 and *n*_2_ = 750 in group 2. The choice for *γ* is 0.5 in each of the groups. The model starts with one susceptible in each group and *n*_*j*_ − 1 susceptibles in group *j*, at time *t* = 0, and then generates new events (infection in group 1, infection in group 2, recovery in group 1, recovery in group 2, denoted with ev = 1, 2, 3, 4, respectively) according to a Poisson process. The first 12 lines of the resulting data are shown.

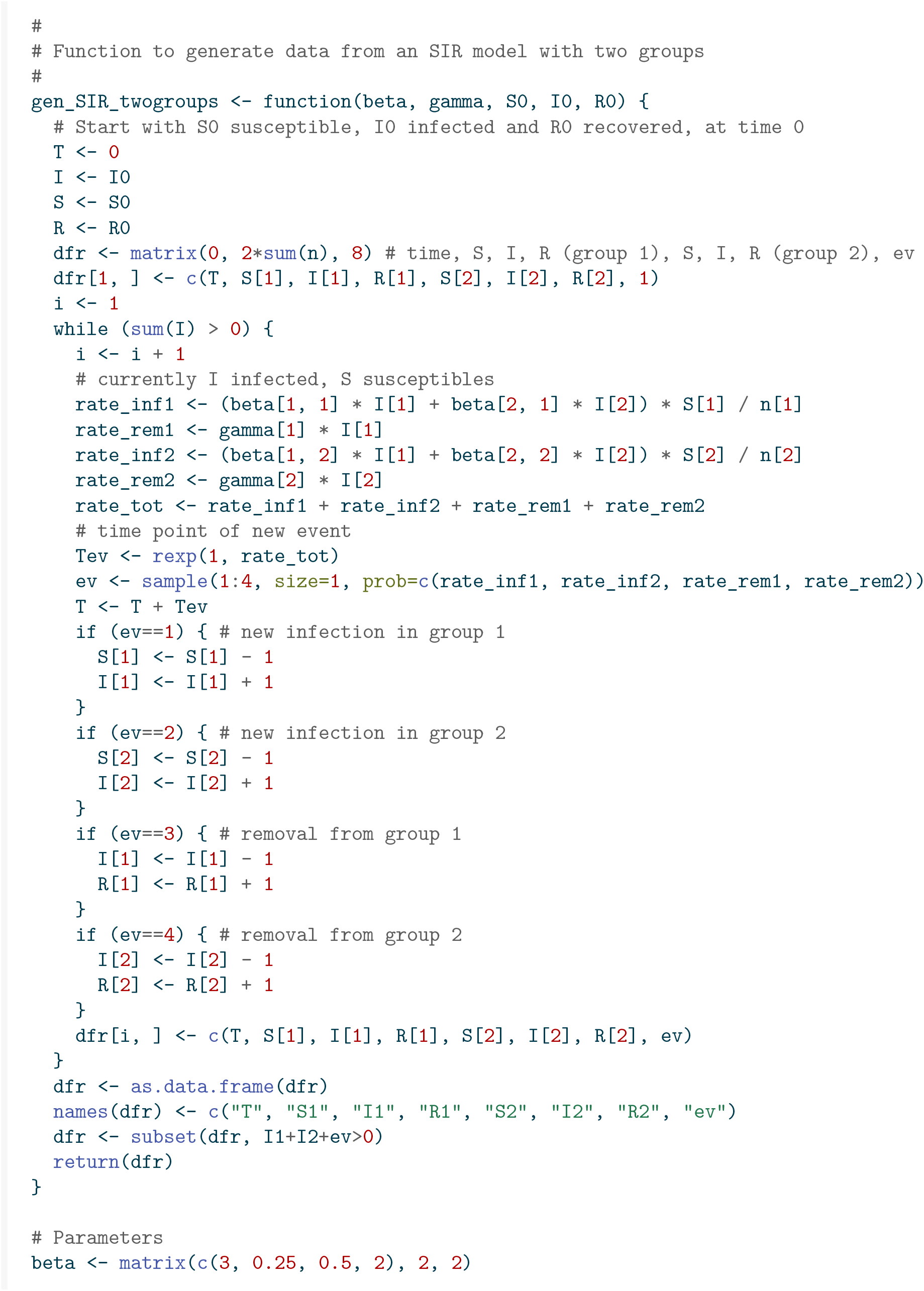
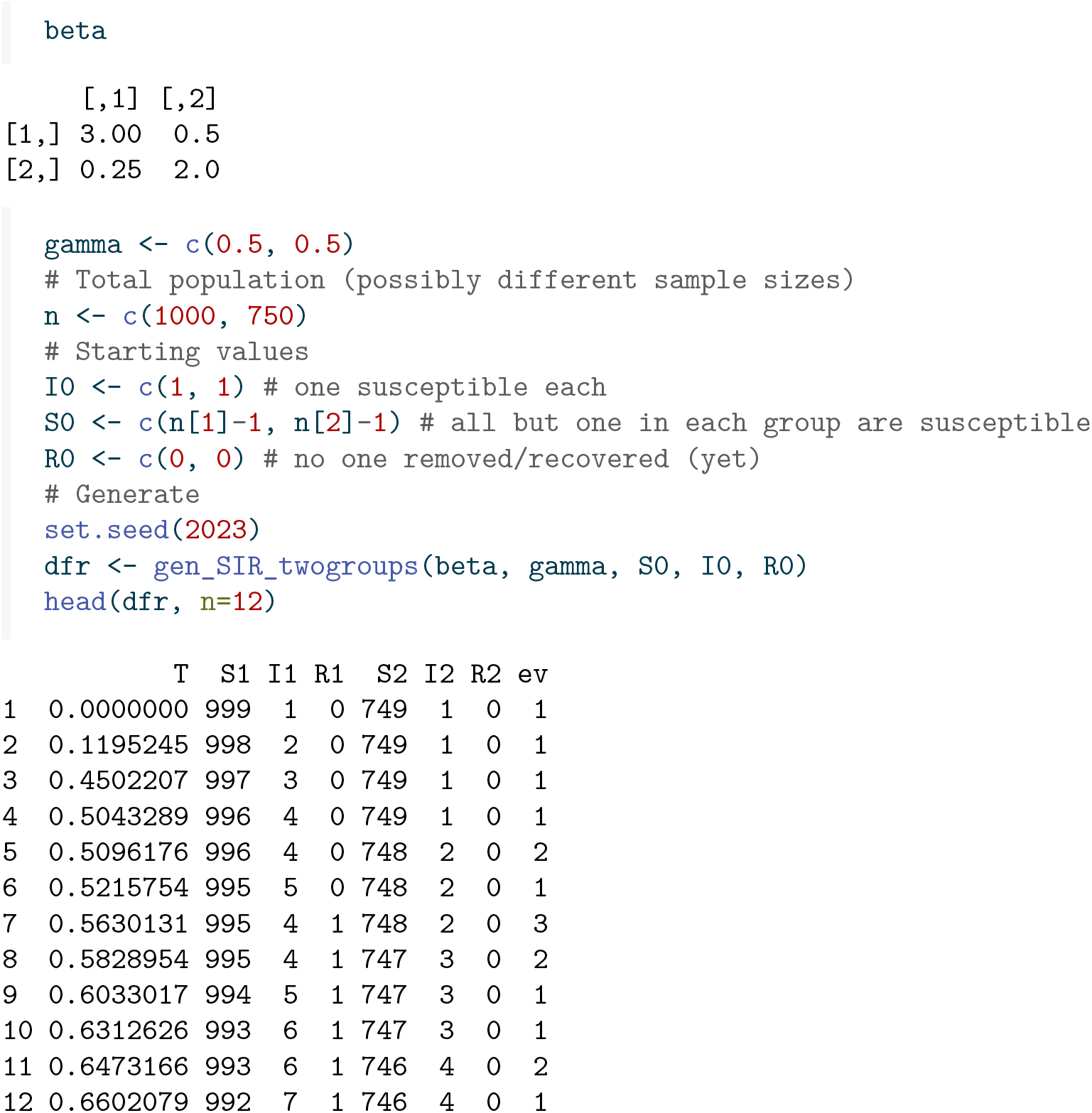

Figure 10 shows a plot (shown until *t* = 2) of the number of infected (solid line) and recovered (difference between dashed and solid lines) for group 1 (black) and group 2 (red) separately.

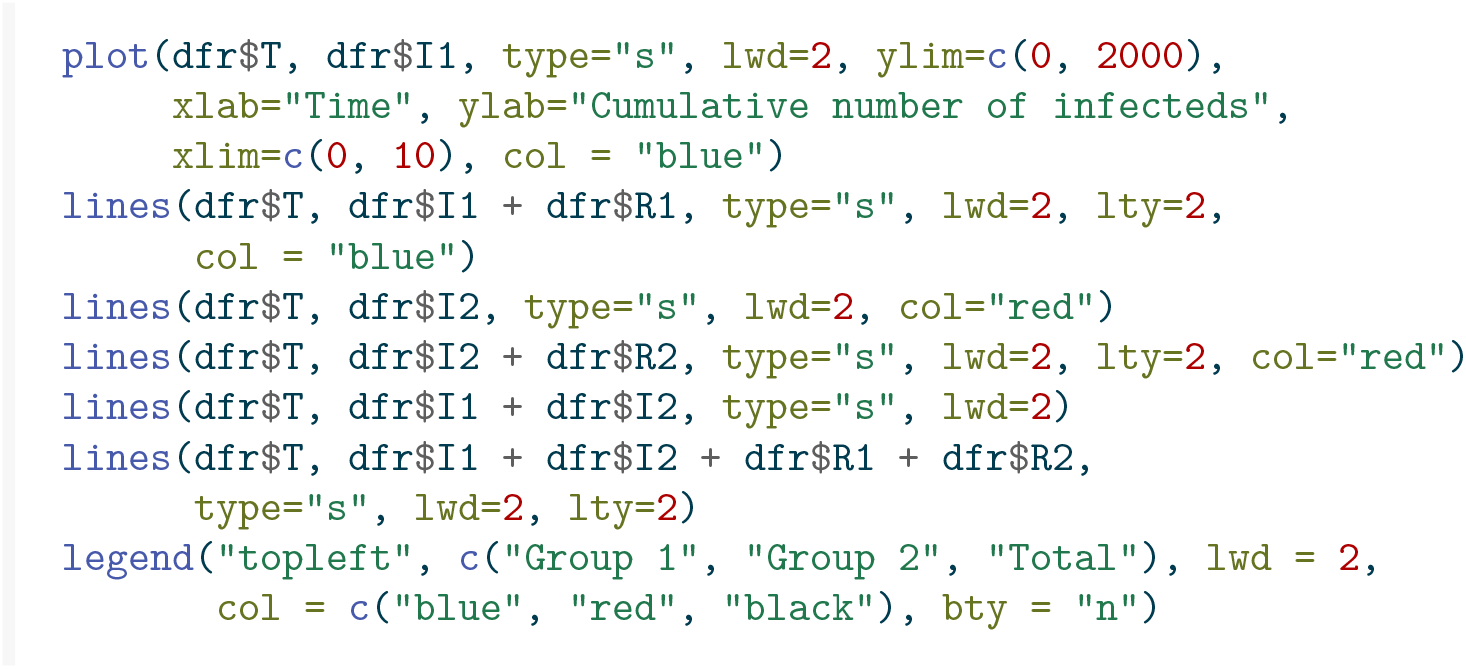

**Figure 9:**
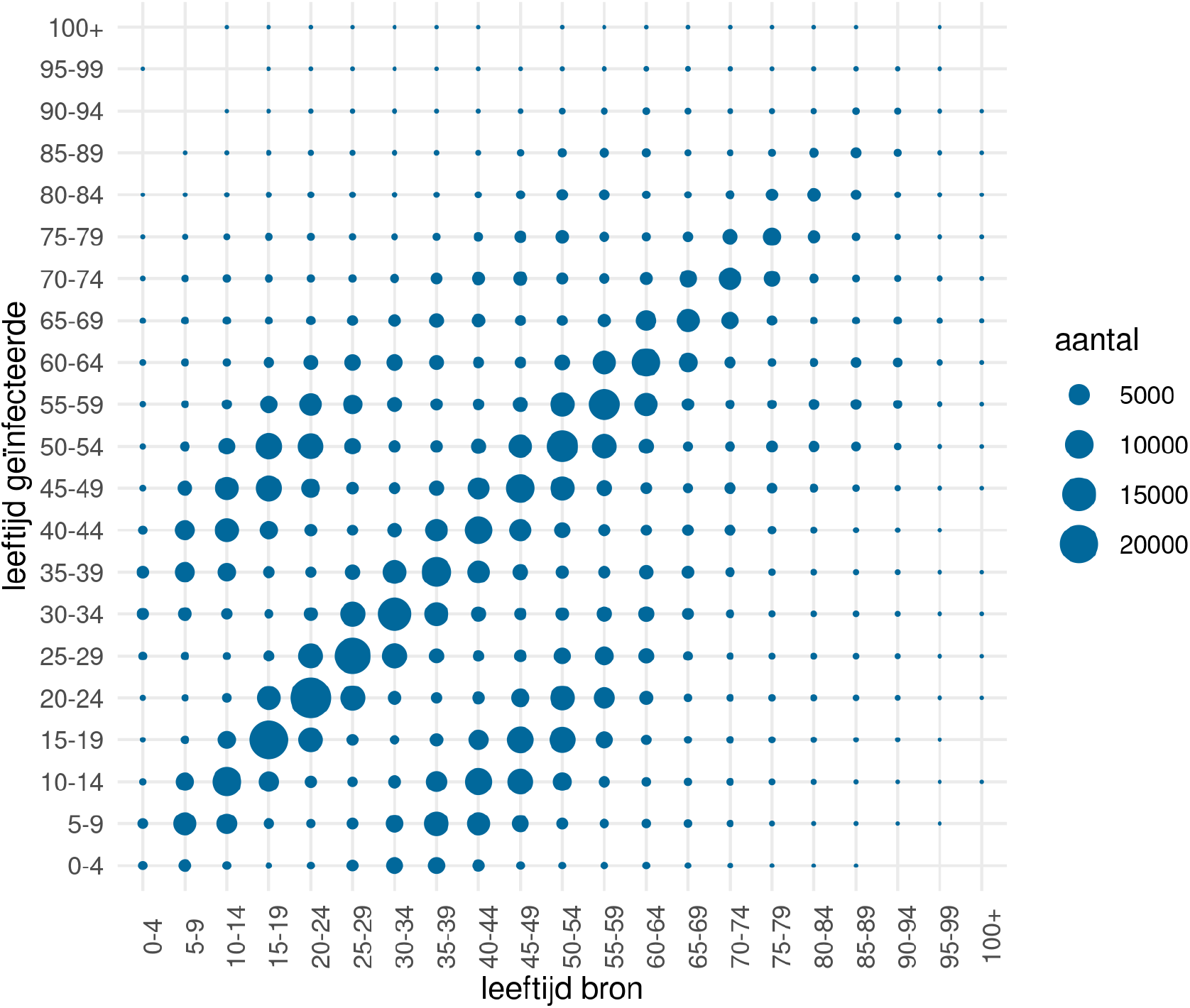
Transmissions across age groups in March 2022 in the Netherlands (leeftijd = age, bron = infector, geinfecteerde = infectee, aantal = number)

**Figure 10:**
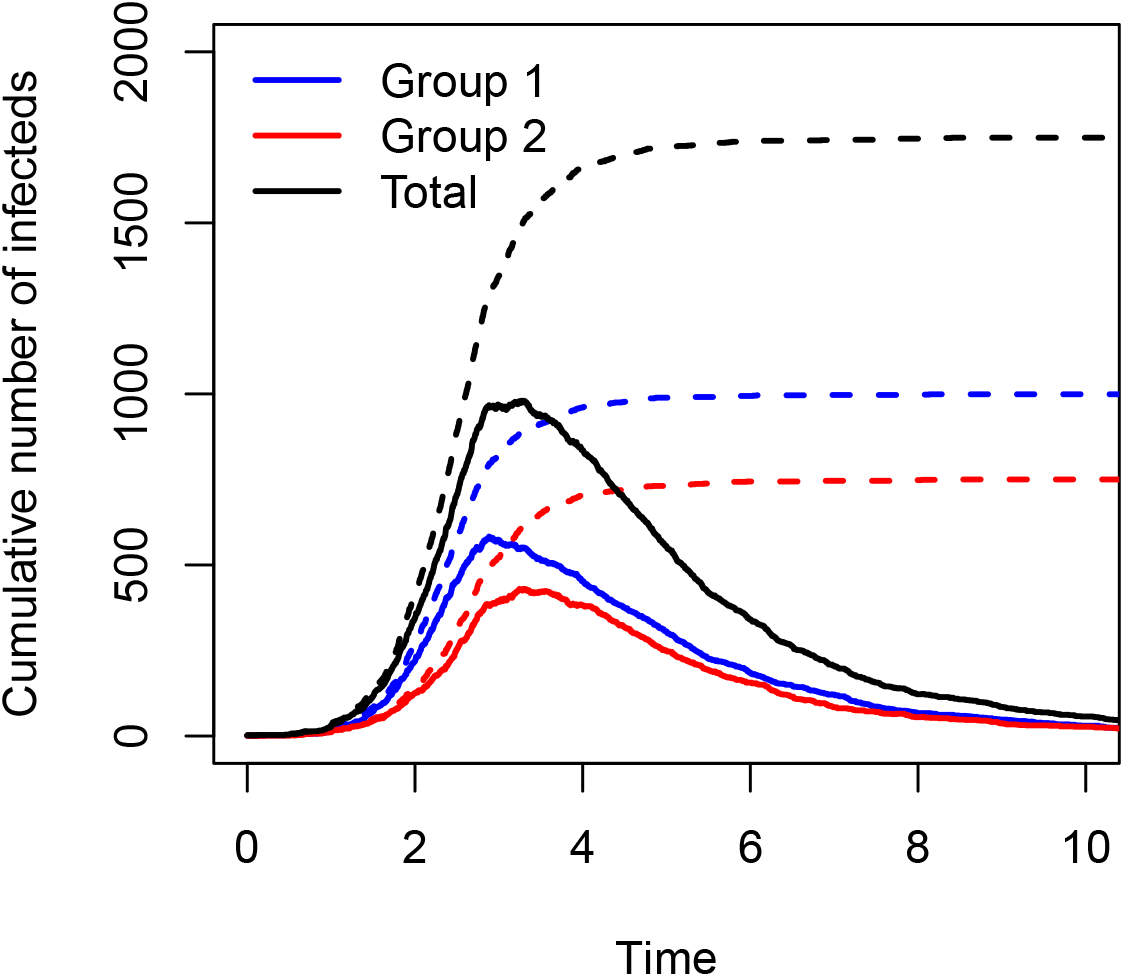
Number of infected and recovered individuals over time within group 1 (blue) and group 2 (red); total numbers in black.

We can adapt the `SIR2surv()` function to the situation of two groups.

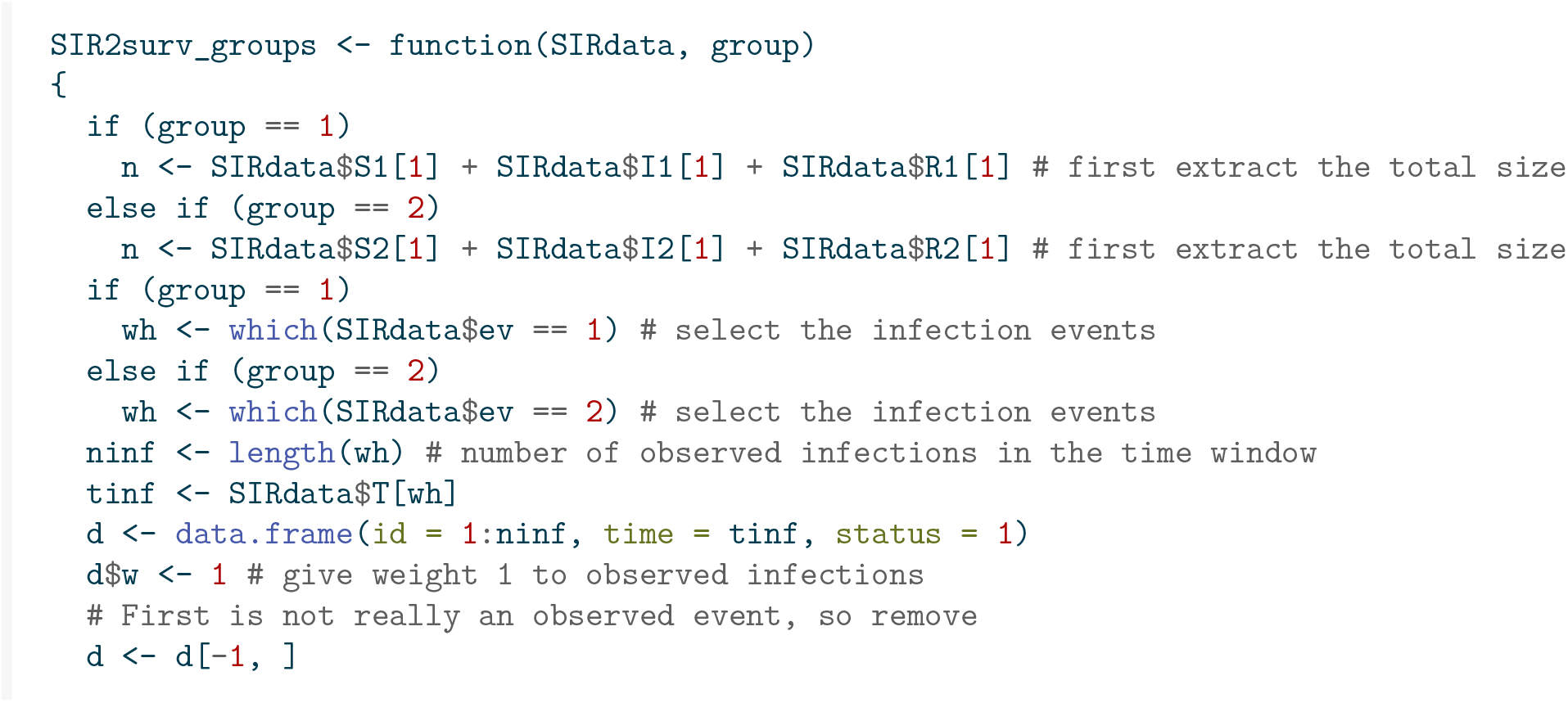
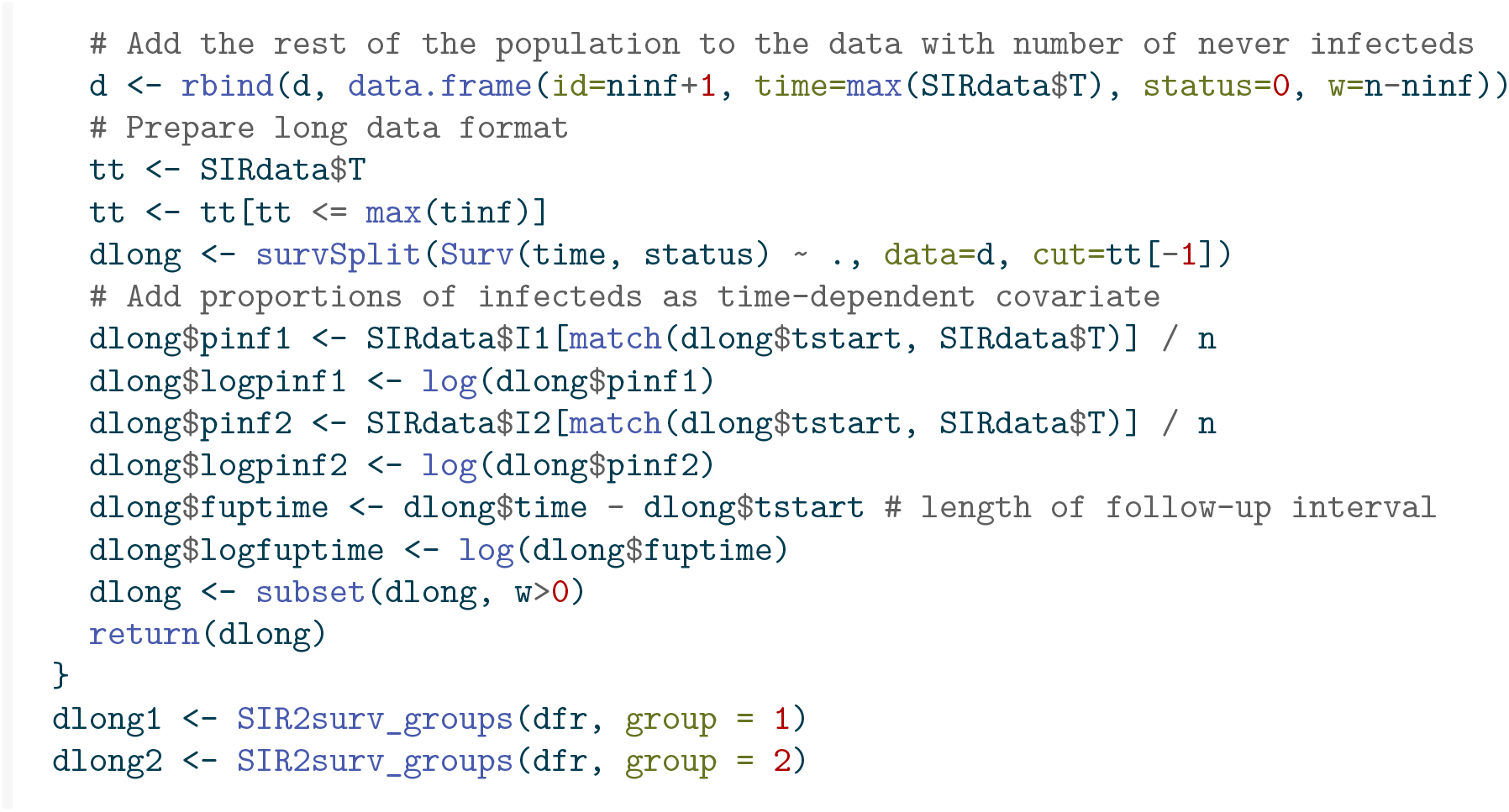

Again we can look from the individual perspective, define a counting process for each individual *I* from each group *j, N*_*ji*_(*t*), having rate 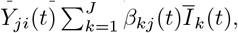, where *Ī*_*k*_(*t*) = *Ī*_*k*_(*t*)*/n*_*j*_, with *n*_*j*_ the size of group *j*. This is an *additive hazards model* without intercept, and *J* time-dependent covariates *Ī*_*k*_(*t*), where *Y*_*ji*_(*t*) is the at risk indicator of subject *i* in group *j* for being susceptible to infection.

To simplify notation, consider one group *j* of susceptibles. We will fix that group, suppress *j* in the notation everywhere, and let *n* be the size of that group. Within this group, individual *i* has rate

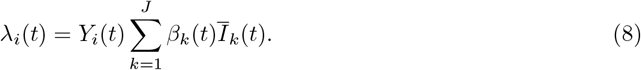

This rate conforms to the additive hazards model with rate 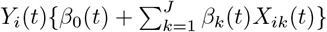, with two non-standard aspects. The first is that in Equation 8 there is no intercept *β*_0_(*t*), the second is that in Equation 8 for each time point *t* the time-dependent covariates *X*_*ik*_(*t*) are the same for all susceptible individuals. This has the important implication that the *k*th column of the matrix **X**(*t*) used in Equation 4 is of the form **Y**(*t*)*Ī*_*k*_(*t*), **Y**(*t*) = (*Y*_1_(*t*), …, *Y*_*n*_(*t*))^⊤^. As a result **X**(*t*) is of rank 1, which implies that the *β*_*k*_(*t*)’s are not identifiable from the data when the *β*_*k*_(*t*)’s are allowed to vary freely. To be able to estimate the *β*_*k*_(*t*)’s one would need some kind of smoothing or restricting the *β*_*k*_(*t*)’s to be constant or piecewise constant on time intervals where for instance interventions are put into place and/or suspended.

In the more restricted model where *β*_*k*_(*t*)’s are assumed to be constant, the *β*_*k*_’s *are* identifiable. Two approaches are possible to estimate the *β*_*k*_ coefficients. The first is maximum likelihood, as an extension of Section 2.2. Define ***β*** = (*β*_1_, …, *β*_*J*_)^⊤^, **X**_*i*_(*t*) = (*X*_*i*1_(*t*), …, *X*_*iJ*_ (*t*)), which here simplifies to **X**_*i*_(*t*) = **I**(*t*) = (*Ī*_1_(*t*), …, *Ī*_*J*_ (*t*)). The log-likelihood of ***β*** is given by

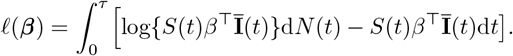

Taking derivatives with respect to the elements of ***β*** and setting to zero gives

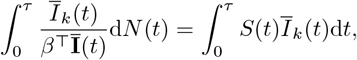

for *k* = 1, …, *J*, which can be solved numerically. The second is an extension of the approach of Lin and Ying (1994). The latter allows to use

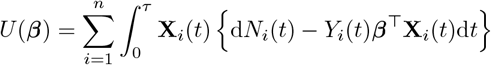

as estimating equations, with ***β*** = (*β*_1_, …, *β*_*J*_)^⊤^, and **X**_*i*_(*t*) = (*X*_*i*1_(*t*), …, *X*_*iJ*_ (*t*)), which here simplifies to **X**_*i*_(*t*) = **I**(*t*) = (*Ī*_1_(*t*), …, *Ī*_*J*_ (*t*)), independent of *i*.

This leads to

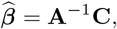

with **A** a *J × J* matrix and **C** a *J* -vector given by

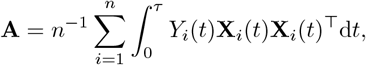

and

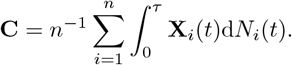

Furthermore the variance-covariance matrix of 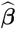 can be consistently estimated by *n*^−1^**A**^−1^**BA**^−1^, with **B** a *J × J* matrix given by

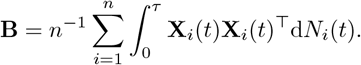

Replacing **X**_*i*_(*t*) by *Ī*(*t*), *Y*_*i*_(*t*) by *S*_*i*_(*t*), and summing over *i*, we get simpler versions of the matrices **A** and **B** and of the vector **C**, namely

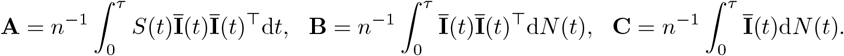

The code below implements these estimators and their standard errors, first in group 1, then in group 2.

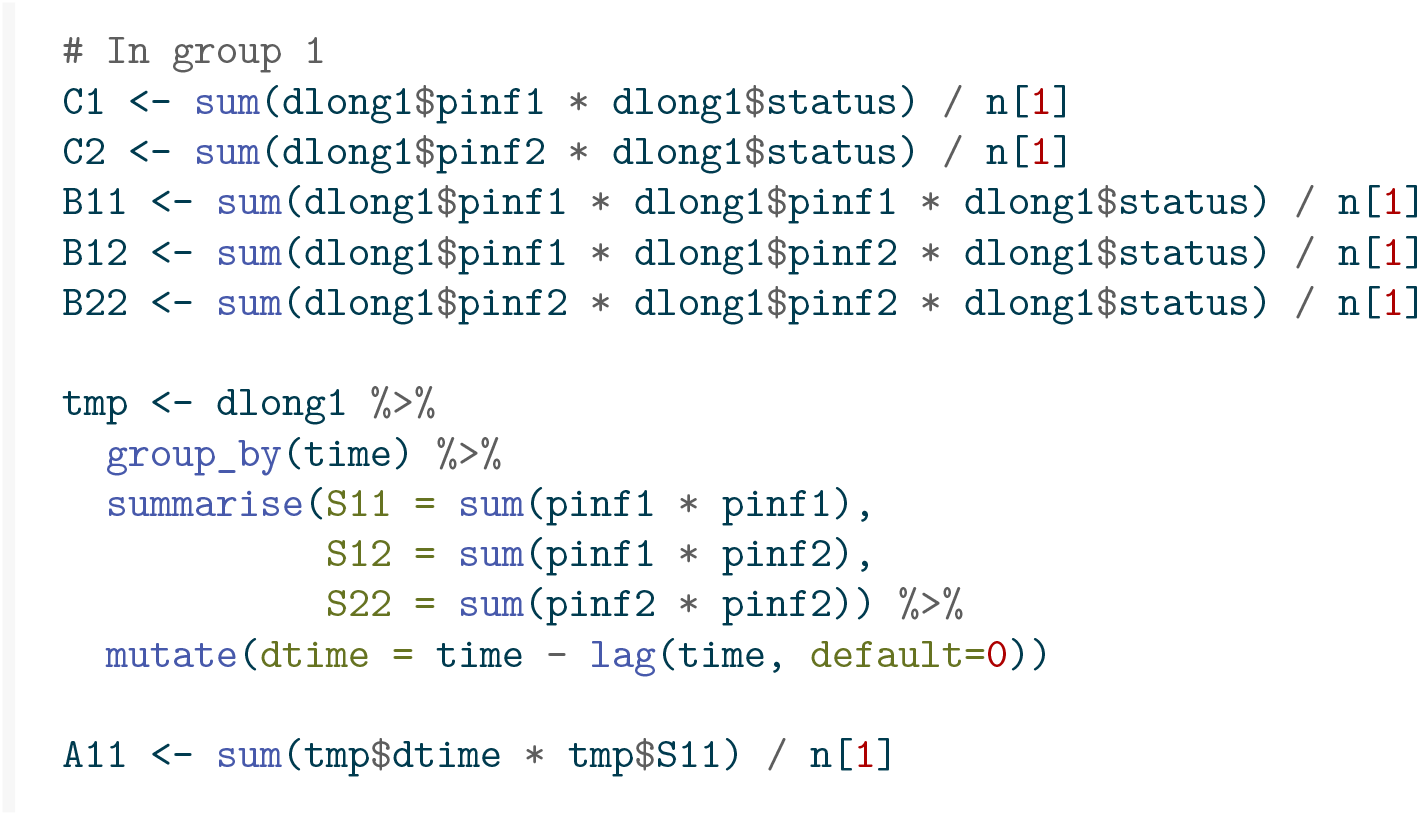
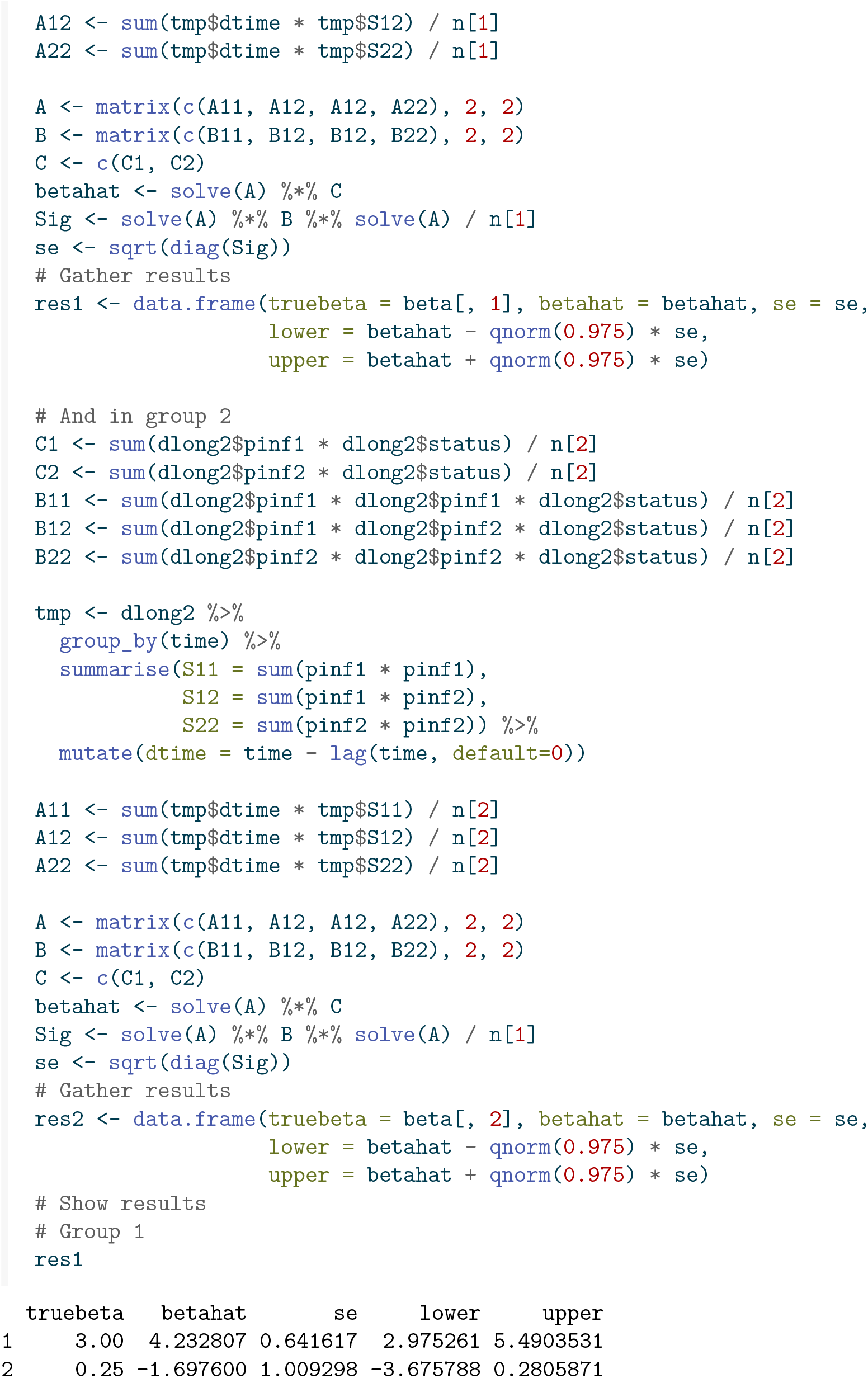
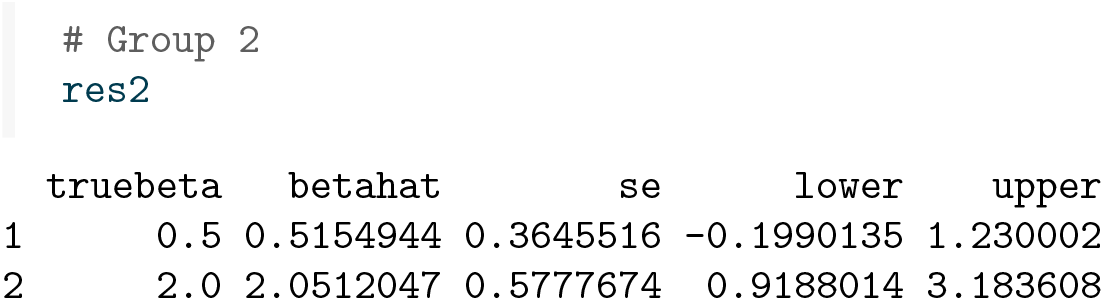

Two things are worth noting. The first is that the estimates are reasonably close to the true values, and well within the 95% confidence intervals. The second is that one of the coefficients is estimated to be negative, which of course is not desirable. It would be of interest to estimate the *β* coefficients under a non-negativity constraint, as pursued in Lu, Goeman, and Putter (2023).

### 4.4 Hybrid (Cox-Aalen) models

As we have seen, the effect of an intervention is most naturally incorporated as a multiplicative effect. The same goes for the effect of measured characteristics of the susceptible individuals, like gender, or perhaps known risk factors for infection. If we want to incorporate such effects multiplicatively and additionally have a structured population that would call for an additive hazards structure, then hybrid models would be of interest where the hazard of subject *i* in group *j* takes the form

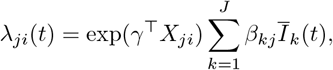

with *X*_*ji*_ a vector of baseline covariates of subject *i* in group *j* would be of interest. This type of model is known as a Cox-Aalen model, and has been studied by Scheike and Zhang (2002). We will not pursue this method in this paper.

### 4.5 Heterogeneity in susceptibility to infection

It is a huge simplification to assume that each susceptible individual is equally susceptible to becoming infected, or that each infected individual is equally likely to infect others. With individual knowledge of covariates, these could be incorporated into the survival analysis models, be they additive hazards, Cox or Poisson models. In the absence of such information, a natural extension to the models considered is to add individual random effects expressing such heterogeneity. In survival analysis, models incorporating such random effects are known under the term *frailty models*, see for instance Balan and Putter (2020). It is possible to fit such frailty models also for SIR models.

The function below generates completely observed data from an SIR model where the transmission parameter associated with a susceptible individual equals *βZ*, with *Z* a gamma random variable with mean one and variance fvar. Thus, the mean transmission parameter over the population of susceptibles equals *β*, but susceptible individuals differ in their degree of susceptibility to infection

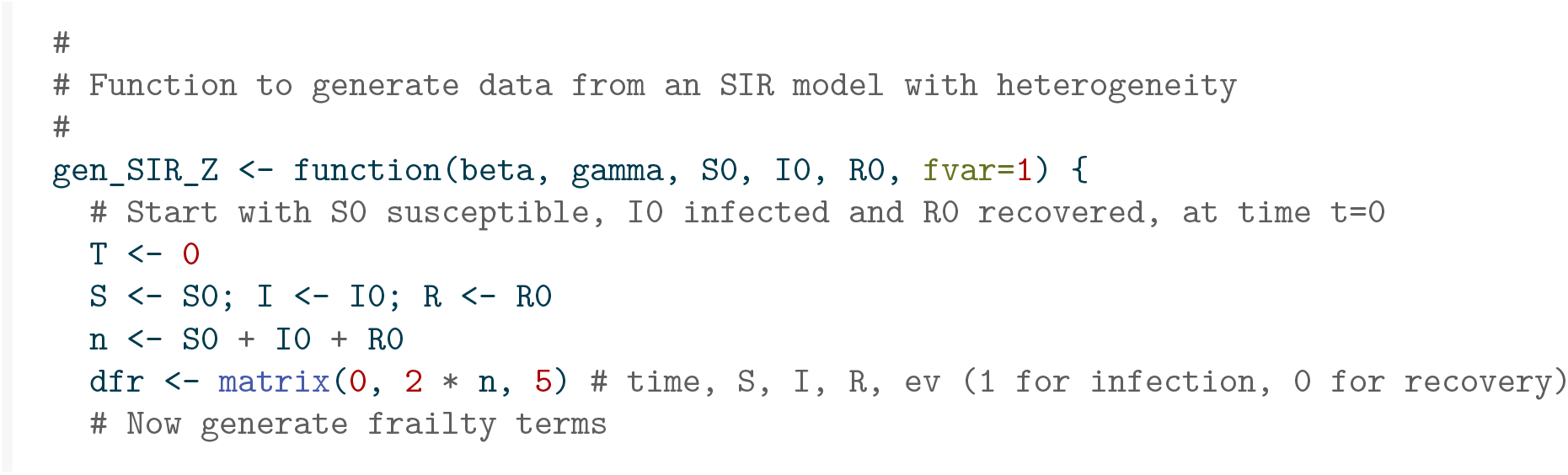
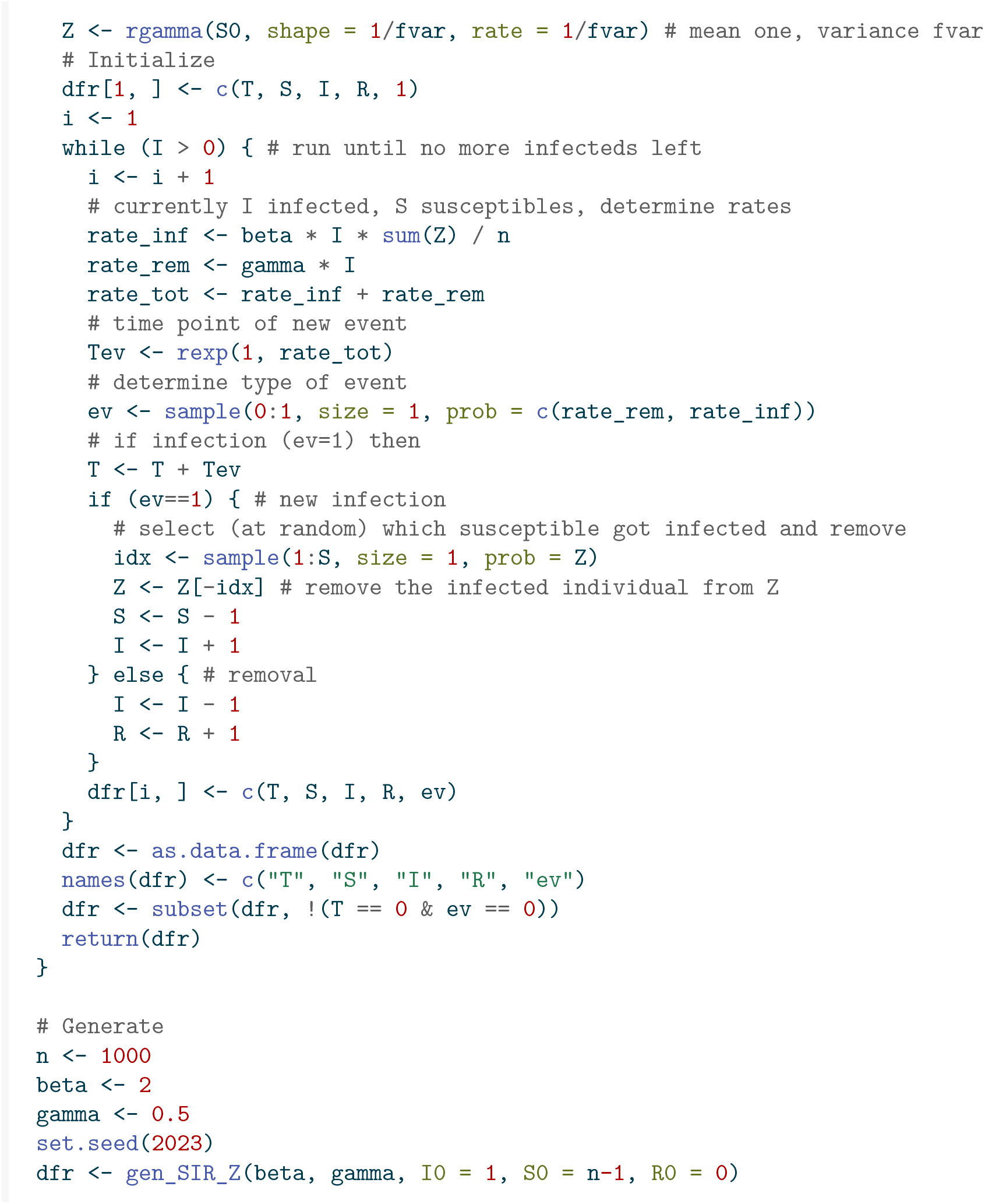

This heterogeneity in susceptibility leads to an epidemic with considerably fewer infections compared to one with the same *β* and *γ* and no such heterogeneity, as can be seen from Figure 11.

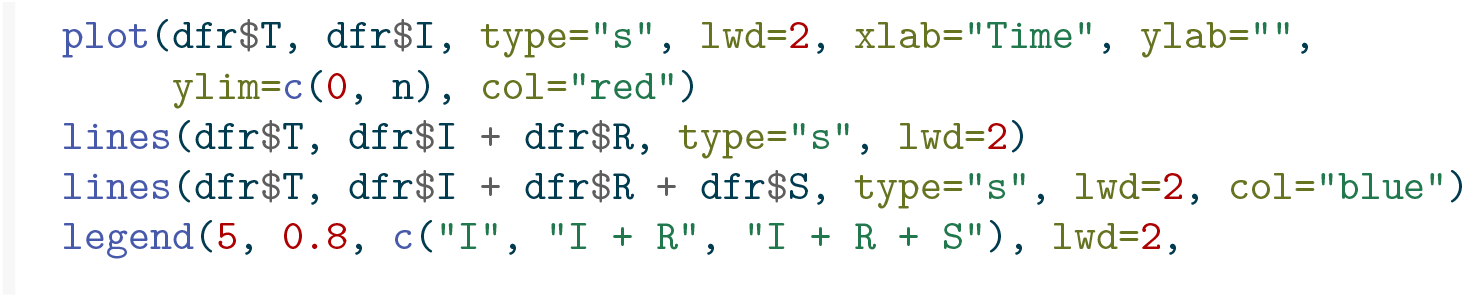
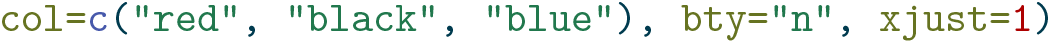

**Figure 11:**
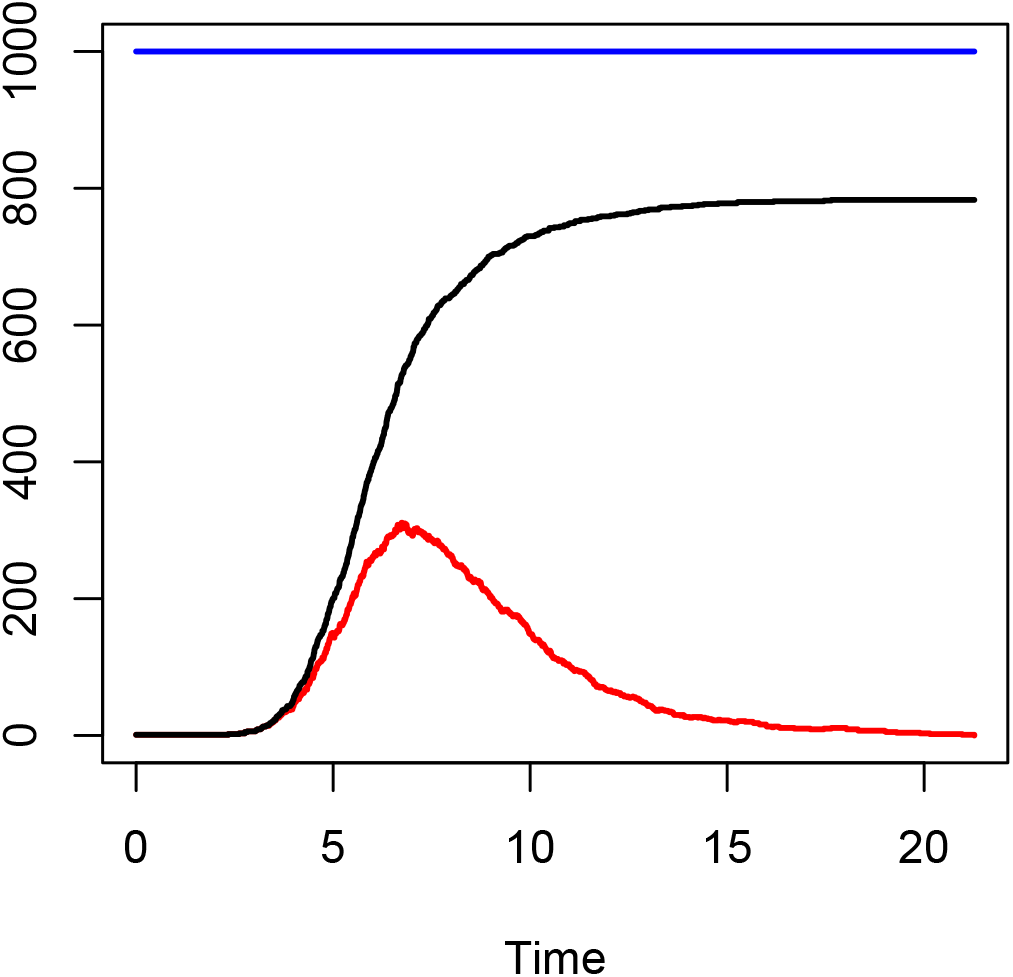
Stacked plot showing the number of susceptible, infected and recovered individuals over time from a stochastic SIR model with heterogeneity in susceptibility.

It can be shown by Jensen’s inequality that the assumption that all individuals are equally susceptible leads to an upper bound for the final proportion infected, see for instance Katriel (2012) and Miller (2012).

Let us now estimate *β* from this generated data, first assuming it is time constant.

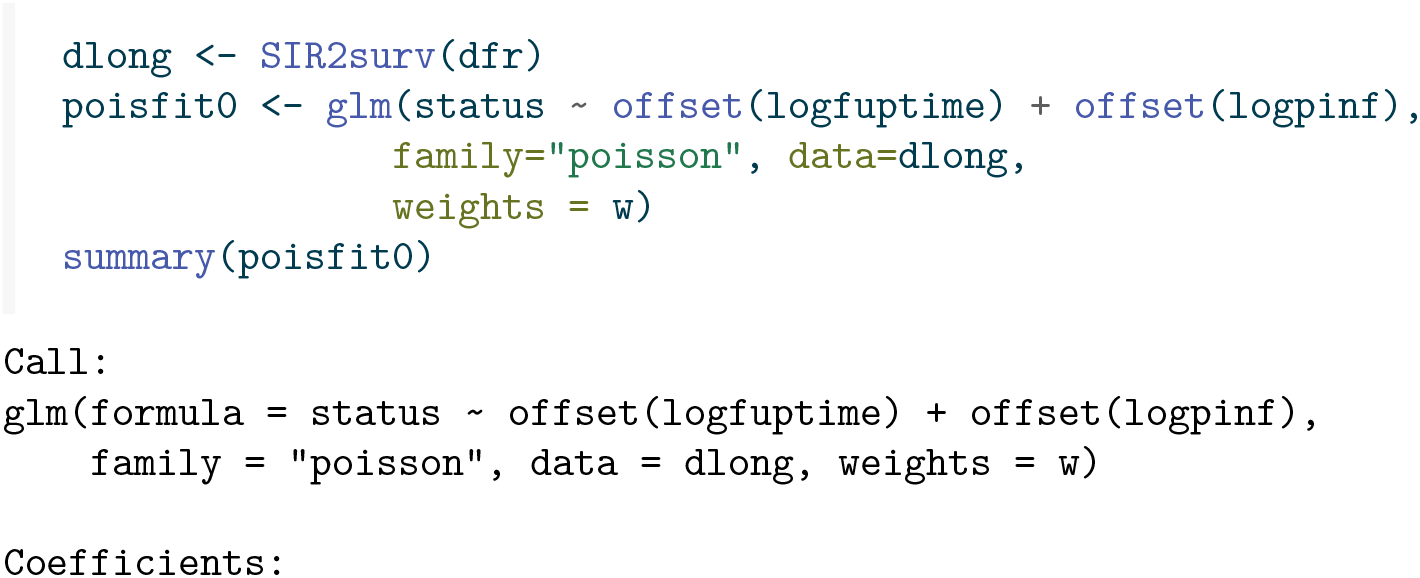
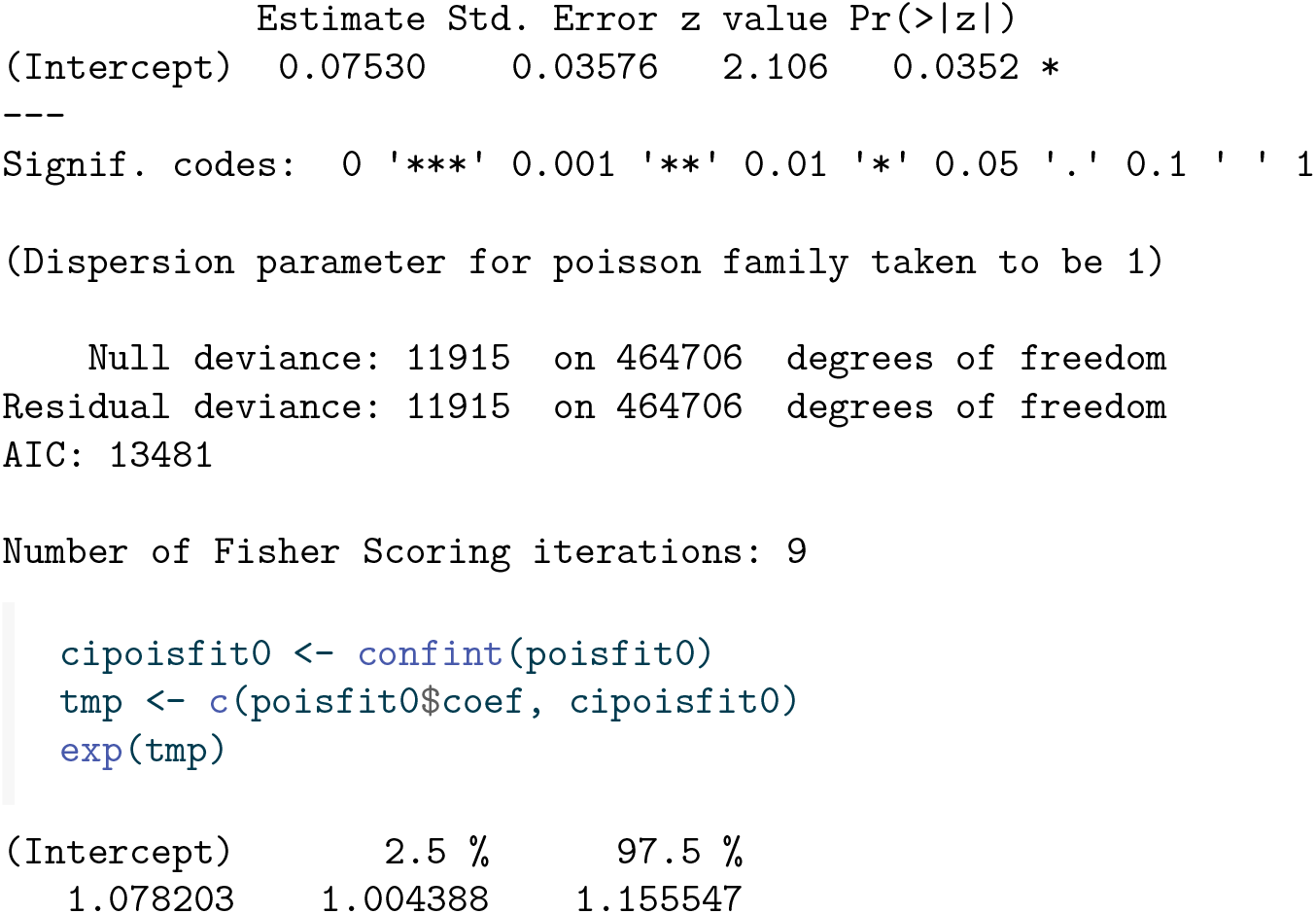

The result, 1.08, is considerably lower than the true value 2, caused by the fact that the most susceptible individuals get infected first, resulting in less susceptible individuals remaining in the susceptible pool over time. As in standard survival analysis, the presence of the frailty terms induces time-varying behaviour of *β*. This is indeed picked up using Poisson regression with splines.

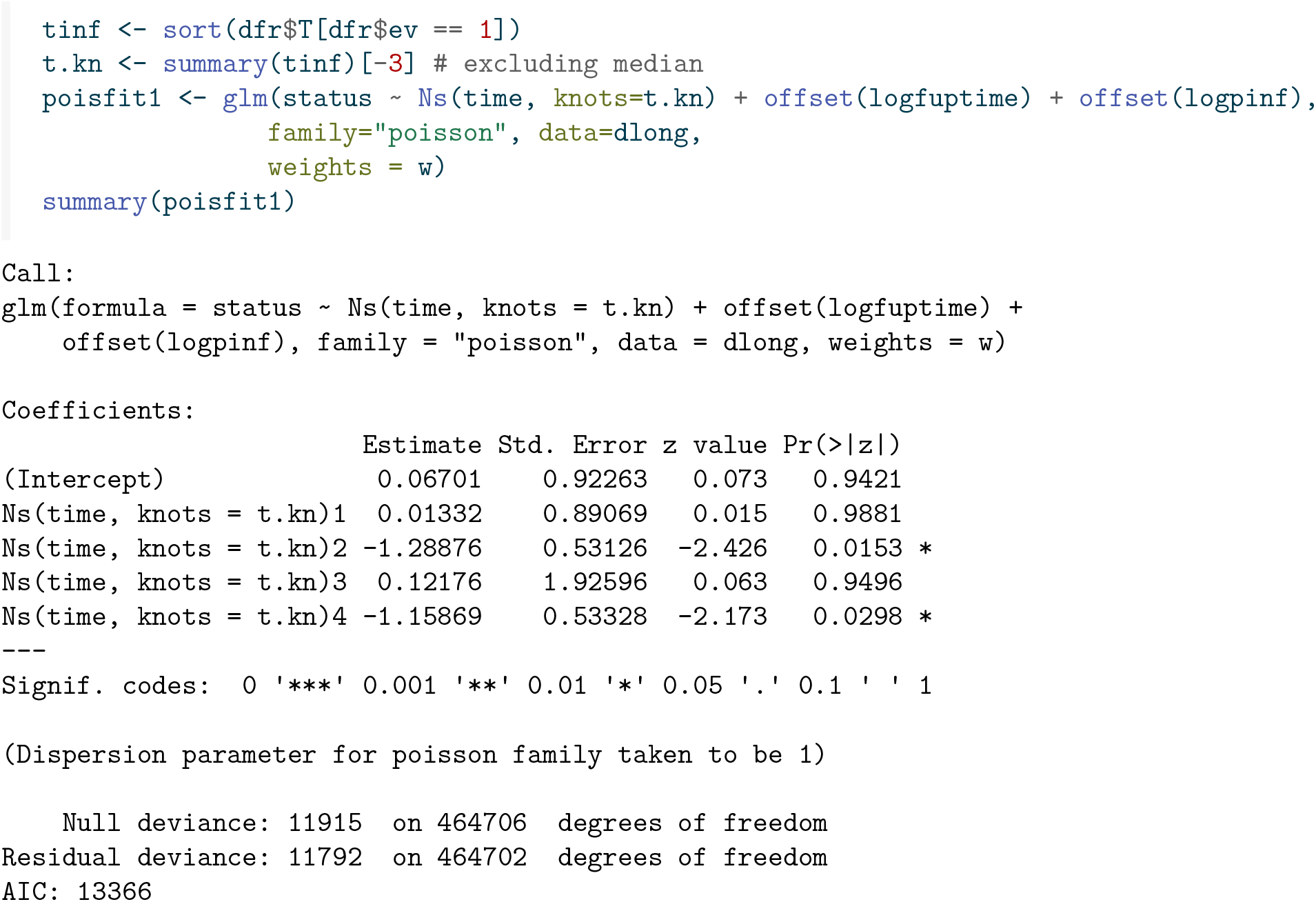
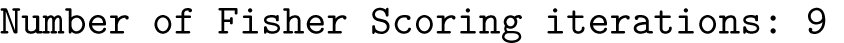

We indeed see non-constant β(*t*) in Figure 12.

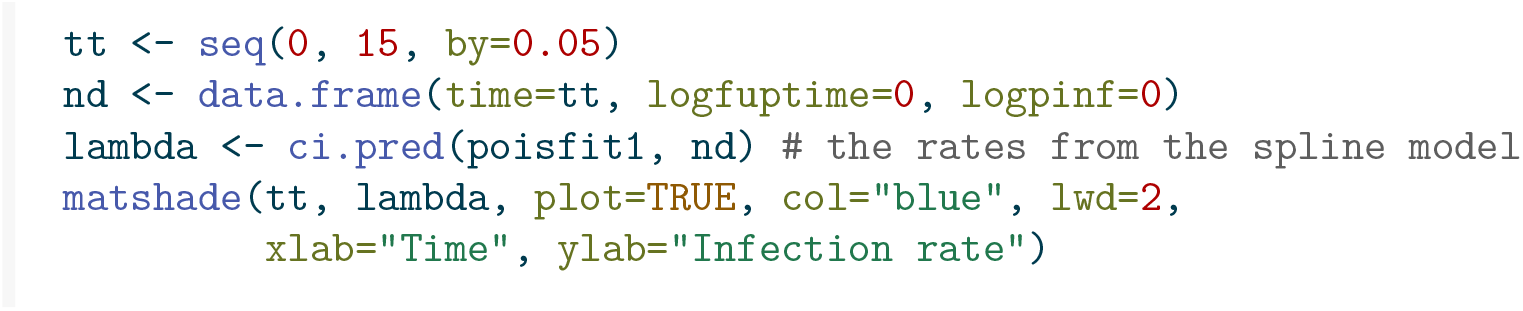

**Figure 12:**
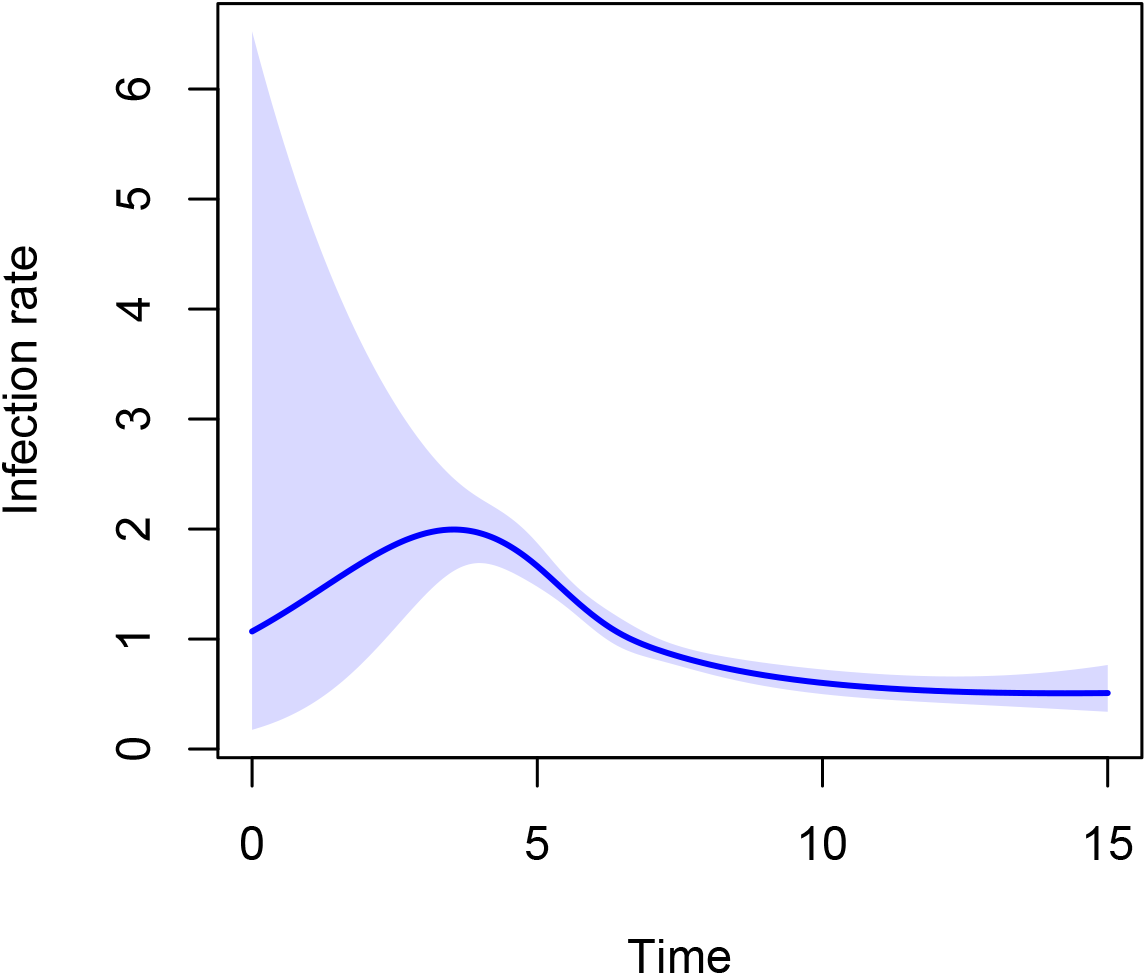
Estimate of *β*(*t*) obtained using Poisson regression with cubic splines from an SIR model with heterogeneity in susceptibility.

Fitting a Cox model with individual frailty terms will not work, because the individual model with individual frailty is not identifiable in the absence of time-fixed covariates. A Poisson model with individual frailty terms seems like a viable option, but all the methods in https://rpubs.com/kaz_yos/glmm1 throw errors. It should be feasible though to write a dedicated EM-algorithm for this, but this is not pursued here.

## 5 Challenges

The type of data that we considered in this paper is too idealized in the sense that we typically do not know the time of infections exactly and we do not know the number of infectious and susceptible individuals exactly. Numbers of infections are usually aggregated over days or weeks. Moreover they are typically reported with some delay and they are incomplete. Methodology needs to be extended to deal with these more realistic data settings. We will not cover all of these aspects here, but we will show how the methods in this paper can still be used to estimate the transmission parameter *β* in case the (correct) probability distribution of the time to recovery after infection is known, and aggregate data on the daily number of newly infected individuals is reported, as was the case for the COVID-19 infection. For now we assume there is no reporting delay or incompleteness.

We start from the fully observed data used before. To make it roughly in line with the recent COVID-19 epidemic, based on Figure 2 it seems reasonable to say that the time unit there is months, and that we have daily updates of the number of new infections. Let us say that after 10 infections the outbreak is “detected” (at day 41), and that we set this to day 0 and start reporting daily new infections after that.

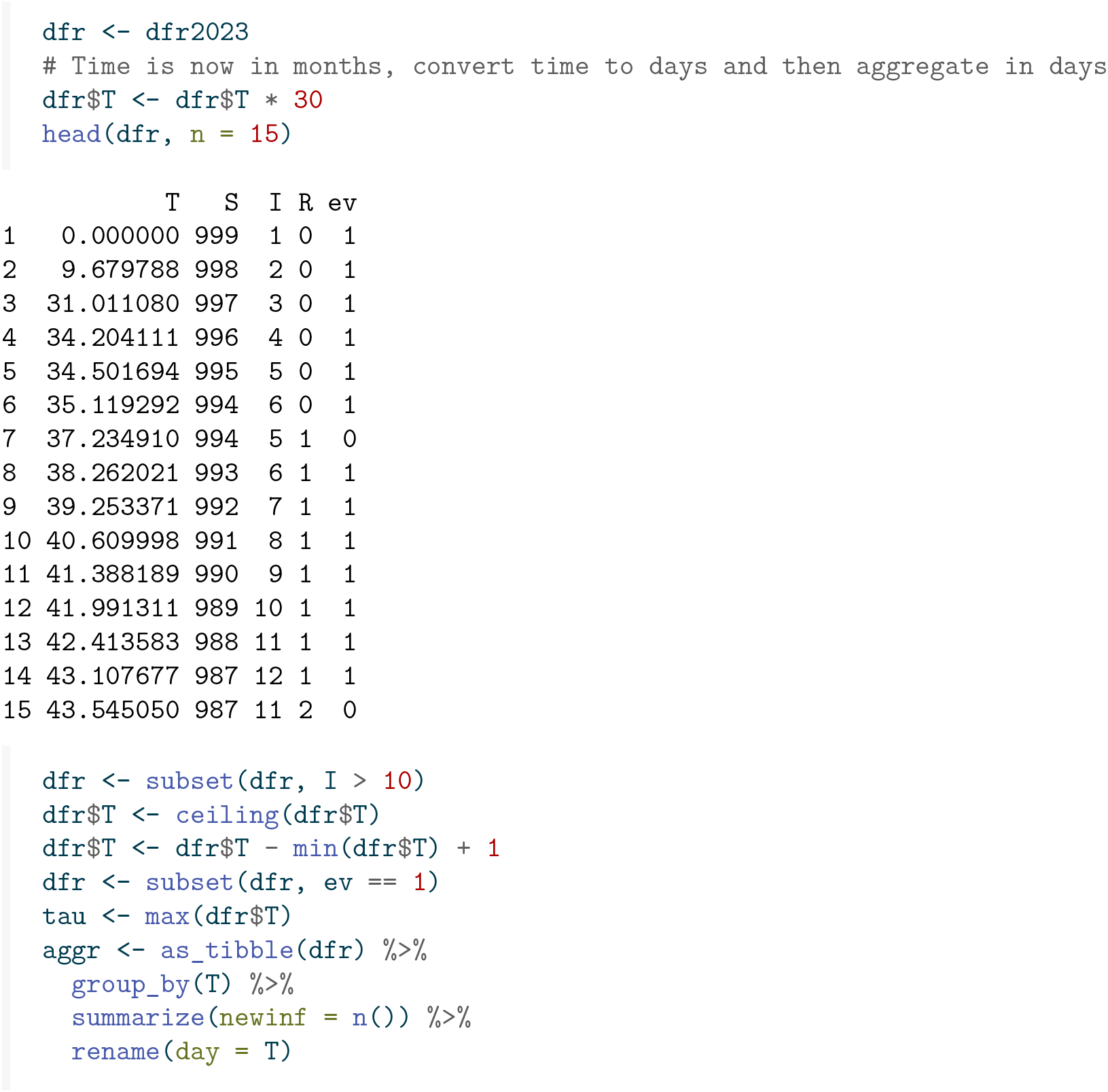

This is what these aggregate data look like, with newinf the number of new infections reported each day.

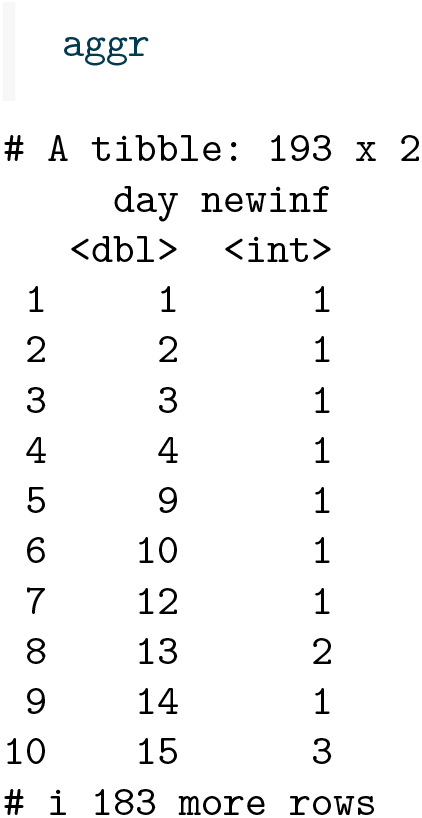

Using the information on time to recovery, each newly infected individual on day *d* will be counted as an infected individual on day *d* + *j* with probability *p* = *P* (time to recovery ≥ *j*). The expected number of infected individuals on day 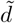 then is a sum of the number of newly infected individuals on day 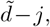 weighted by *p*_*j*_. We can then create a daily analysis data set where each day is represented by a number of individuals becoming infected with `status` = 1 (the number of newly infected individuals on that day), and a number of individuals that are susceptible and did not get infected on that day with `status` = 0 (the population size *n* minus the cumulative number of infected individuals until and including that day).

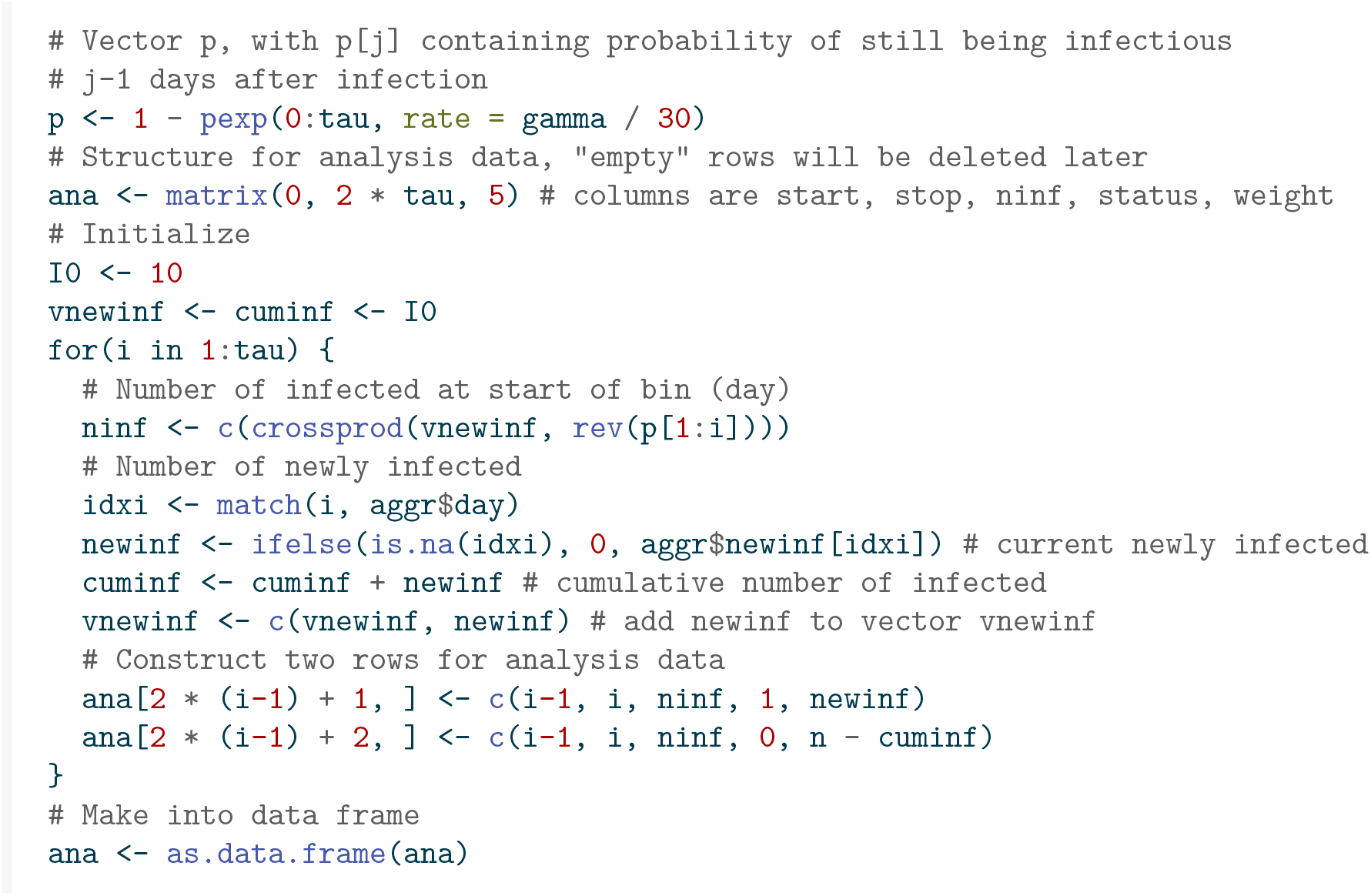
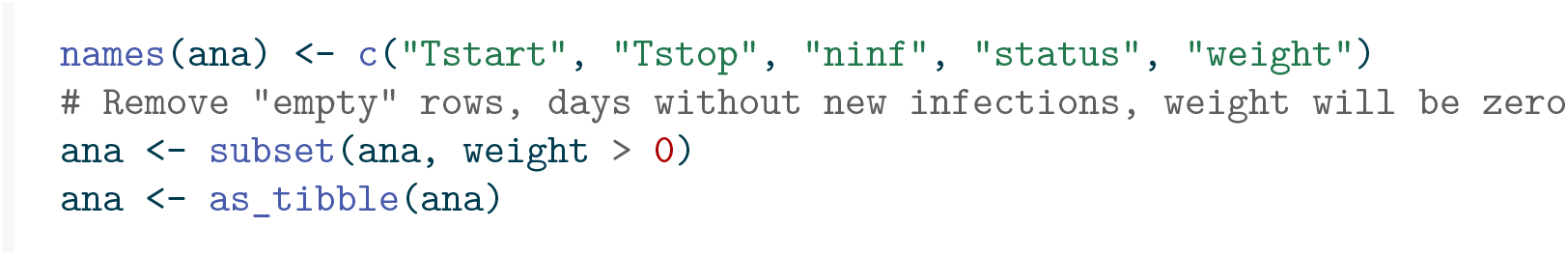

This is what the data look like for the first seven days.

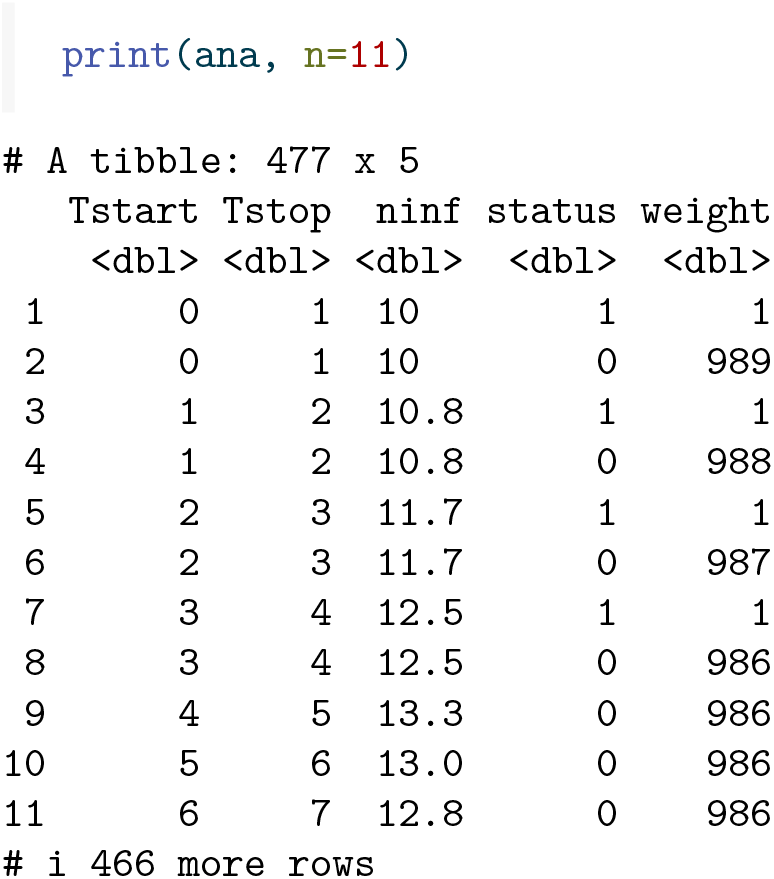

Note that on days 5 through 7 no new infections occurred, we therefore see no rows on these days with `status = 1` .

Figure 13 shows the expected number of infected individuals over time (in days since the outbreak was detected).

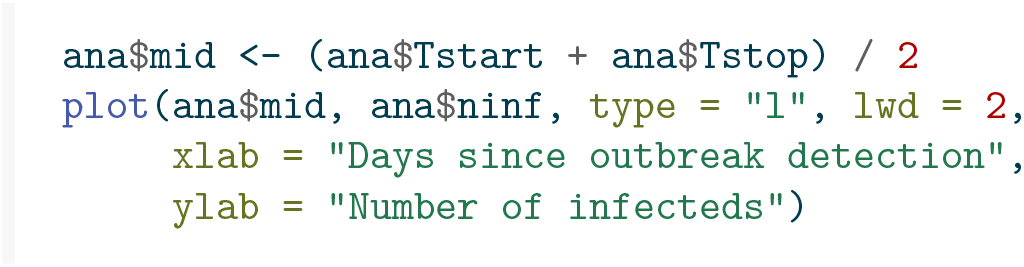

**Figure 13:**
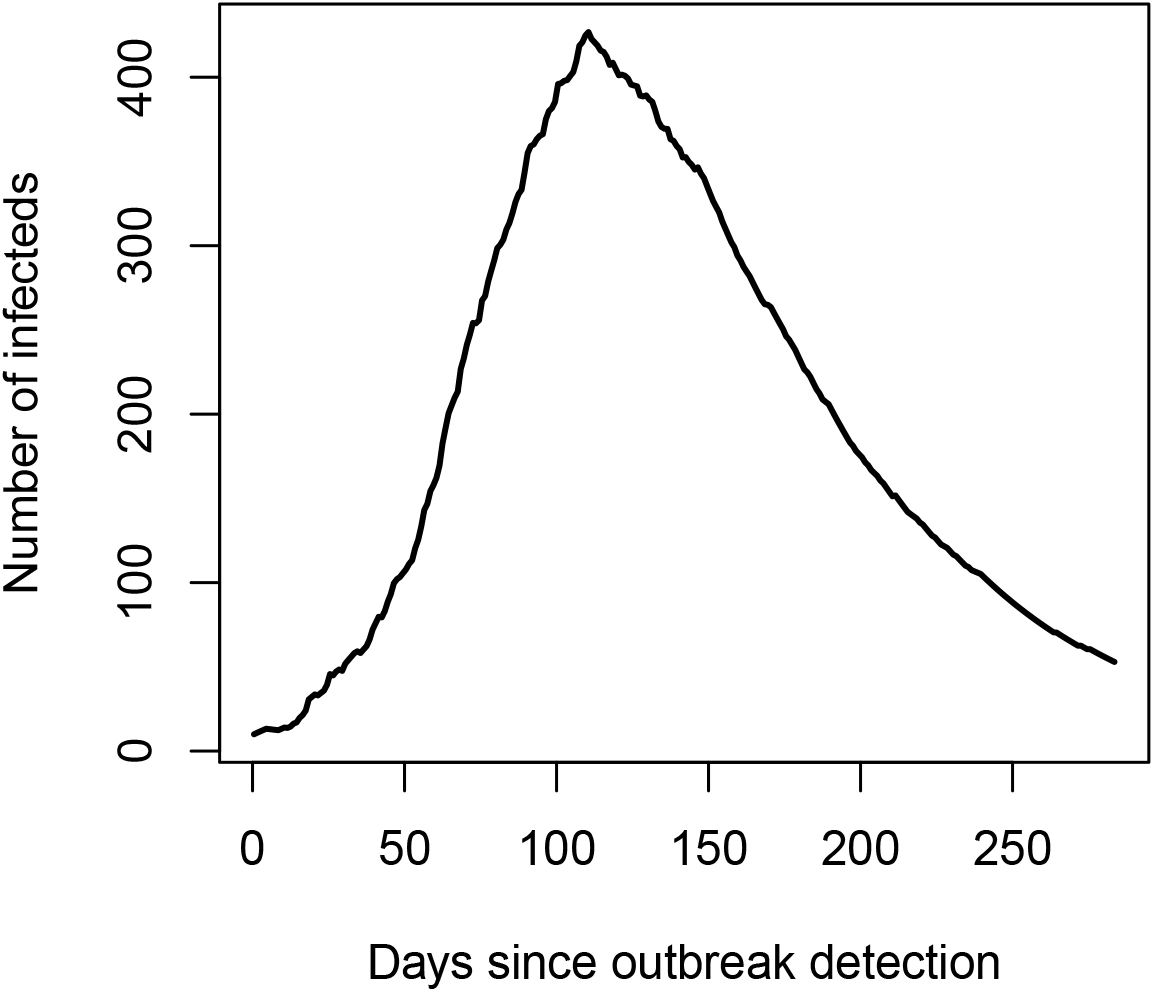
Expected number of infecteds over time.

After calculating `pinf`, the proportion of infected individuals, and `fuptime`, the time between the start and end of the reporting time period, which was one day, 1/30 of a month, we can then use Poisson regression with log link and `logpinf` and `logfuptime` as `offsets`, to obtain an estimate of *β*.

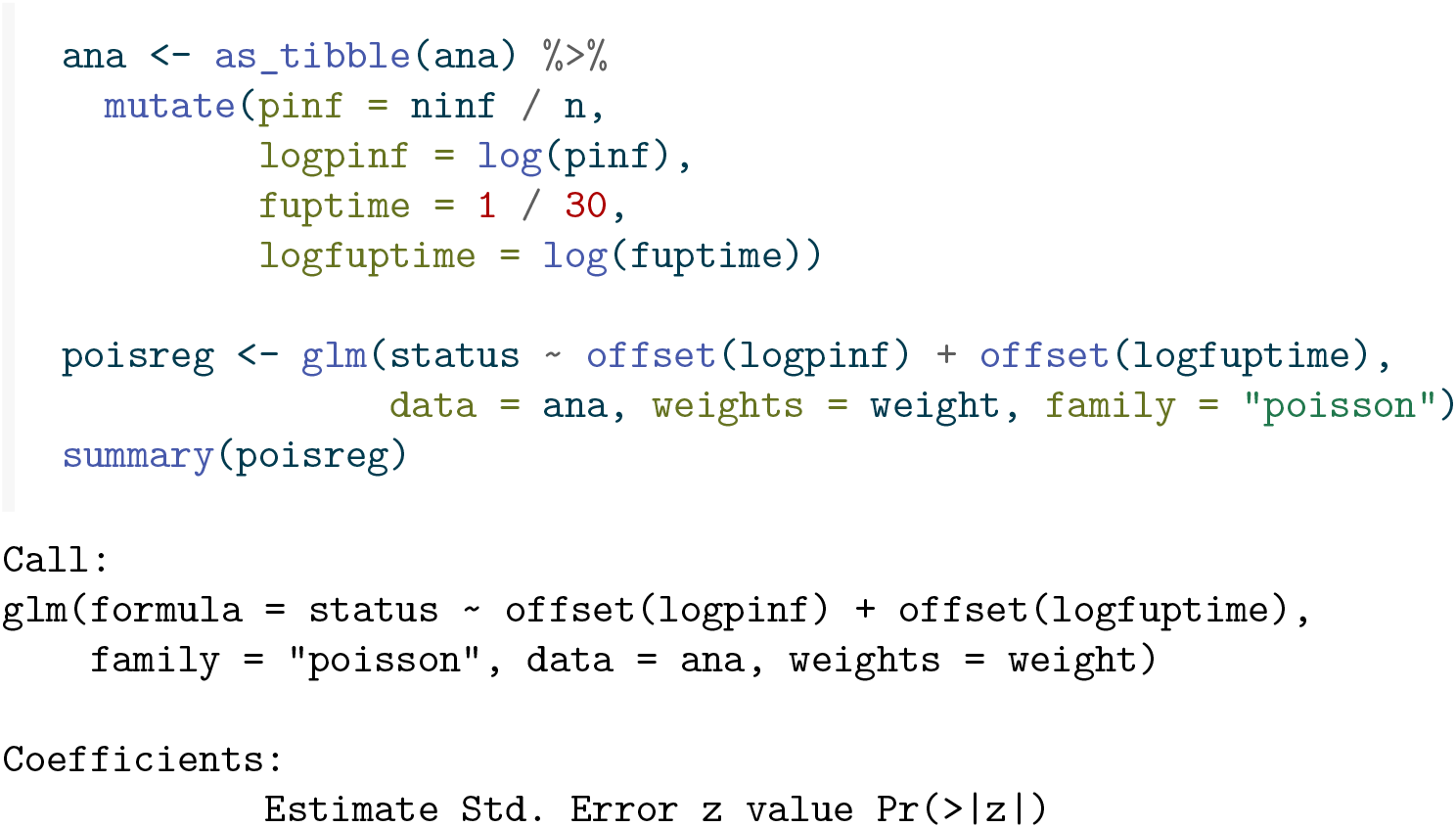
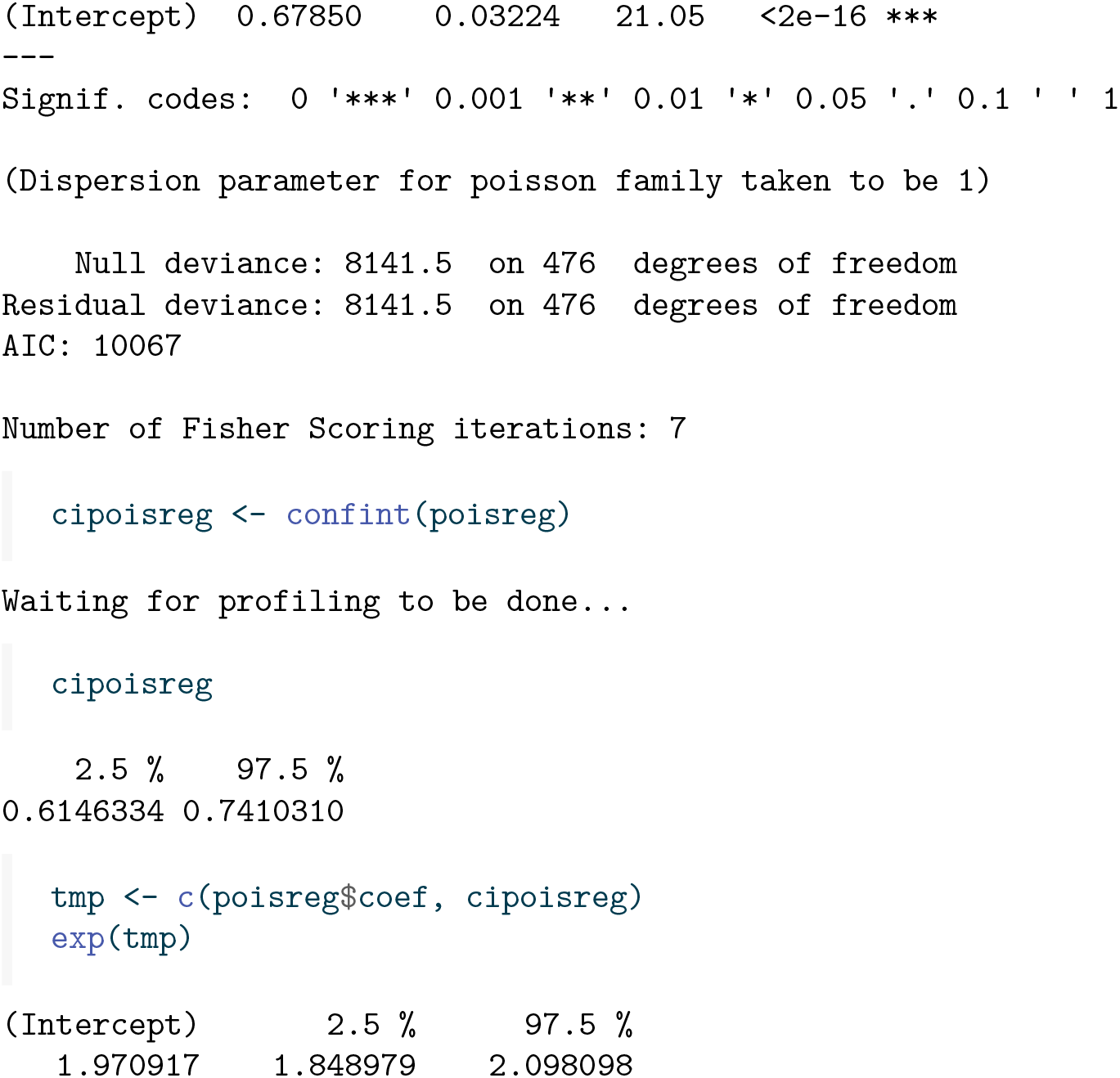

The result again is very close to the true value of *β* = 2. The Poisson regression also gives a confidence interval. Note, however, that the estimate of the standard error of 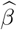 is too optimistic, for two reasons. First, assuming known time-to-recovery distribution, the randomness in the actual time-to-recovery is ignored and replaced by their expectations. Second, the time-to-recovery distribution is typically estimated with considerable uncertainty in itself. These sources of randomness need to be taken into account to obtain correct standard errors and confidence intervals. This is outside the scope of this manuscript.

## 6 Discussion

In this manuscript we have shown how standard methods from survival analysis can be used to estimate pivotal quantities in SIR models. In particular we have focused on estimating the transmission parameter in the SIR model. We have illustrated the use of multiplicative models like the Cox model and Poisson regression, and of the additive hazards model, and we have argued for the usefulness of the Cox-Aalen model, which is a hybrid of multiplicative and additive models. The possibility of using these standard models with the wide availability of software to elucidate underlying pivotal parameters opens possibilities in many situations, for instance in structured and/or clustered data.

Interestingly, in contrast to what seems to be implicit in the literature (Kenah 2011, 2013, 2015), we do not need information on who infected whom, but correct knowledge of the number of susceptible and infectious individuals over time is needed. In most realistic situations this knowledge is not readily available and needs to be further estimated from the available data and assumptions.

Clearly, more work is needed. First and most importantly, the data challenges need to be addressed. Issues like incompleteness of reporting of infections, reporting delay will severely complicate reliable knowledge of the number of infected and infectious individuals over time. Second, the issue of heterogeneity in susceptibility is of major interest. Using frailty models seems a very promising way of estimating the extent of variability in susceptibility in the population. Third, while multiplicative models have already been used to estimate the effect of interventions in slowing down the spread of an infection, additive hazards models have to the best of our knowledge not been used in infectious disease models. Finally, the use of hybrid multiplicative - additive models such as the Cox-Aalen model seems particularly attractive.

## Data Availability

All data produced in the present study are available upon reasonable request to the authors

## Notes

### Competing Interest Statement

The authors have declared no competing interest.

### Funding Statement

This study did not receive any funding

### Summary of Updates

Corrected Inf appearing in additive hazards results in Section 3.1, and corrected Section 5 (was using data with heterogeneity for analysis instead of data of Section 2). Plus very minor text changes.

